# BEAT Tuberculosis: a randomized controlled trial of a 6-month strategy for rifampicin-resistant tuberculosis

**DOI:** 10.1101/2025.05.04.25326549

**Authors:** Francesca Conradie, Tasnim Badat, Asanda Poswa, Shakira Rajaram, Shaneen Kooverjee, Gary Maartens, Graeme Meintjes, Norbert Ndjeka, H.S Schaaf, Jennifer Hughes, Pauline Howell, Patrick Phillips

## Abstract

**Background:** Safer, more effective treatment regimens for rifampicin-resistant tuberculosis are needed. We evaluated a novel six-month RR-TB treatment strategy.

**Methods:** We conducted a phase III, open-label, multi-centre, pragmatic, noninferiority randomised-controlled strategy trial in South Africa to assess the effectiveness and safety of a 6-month strategy for individuals with pulmonary RR-TB. Participants aged six years and older, including pregnant and breastfeeding women and those with fluoroquinolone resistance, were randomised to receive either a 6-month strategy regimen of bedaquiline, delamanid, linezolid, levofloxacin and /or clofazimine (BDLLfxC) (Study Strategy) or the then-current South African nine-month, or longer regimen(Control Strategy). Treatment for both groups was adjusted based on the second-line drug susceptibility test results. The primary effectiveness measure was successful treatment completion at the end of treatment and a successful end-of-follow-up (76 weeks) outcome, compared using an adjusted risk difference with a 10% non-inferiority margin.

**Results:** Of 432 individuals screened, 403 were randomised (203 and 200 to the control and study strategies, respectively). One participant in the study strategy arm never started treatment. Successful outcomes were achieved in 172/200 (86.0%) and 174/202 (86.1%) in the control and study strategies, respectively; the adjusted risk difference was -0.2% (95% CI -6.9% to 6.5%), demonstrating non-inferiority (p=0.0014 test for non-inferiority). Seventy-four (37.0%) and 63 (31.2%) in the control and study strategies, respectively, experienced severe adverse events during treatment; 10 participants in each strategy died.

**Conclusions:** The 6-month study strategy regimen demonstrated non-inferior effectiveness and safety comparable to the South African standard-of care-TB regimen.

*Trial registration number:* NCT04062201

## Introduction

The World Health Organization (WHO) estimated that 400,000 people fell ill with rifampicin-resistant TB (RR-TB) in 2023.(1) Until recently, treatment regimens for RR-TB were lengthy, lasting from 9 to 18 months, and with up to seven medications. The pill burden and complexity of these regimens, coupled with associated adverse events, led to poor treatment outcomes.(2)

Over the past decade, three new drugs—bedaquiline, pretomanid, and delamanid—for RR-TB have been approved by at least one stringent regulatory authority, including the South African Health Products Regulatory Authority. In addition, repurposed medications such as linezolid, levofloxacin, moxifloxacin, and clofazimine have shown improved outcomes in treating RR-TB.

Several trials, including Nix-TB(3), ZeNix(4), TB-PRACTECAL(5) and endTB(6) demonstrated 6-month regimens containing bedaquiline, linezolid, and pretomanid combinations, leading to 80-90% cure rates. As a result, the WHO revised its guidelines for treating RR-TB in 2022 to include a six-month regimen of bedaquiline, pretomanid, linezolid, and moxifloxacin (BPaLM) as the recommended standard of care(7). There are, however, notable gaps: pretomanid is currently not recommended for children under 14 years of age or pregnant and breastfeeding women by the WHO, FDA, EMA or SAPHRA.

Building on this, the BEAT Tuberculosis trial was conceptualised. We evaluated a bedaquiline, delamanid, and linezolid-based regimen with levofloxacin or clofazimine or both (BDLLfxC) as the Study strategy compared to the Standard of Care Strategy in South Africa as the Control strategy. Our objective was to evaluate the efficacy and safety of an inclusive treatment strategy for the treatment RR-TB, including fluoroquinolone-resistant RR-TB, with regimens that can be given to most individuals.

## Methods

### Study design

BEAT Tuberculosis was a phase III, open-label, multi-centre, pragmatic, randomised-controlled strategy trial to demonstrate non-inferiority. We enrolled a broad group of participants with pulmonary RR-TB.

### Study Participants

Participants were recruited from two urban sites in South Africa. Inclusion criteria were pulmonary TB with resistance to at least rifampicin by genotypic or phenotypic drug susceptibility testing (DST), age six years or older, and pregnant and breastfeeding women (Full inclusion and exclusion criteria Supplementary Appendix. Section 2.6).

### Enrolment and Interventions

Participants were randomly allocated to receive either strategy, stratified by site and HIV status using blocked randomisation with blocks of varying sizes. This was implemented using a web-based system to ensure allocation concealment before enrolment.

The study strategy comprised five drugs: bedaquiline, linezolid, delamanid, levofloxacin, and clofazimine (BDLLfxC). Upon receipt of the fluoroquinolone DST result, either levofloxacin (if fluoroquinolone-resistant) or clofazimine (if fluoroquinolone-susceptible) was discontinued (or not initiated if results were available at the time of randomisation). The control strategy adhered to South African standard treatment for RR-TB (at the time of enrolment), a nine-month, all-oral regimen or a longer individualised regimen if resistance to fluoroquinolones was identified. (Full information regimens see Supplementary Appendix 2.5) Scheduled visits took place every two weeks for the initial eight weeks, followed by visits every four weeks until treatment completion. Post-treatment appointments occurred monthly for the first three months and subsequently every three months. All participants were monitored for 76 weeks. Microbiological specimens were processed as routine specimens at the National Health Laboratory Service (NHLS). Xpert MTB/RIF Ultra typically served as the first diagnostic specimen. Line-probe assays (GenoType MTBDR*plus* and MTBDR*sl*, Life Science, Nehren, Germany) were performed to determine susceptibility to isoniazid, second-line injectables, and fluoroquinolone antibiotics. In acid-fast bacilli smear-positive specimens, these assays were conducted on the clinical isolate, while in smear-negative specimens, they were performed on cultured isolates. Additionally, phenotypic bedaquiline susceptibility was evaluated on fluoroquinolone-resistant isolates. Sputum samples for mycobacterial cultures were collected monthly during treatment, three months post-treatment, and then every three months until 76 weeks post-randomisation. (For a comprehensive schedule of events, refer to Supplementary Appendix 2.7).

### Therapy Adherence support

Most participants were treated as outpatients unless inpatient care was needed. Participants selected a family or community member to facilitate directly observed therapy (DOT). This community-based DOT supporter had standardised training and was asked to sign an adherence log. Medication was packed into daily labelled bags, which the participants were asked to return when they came for visits. If missed doses were noted, the DOT supporter was contacted and asked to intervene.

### Safety and dose modifications

Adverse events were recorded at every study visit, and laboratory safety tests were performed at every scheduled visit during treatment. Adverse events were graded using the Division of AIDS (DAIDS) Table for Grading the Severity of Adult and Pediatric Adverse Events.(8). Electrocardiogram (ECG) monitoring, visual acuity examinations, and specific assessments for peripheral neuropathy using a Brief Peripheral Neuropathy rating scale were performed at scheduled intervals.

In the study strategy, no changes to the medication dosing were allowed unless the weight category changed. If a participant experienced an adverse reaction to linezolid, the drug could be stopped either temporarily or permanently. After the adverse reaction improved to Grade 1 or the baseline level, linezolid was resumed at a dose of 600mg. There was no provision for dose reduction following a linezolid-related adverse reaction.

### Outcome Measures

The primary efficacy endpoint was a successful end of treatment (based on WHO programmatic definitions) followed by a successful end of follow-up outcome.(9) (For detailed definitions. Supplementary Appendix.2.9

### Study Oversight

All participants provided written informed consent, with assent from minors and consent from parents or guardians. An independent data monitoring committee reviewed safety and efficacy data every six months. The study was approved by the Human Research Ethics Committee of the University of the Witwatersrand (FWA00000715). The principal investigator and trial steering committee managed the trial, and the authors guaranteed the data’s accuracy and protocol fidelity.

### Statistical Analysis

The primary effectiveness analysis is based on the intention-to-treat analysis (ITT) population, including all randomised participants.

For the primary analysis, the risk difference between participants with a successful outcome at the end of treatment and follow-up at 76 weeks after randomisation between the control and study strategies was calculated with 95% confidence intervals (CI), stratified by HIV and site using Cochran-Mantel-Haenszel weights. (10) Non-inferiority was indicated if the upper bound of the 95% CI was less than the 10% margin of non-inferiority.

The proportion of participants with a primary safety outcome, defined as any grade 3 to 5 adverse event up to 14 days after the last dose of any anti-TB medications in a participant who took one or more days of treatment, was estimated, and the difference in proportion calculated with 95% CIs.

## Results

### Participants

Four hundred thirty-two participants were screened, and 403 (93%) were randomised between August 2019 and October 2022 (Figure 1). The median age was 35.0 years, with 30 (7%) participants under 18 (the youngest was eight). Out of the total, 205 (51%) participants were living with HIV. Body mass index (BMI) was less than 18.5 kg/m^2^ among 167 (41%) participants. Baseline characteristics were well balanced between strategies (Table 1). Of 170 (42%) female participants, 9 (6%) were pregnant at baseline.

**Table 1:**
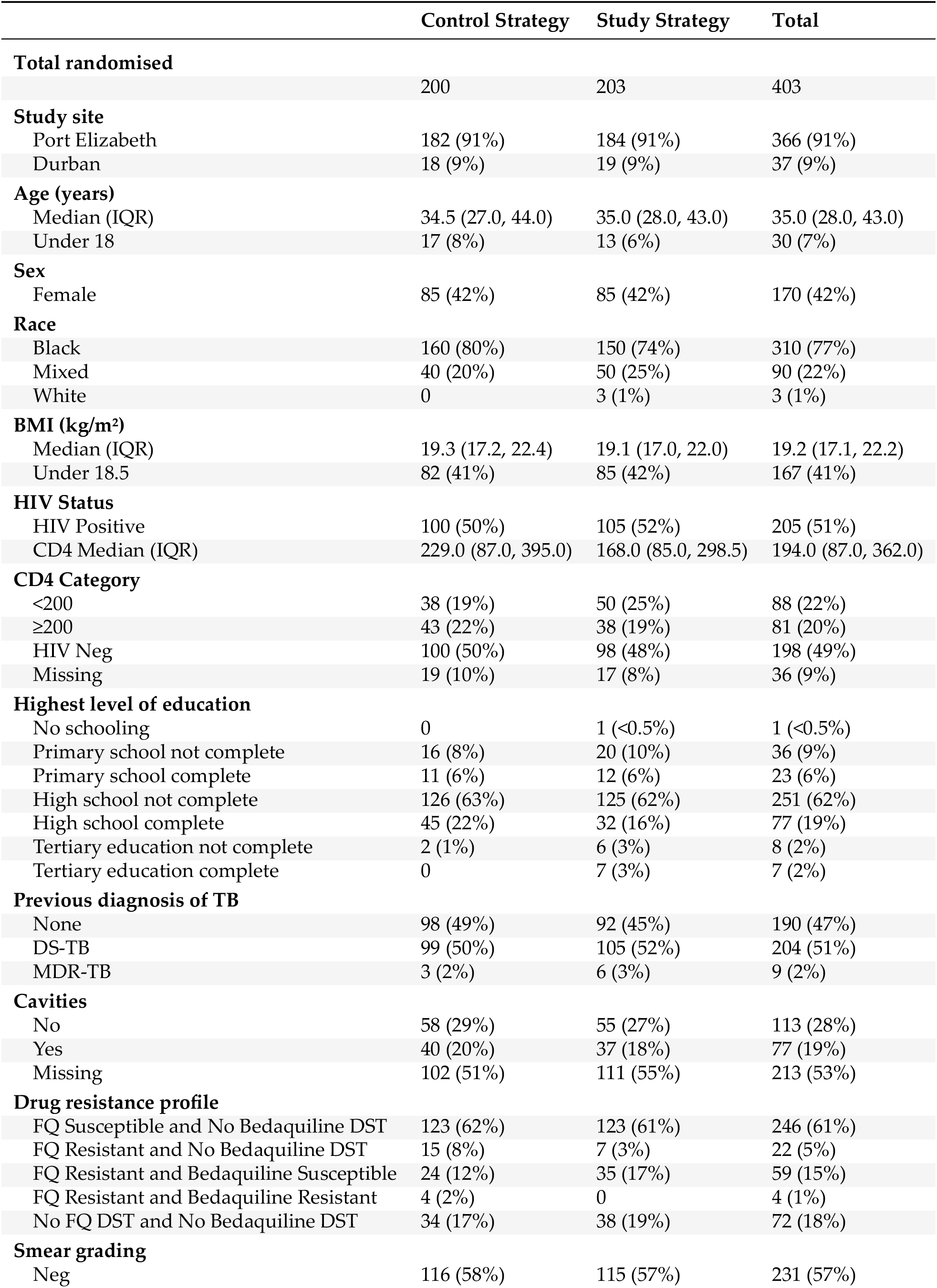
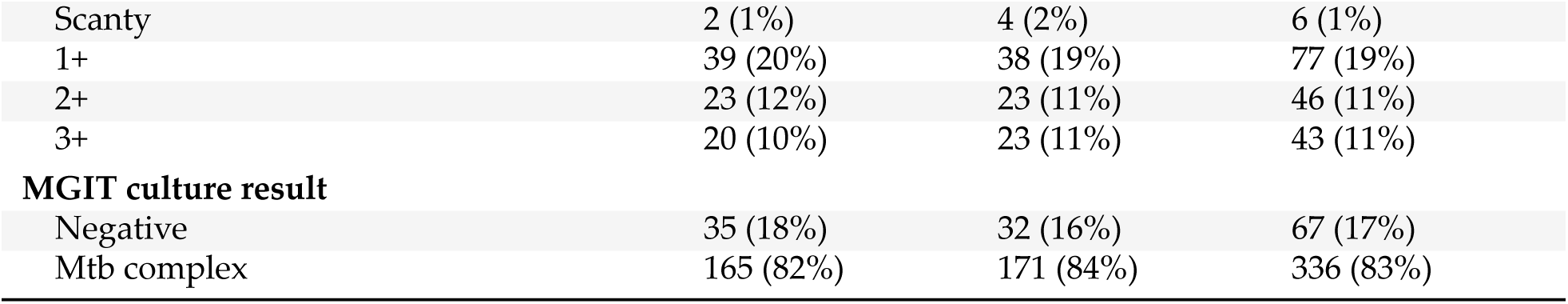
Characteristics of all randomised participants at baseline.

|  | Control Strategy | Study Strategy | Total |
| --- | --- | --- | --- |
| **Total randomised** | 200 | 203 | 403 |
| **Study site** |  |  |  |
| Port Elizabeth | 182 (91%) | 184 (91%) | 366 (91%) |
| Durban | 18 (9%) | 19 (9%) | 37 (9%) |
| **Age (years)** |  |  |  |
| Median (IQR) | 34.5 (27.0, 44.0) | 35.0 (28.0, 43.0) | 35.0 (28.0, 43.0) |
| Under 18 | 17 (8%) | 13 (6%) | 30 (7%) |
| **Sex** |  |  |  |
| Female | 85 (42%) | 85 (42%) | 170 (42%) |
| **Race** |  |  |  |
| Black | 160 (80%) | 150 (74%) | 310 (77%) |
| Mixed | 40 (20%) | 50 (25%) | 90 (22%) |
| White | 0 | 3 (1%) | 3 (1%) |
| **BMI (kg/m<sup>2</sup>)** |  |  |  |
| Median (IQR) | 19.3 (17.2, 22.4) | 19.1 (17.0, 22.0) | 19.2 (17.1, 22.2) |
| Under 18.5 | 82 (41%) | 85 (42%) | 167 (41%) |
| **HIV Status** |  |  |  |
| HIV Positive | 100 (50%) | 105 (52%) | 205 (51%) |
| CD4 Median (IQR) | 229.0 (87.0, 395.0) | 168.0 (85.0, 298.5) | 194.0 (87.0, 362.0) |
| **CD4 Category** |  |  |  |
| <200 | 38 (19%) | 50 (25%) | 88 (22%) |
| ≥200 | 43 (22%) | 38 (19%) | 81 (20%) |
| HIV Neg | 100 (50%) | 98 (48%) | 198 (49%) |
| Missing | 19 (10%) | 17 (8%) | 36 (9%) |
| **Highest level of education** |  |  |  |
| No schooling | 0 | 1 (<0.5%) | 1 (<0.5%) |
| Primary school not complete | 16 (8%) | 20 (10%) | 36 (9%) |
| Primary school complete | 11 (6%) | 12 (6%) | 23 (6%) |
| High school not complete | 126 (63%) | 125 (62%) | 251 (62%) |
| High school complete | 45 (22%) | 32 (16%) | 77 (19%) |
| Tertiary education not complete | 2 (1%) | 6 (3%) | 8 (2%) |
| Tertiary education complete | 0 | 7 (3%) | 7 (2%) |
| **Previous diagnosis of TB** |  |  |  |
| None | 98 (49%) | 92 (45%) | 190 (47%) |
| DS-TB | 99 (50%) | 105 (52%) | 204 (51%) |
| MDR-TB | 3 (2%) | 6 (3%) | 9 (2%) |
| **Cavities** |  |  |  |
| No | 58 (29%) | 55 (27%) | 113 (28%) |
| Yes | 40 (20%) | 37 (18%) | 77 (19%) |
| Missing | 102 (51%) | 111 (55%) | 213 (53%) |
| **Drug resistance profile** |  |  |  |
| FQ Susceptible and No Bedaquiline DST | 123 (62%) | 123 (61%) | 246 (61%) |
| FQ Resistant and No Bedaquiline DST | 15 (8%) | 7 (3%) | 22 (5%) |
| FQ Resistant and Bedaquiline Susceptible | 24 (12%) | 35 (17%) | 59 (15%) |
| FQ Resistant and Bedaquiline Resistant | 4 (2%) | 0 | 4 (1%) |
| No FQ DST and No Bedaquiline DST | 34 (17%) | 38 (19%) | 72 (18%) |
| **Smear grading** |  |  |  |
| Neg | 116 (58%) | 115 (57%) | 231 (57%) |

Table 1: Characteristics of all randomised participants at baseline. (*continued*)
|  | Control Strategy | Study Strategy | Total |
| --- | --- | --- | --- |
| Scanty | 2 (1%) | 4 (2%) | 6 (1%) |
| 1+ | 39 (20%) | 38 (19%) | 77 (19%) |
| 2+ | 23 (12%) | 23 (11%) | 46 (11%) |
| 3+ | 20 (10%) | 23 (11%) | 43 (11%) |
| **MGIT culture result** |  |  |  |
| Negative | 35 (18%) | 32 (16%) | 67 (17%) |
| Mtb complex | 165 (82%) | 171 (84%) | 336 (83%) |

**Table 2:**
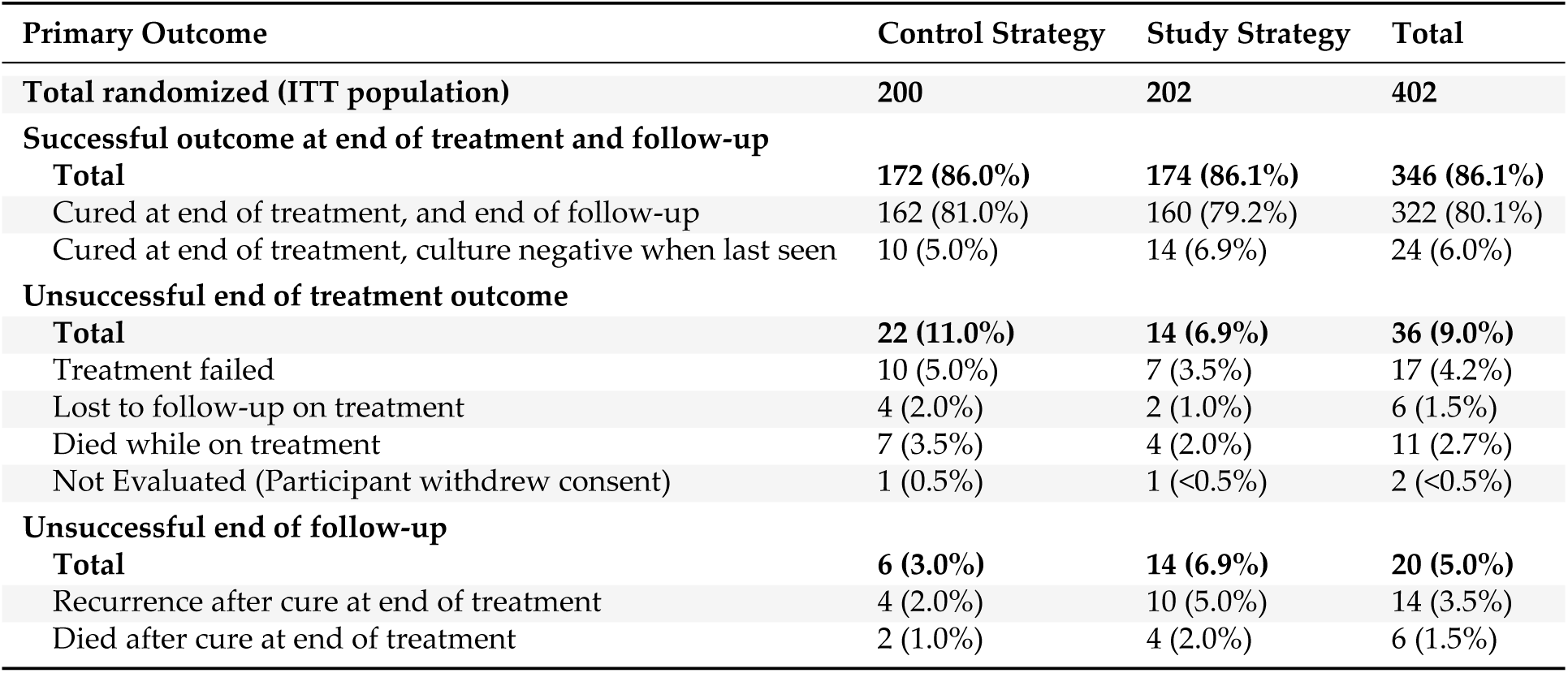
Primary Efficacy Analysis, 76 week outcome, in the intention-to-treat anlaysis population.

**Figure 1.**
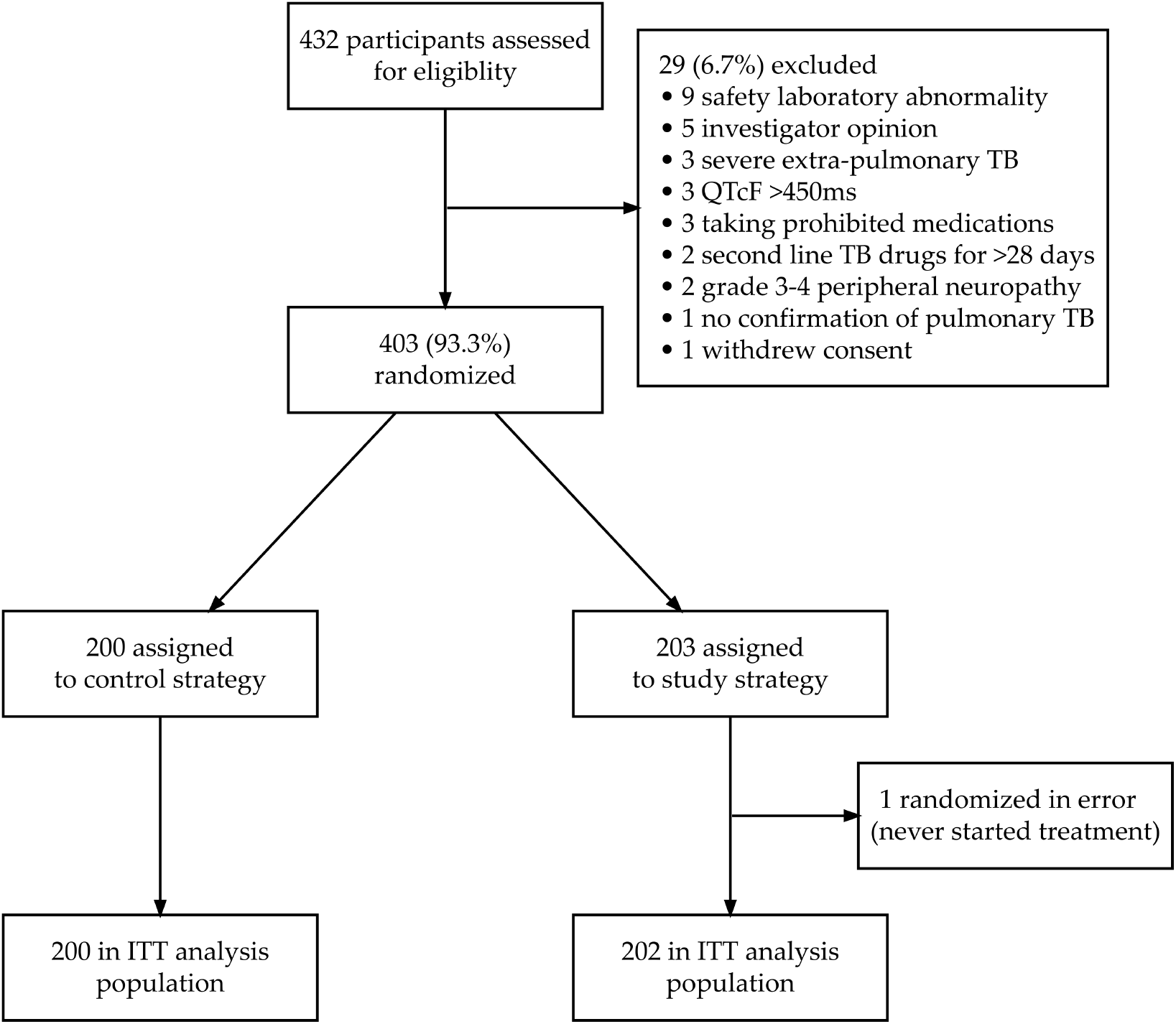
Screening, Randomization, and Follow-up. ITT indicates intention-to-treat.

**Figure 2.**
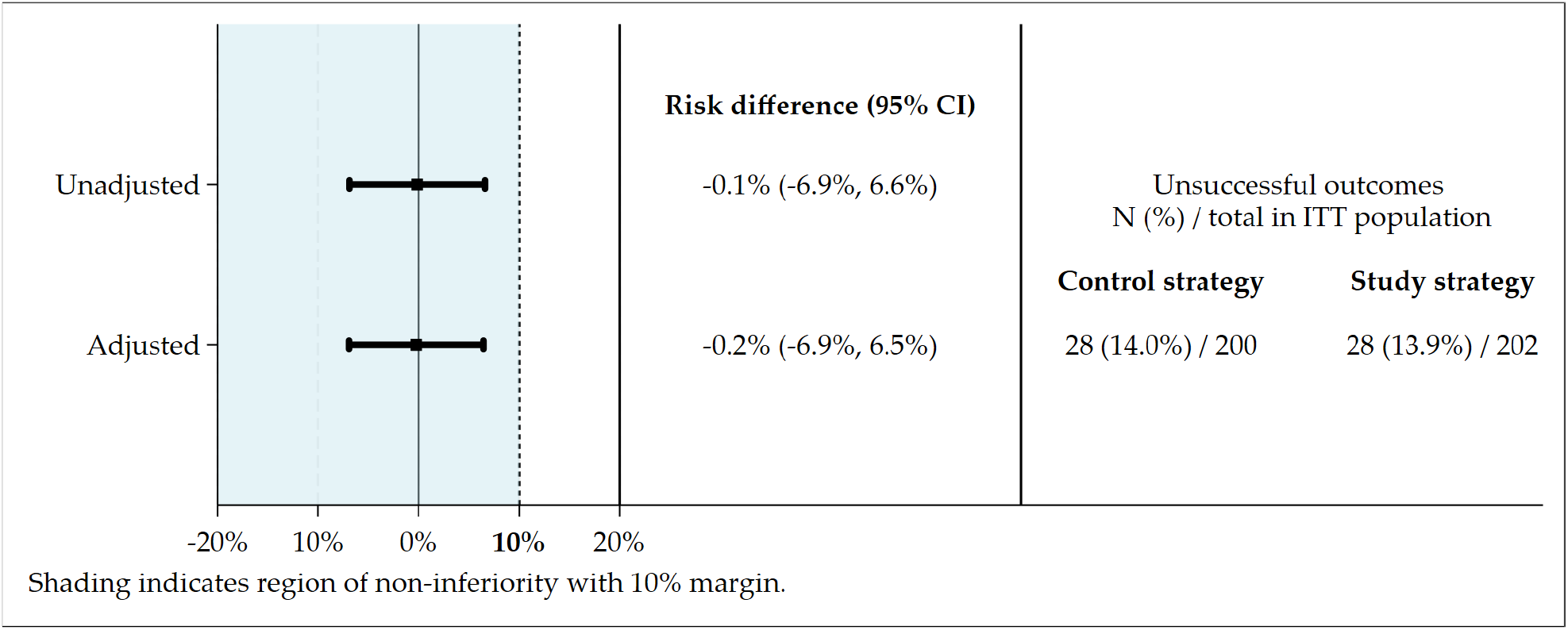
Primary efficacy analyses. Figure shows the results of the primary efficacy analysis in the intention-to-treat analysis population. The noninferiority margin of 10 percentage points is designated by the dashed vertical line. ITT indicates intention-to-treat; CI indicates confidence interval.

Of the 403 participants, 72 (18%) were diagnosed with Xpert MTB/RIF Ultra and had no subsequent fluoroquinolone resistance testing as there was no growth in the culture or the culture was contaminated. Eighty-five (21%) participants’ isolates had fluoroquinolone resistance; phenotypic bedaquiline DST was successfully done for 63 (74%) of these isolates, and 4 (6.4%) had bedaquiline resistance before starting treatment. These participants, irrespective of treatment arm, had treatment modified to an individualised rescue regimen.

### Effectiveness

One participant was randomised in error and did not start treatment. Of the 402 in the primary analysis, 172/200 (86.0%) and 174/202 (86.1%) had successful outcomes at both the end of treatment and 76 weeks in the control and study strategies, respectively, giving an adjusted risk difference of -0.2% (95% CI -6.9% to 6.5%) demonstrating non-inferiority (p=0.0014 test for non-inferiority). There was no difference in time to stable negative culture conversion by strategy nor fluoroquinolone susceptibility (See Figure S3.24 and S3.25 in the supplement). There was no evidence that treatment effects varied according to age, sex, HIV status, sputum-smear status, cavitation on chest x-ray, or fluoroquinolone resistance (For full analysis see Supplementary Appendix, section 3.12), although our study was not powered to rule out differences across any subgroups.

### Resistance acquisition

Seventeen participants experienced treatment failure (failure to culture convert or culture reversion): 10 participants (5.0%) in the control strategy and seven participants (3.5%) in the study strategy. Five participants in the control strategy and three in the study strategy had resistance to bedaquiline at the time of failure (Table S3.23 and S3.24). Fourteen participants experienced a recurrence: 4 (2.0%) from the control strategy and 10 (5.0%) from the study strategy. Two participants in the study strategy exhibited resistance to bedaquiline at the time of relapse. However, as this was a pragmatic study employing standard laboratory testing, resistance data for some participants was missing.

### Safety

Twenty participants died during treatment and follow-up, with 10 (5.0%) in each group. A total of 74 participants (37.0%) in the control group and 63 participants (31.2%) in the study group experienced severe (grade 3-5) adverse events (AEs) during treatment (primary safety outcome). There was no evidence of differences between groups in any safety outcomes, as all 95% CIs of risk differences included 0% (Table 3).

**Table 3:**
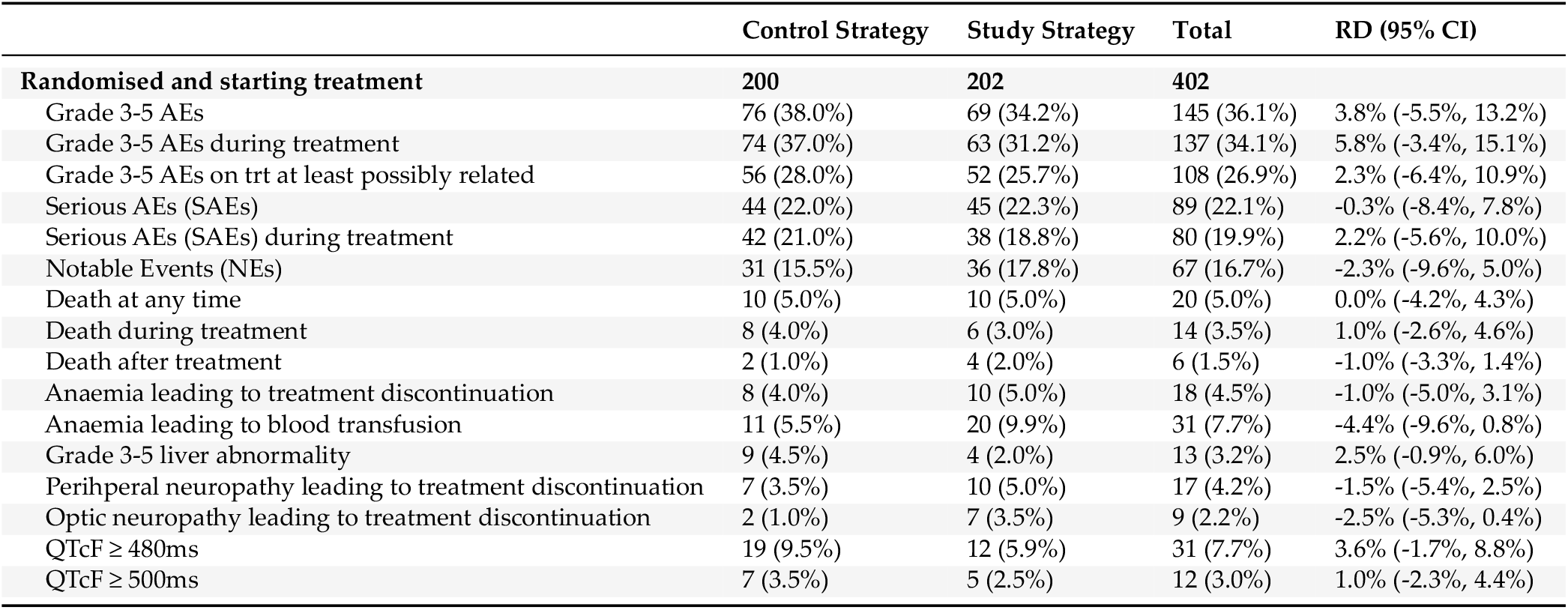
Safety summary. The safety analysis population included all the participants who had undergone randomization and received at least one dose of the assigned treatment. AE indicates Adverse Event; RD indicates Risk Difference; QTcF indicates Fridericia-corrected QT interval.

Fifty-six (28.0%) and fifty-two (25.7%) participants in the control and study strategies, respectively, experienced severe AEs that were classified as at least possibly related to the study drugs. The most common severe AE was anaemia, attributed to linezolid, which occurred in 29 (14.5%) participants in the control strategy and 33 (16.3%) in the study strategy (see Table S3.17). Most cases of anaemia occurred within the first eight weeks of treatment (refer to Figure S3.17). Anaemia was managed by temporarily interrupting linezolid treatment for a few days and transfusing packed red cells. A transfusion was administered to 11/200 (5.5%) and 20/202 (9.9%) participants in the control and study strategies. Other manifestations of myelosuppression, such as thrombocytopenia and neutropenia, were rare. Peripheral neuropathy, leading to the permanent discontinuation of linezolid, occurred in seven (3.5%) participants in the control strategy and ten (5.0%) in the study strategy. Optic neuritis was observed in nine (2.2%) participants, with seven (3.5%) in the study strategy. Nine (4.5%) severe liver-related AEs were in the control strategy compared to four (2.0%) in the study strategy.

Asymptomatic QTcF prolongation >500 ms occurred in 7 (3.5%) participants in the control strategy and 5 (2.5%) participants in the study strategy.

One child on the control strategy developed neuropsychiatric adverse reactions, which were attributed to delamanid.

In a combined efficacy-safety outcome, 115 (57.5%) and 123 (60.9%) participants completed the study with a favourable outcome and without any adverse events of grade 3 or greater in the control and study strategies, respectively (Table S3.27, online supplement).

### Treatment interruptions and extensions

Of the 202 participants who began the study strategy, 8/202 (4%) did not complete 24 weeks of treatment due to death or being lost to follow-up; 154/194 (79%) participants completed 24 weeks without interruptions of any drug, including linezolid. Twenty (10%) participants discontinued linezolid before reaching 24 weeks, with a median discontinuation time of 16 weeks (IQR 8-20). Additionally, 23 participants experienced at least one physician-directed treatment interruption of linezolid for at least 3 days, of which three discontinued linezolid early. Three participants (1.5%) in the study strategy had their treatment extended beyond 24 weeks due to late culture conversion.

### Special populations

An ultrasound confirmed a live intrauterine pregnancy, followed by a formal anomaly scan at 16 weeks. Post-delivery, the infant was examined for anomalies and signs of TB. Of the nine pregnant women enrolled, one was in the first trimester, six were in the second trimester, and two were in the third trimester (See Supplementary Appendix Table S3.18). Additionally, one woman became pregnant during the trial. All pregnancies resulted in singleton live births with one premature delivery. One woman in the study strategy experienced a recurrence of TB disease (See Supplementary Appendix Table S3.19). Two severe adverse events occurred in this group that were not drug-related (See Supplementary Appendix 3.18).

Thirty participants aged 8 to 17 years with confirmed RR-TB were enrolled, comprising 17 in the control group and 13 in the study strategy group. Among them, 7 (24%) had fluoroquinolone resistance (Table 3.28, supplementary appendix). An adolescent participant was identified as having resistance to bedaquiline and clofazimine at baseline and was subsequently switched to an individualised rescue regimen. There were five adverse events related to linezolid, three of which were graded 3 or 4 (two cases of anemia, one case of peripheral neuropathy, and two cases of optic neuritis, Table S3.31). Delamanid was discontinued in one participant due to neuropsychiatric adverse reactions. All those under 18 years of age had a successful effectiveness outcome at the end of treatment and follow-up.

## Discussion

The BEAT Tuberculosis strategy using bedaquiline, delamanid, and linezolid with levofloxacin and/or clofazimine for 6 months was non-inferior to the standard of care strategy in South Africa for RR-TB. Our findings are consistent with those from other trials of shorter regimens containing bedaquiline, delamanid or pretomanid, and linezolid. (4, 6, 11) - including a single-arm study where bedaquiline, delamanid, linezolid, and clofazimine were given for fluoroquinolone-resistant TB.(12)

The results of this trial have already informed WHO guidelines for the treatment of drug-resistant TB, with this 4-5-drug regimen included as one of the recommended regimens in the July 2024 update.(13)

The outcomes of our study using a delamanid-based regimen were comparable to those of a pretomanid-based regimen combined with bedaquiline and linezolid. Although they belong to the same class, there has been no direct comparison between delamanid and pretomanid in clinical research studies, and preclinical studies suggest each could replace the other in a regimen(14). Certain specific populations that, at this stage, cannot access pretomanid include children under 14 years and pregnant or lactating women. Our findings provide evidence for an effective regimen for children under 14 and pregnant women, offering more therapeutic options for all individuals with RR-TB. BEAT Tuberculosis was designed as a strategic trial to treat most individuals with RR-TB. It was not intended to evaluate the contribution of any single drug to the regimen.

The main strength of this trial is that it included a randomised comparison to a well-established and standardised control.(15). The two studies supporting the evidence for the BPaL regimen (NIX(3) and Zenix(4)) did not have an internal comparison group. In contrast, other recent RR-TB trials have included randomised internal control groups that varied between sites, such as TB-PRACTECAL, and during the trial’s conduct, as in STREAM Stage 2.(16) In addition, our study was designed as a single pragmatic strategy trial to include participants representative of those in the South African National TB programme. Other strengths included the high screening success rate, which more accurately represented the population requiring RR-TB treatment.

The drug causing the most adverse events in the study regimen was linezolid, but most participants could tolerate a dose of 600mg for 6 months. Short interruptions, usually for anaemia, could be managed at the site level. Regarding safety, it should be noted that only alanine transaminase was assessed routinely (every month) among hepatotoxicity evaluations, and a full liver function panel was performed only when ALT exceeded 3 times the upper limit of normal. This approach may underestimate hepatic events compared to studies where the entire panel is conducted during each hepatotoxicity evaluation encounter.

Bedaquiline resistance is becoming increasingly challenging in the treatment of MDR-TB. In this study, only fluoroquinolone-resistant isolates underwent phenotypic testing for bedaquiline resistance, the standard practice. The acquisition of bedaquiline resistance was more common in participants with RR-TB who also had resistance to fluoroquinolones (pre-extensively drug-resistant TB). Bedaquiline’s long half-life sets it apart from the other drugs in the regimen. Therefore, any treatment interruptions due to adherence challenges would result in effective monotherapy at

The BEAT Tuberculosis programme followed a pragmatic approach similar to that of the South African National TB Programme (NTP) regarding adherence support. Clinic-based Directly Observed Therapy (DOT) is not considered the standard of care. Individuals with risk factors for poor adherence were eligible for enrollment in BEAT Tuberculosis, unlike many other TB clinical trials. However, the adherence measures implemented in this trial are more stringent than the standard of care, encompassing pill counts and return verifications.

This trial had several limitations. It was conducted in only one country, South Africa, which limits the strategies’ generalizability to other regions with different resistance patterns and HIV prevalence. This strategy may require examination in other settings and against other control regimens to assess its broader generalizability and importance. In addition, treatment arms were not blinded, as the number of medications and the duration of the two regimens made this unfeasible.

An additional limitation was missing data. This was a pragmatic trial conducted within the South African NTP, in which we adhered to the diagnostic pathways outlined by the South African NTP. The diagnosis of TB was primarily established using the Xpert system, followed by DST, which was conducted using line-probe assays, which did not yield a result in 20% of cases. If all follow-up cultures returned negative results, it would be impossible to determine drug susceptibility. In the case of unknown FQ resistance, participants assigned to the study strategy continued treatment with all five drugs, while those in the control group received the standard regimen. Additionally, bedaquiline susceptibility testing was carried out only when fluoroquinolone resistance was confirmed. The study also occurred during the COVID-19 pandemic and the subsequent lockdowns. Due to disruptions in the healthcare system and the relocation of the research site, chest radiographs were not always performed. Having a chest radiograph that aligned with the diagnosis of TB before starting treatment was not included as a criterion for participation. Nevertheless, follow-up was generally good with few participants lost to follow-up on treatment or after treatment among those with end of treatment cure.

## Data Availability

All data produced in the present study are available upon reasonable request to the authors

## Conclusion

The 6-month study strategy regimen demonstrated efficacy and safety comparable to the South African standard-of-care RR/MDR-TB treatment.

This trial further expands treatment options for RR-TB by providing evidence to support a six-month strategy that offers a treatment option for both fluoroquinolone-susceptible and fluoroquinolone-resistant RR-TB and applicable to a patient population that includes children and pregnant and breastfeeding women.

## Acknowledgements

We thank the South African National TB Programme and the Eastern Cape and KwaZulu Provincial Departments of Health for donating the medications and laboratory support. We are grateful to the Independent Data Monitoring Committee members: Prof. Robert Horsburgh [chair], Daniel Grint, and Mohammed Rassool. This study was funded by USAID Cooperative Agreement No. 72067418CA00006).

**Tables and Figures for BEAT Tuberculosis: a randomized controlled trial of a short strategy for rifampicin-resistant tuberculosis**

## References

1. Global Tuberculosis Report 2024.; 2024.

2. WHO consolidated guidelines on drug resistant tuberculosis treatment. Geneva: World Health Organization; 2019.

3. Conradie F, Diacon AH, Ngubane N, Howell P, Everitt D, Crook AM, et al. Treatment of highly drug-resistant pulmonary tuberculosis. N Engl J Med. 2020;382(10):893–902.

4. Conradie F, Bagdasaryan TR, Borisov S, Howell P, Mikiashvili L, Ngubane N, et al. Bedaquiline-Pretomanid-Linezolid Regimens for Drug-Resistant Tuberculosis. N Engl J Med. 2022;387(9):810–23.

5. Nyang’wa BT, Berry C, Kazounis E, Motta I, Parpieva N, Tigay Z, et al. Short oral regimens for pulmonary rifampicin-resistant tuberculosis (TB-PRACTECAL): an open-label, randomised, controlled, phase 2B-3, multi-arm, multicentre, non-inferiority trial. Lancet Respir Med. 2024;12(2):117–28.

6. Guglielmetti L, Khan U, Velasquez GE, Gouillou M, Abubakirov A, Baudin E, et al. Oral Regimens for Rifampin-Resistant, Fluoroquinolone-Susceptible Tuberculosis. N Engl J Med. 2025;392(5):468–82.

7. WHO consolidated guidelines on tuberculosis. Module 4: Treatment – tuberculosis care and support. Geneva: World Health Organization; 2022.

8. sU.S. Department of Health and Human Services NIoH, National Institute of Allergy and Infectious Diseases, Division of AIDS. Division of AIDS (DAIDS). Table for Grading the Severity of Adult and Pediatric Adverse Events 2017 [Available from: https://rsc.niaid.nih.gov/sites/default/files/daidsgradingcorrectedv21.pdf.

9. World Health Organization. Definitions and reporting framework for tuberculosis. 2013 revision, updated December 2014. Geneva: World Health Organization; 2013. https://appswhoint/iris/handle/10665/79199. 2013.

10. Mohamed K, Embleton A, Cuffe RL. Adjusting for covariates in non-inferiority studies with margins defined as risk differences. Pharmaceutical statistics. 2011;10(5):461–6.

11. Conradie F, Diacon AH, Ngubane N, Howell P, Everitt D, Crook AM, et al. Treatment of highly drug-resistant pulmonary tuberculosis. N Engl J Med. 2020;382(10):893–902.

12. Padmapriyadarsini C, Vohra V, Bhatnagar A, Solanki R, Sridhar R, Anande L, et al. Bedaquiline, Delamanid, Linezolid and Clofazimine for Treatment of Pre-extensively Drug-Resistant Tuberculosis. Clin Infect Dis. 2022;76(3):e938–46.

13. Key updates to the treatment of drug-resistant tuberculosis: rapid communication. Geneva: World Health Organization; 2024.

14. Mudde SE, Upton AM, Lenaerts A, Bax HI, De Steenwinkel JEM. Delamanid or pretomanid? A Solomonic judgement! Journal of Antimicrobial Chemotherapy. 2022;77(4):880–902.

15. Ndjeka N, Campbell JR, Meintjes G, Maartens G, Schaaf HS, Hughes J, et al. Treatment outcomes 24 months after initiating short, all-oral bedaquiline-containing or injectable-containing rifampicin-resistant tuberculosis treatment regimens in South Africa: a retrospective cohort study. Lancet Infect Dis. 2022;22(7):1042–51.

16. Goodall RL, Nunn AJ, Meredith SK, Bayissa A, Bhatnagar AK, Chiang CY, et al. Long-term efficacy and safety of two short standardised regimens for the treatment of rifampicin-resistant tuberculosis (STREAM stage 2): extended follow-up of an open-label, multicentre, randomised, non-inferiority trial. Lancet Respir Med. 2024;12(12):975–87.

