## Supplement for "BEAT Tuberculosis: a randomized controlled trial of a 6-month strategy for rifampicin-resistant tuberculosis"

.

### BEAT Tuberculosis
### (Building Evidence for Advancing new Treatment for Tuberculosis)

**An open label, randomized controlled trial to establish the efficacy and safety of a Study Strategy consisting of 6 months of bedaquiline (BDQ), delamanid (DLM), and linezolid (LNZ), with levofloxacin (LVX) and clofazimine (CFZ) compared to the current South African Standard of Care (Control Strategy) for 9 months for the treatment of rifampicin resistant tuberculosis (RR-TB).**

### Supplementary Appendix

#### May 2, 2025

### Table of contents

**1 Full list of all study contributors** 8

**2 Supplementary Methods** 9
2.1 Background 9
2.2 Study Objectives 9
2.2.1 Primary Objectives: 9
2.2.2 Secondary Objectives 9
2.3 Randomization 10
2.4 Diagnostic Triage 10
2.5 Interventions 10
2.5.1 Study strategy 10
2.5.2 Control Strategy 11
2.5.3 Drug Doses (both strategies) 11
2.6 Full Eligibility Criteria 12
2.6.1 Inclusion Criteria 12
2.6.2 Exclusion Criteria 13
2.7 Schedule of Events 14
2.7.1 On Treatment 14
2.7.2 Off Treatment 16
2.8 Bacteriological Procedures 16
2.9 Primary Efficacy Outcome Definition 17
2.9.1 End of Treatment 17
2.9.2 End of Follow-up 17

**3 Supplementary Results** 19
3.1 Screening and Randomization 19
3.2 Baseline Characteristics 22
3.3 Treatment Regimens 27
3.3.1 Treatment prior to Randomization 27
3.3.2 Initial regimen at baseline 28
3.3.3 Summary of duration of treatment (not including doses reported prior to randomization) 28
3.3.4 Summary of duration of treatment (including doses reported prior to randomization) 33
3.4 Safety: Adverse Events 38
3.5 Safety laboratory parameter summaries 42
3.6 ECG Results 46
3.7 Change in Weight 47
3.8 Pregnancy 48
3.9 Time to stable culture negative conversion 49
3.10 Primary Efficacy 50
3.10.1 Primary outcome: Combined end of treatment and end of follow-up outcome 50
3.11 Change in resistance profile during and after treatment 52
3.12 Subgroup Analyses of the Primary Efficacy and Safety Outcomes 54
3.12.1 Study Site 55
3.12.2 Gender 56
3.12.3 Pregnancy 57
3.12.4 Age at Baseline 58
3.12.5 BMI 59

S3.54 Subgroup by smear grading at baseline: Risk difference plot of the primary safety outcome. 68
S3.55 Subgroup by injectable resistance: Non-inferiority plot of the primary efficacy outcome. 69
S3.56 Subgroup by injectable resistance: Risk difference plot of the primary safety outcome. . 69
S3.57 Subgroup by fluoroquinolone resistance: Non-inferiority plot of the primary efficacy outcome. . . . . . . . . . . . . . . . . . . . . . . . . . . . . . . . . . . . . . . . 70
S3.58 Subgroup by fluoroquinolone resistance: Risk difference plot of the primary safety outcome. 70
S3.59 Subgroup by bedaquiline resistance: Non-inferiority plot of the primary efficacy outcome. 71
S3.60 Subgroup by bedaquiline resistance: Risk difference plot of the primary safety outcome. 71
S3.61 Subgroup by clofazimine resistance: Non-inferiority plot of the primary efficacy outcome. 72
S3.62 Subgroup by clofazimine resistance: Risk difference plot of the primary safety outcome. 72
S3.63 Subgroup by linezolid resistance: Non-inferiority plot of the primary efficacy outcome. 73
S3.64 Subgroup by linezolid resistance: Risk difference plot of the primary safety outcome. . 73
S3.65 Subgroup by isoniazid resistance: Non-inferiority plot of the primary efficacy outcome. 74
S3.66 Subgroup by isoniazid resistance: Risk difference plot of the primary safety outcome. . 74
S3.67 Subgroup by baseline resistance pattern: Non-inferiority plot of the primary efficacy outcome. . . . . . . . . . . . . . . . . . . . . . . . . . . . . . . . . . . . . . . . 75
S3.68 Subgroup by baseline resistance pattern: Risk difference plot of the primary safety outcome. 75
S3.69 Subgroup by ALT (U/L): Non-inferiority plot of the primary efficacy outcome. . . . . . 76
S3.70 Subgroup by ALT (U/L): Risk difference plot of the primary safety outcome. . . . . . . 76
S3.71 Subgroup by haemoglobin (g/dL): Non-inferiority plot of the primary efficacy outcome. 77
S3.72 Subgroup by haemoglobin (g/dL): Risk difference plot of the primary safety outcome. . 77
S3.73 Subgroup by platelets (x10$^9$/L): Non-inferiority plot of the primary efficacy outcome. . 78
S3.74 Subgroup by platelets (x10$^9$/L): Risk difference plot of the primary safety outcome. . . 78
S3.75 Subgroup by neutrophils (x10$^9$/L): Non-inferiority plot of the primary efficacy outcome. 79
S3.76 Subgroup by neutrophils (x10$^9$/L): Risk difference plot of the primary safety outcome. 79
S3.77 Subgroup by creatinine (mmol/L): Non-inferiority plot of the primary efficacy outcome. 80
S3.78 Subgroup by creatinine (mmol/L): Risk difference plot of the primary safety outcome. . 80
S3.79 Subgroup by albumin (g/L): Non-inferiority plot of the primary efficacy outcome. . . . 81
S3.80 Subgroup by albumin (g/L): Risk difference plot of the primary safety outcome. . . . . 81
S3.81 Subgroup by cavities on chest x-ray: Non-inferiority plot of the primary efficacy outcome. 82

S3.82 Subgroup by cavities on chest x-ray: Risk difference plot of the primary safety outcome. 82
S3.83 Subgroup by chest x-ray result: Non-inferiority plot of the primary efficacy outcome. . 83
S3.84 Subgroup by chest x-ray result: Risk difference plot of the primary safety outcome. . . 83
S3.85 Subgroup by chest x-ray consistent with TB: Non-inferiority plot of the primary efficacy outcome. . . . . . . . . . . . . . . . . . . . . . . . . . . . . . . . . . . . . . . . . 84
S3.86 Subgroup by chest x-ray consistent with TB: Risk difference plot of the primary safety outcome. . . . . . . . . . . . . . . . . . . . . . . . . . . . . . . . . . . . . . . . . 84

### 1 Full list of all study contributors

**Isango Lethemba TB Research Unit**

Ayanda Daniso, Bernadette Rance, Buntu Myemane, Hannelise Feyt, Kelvin Vaaltyn ; Lubabalo Dingana Mary Ngodwana, Maxine Leo, Natalene Gallant, Sonja Goliath, Tamsin Economou, Tasneem Noorshib, Xabisa Makeleni, Zaheer Gaida, Zoleka Nini, Judy Sparg, and Abelang Mosweu.

**CHRU, Durban South Africa**

Londiwe Luthuli, Sikhumbuzo Majola, Ella Lesego Ndlovu, Thobile Shinga, Nompumelelo Ndlovu, Zanele Dlokweni, Ravi Maharaj, Muziwandile Hezekial Ndlovu, Onke Hubela, Silindokuhle Goge, Deborah Chili, and Nompumelelo Motaung.

**Jose Pearson TB Hospital Management**

Siziwe Ntsabo and Limpho Ramangoela.

**King Dinizulu Hospital Management**

Z Dlamini and T Mabesa.

**Clinical HIV Research Unit, Helen Joseph Hospital, University of the Witwatersrand, Johannesburg, South Africa (Medical Monitors)**

Pauline Howell and Nokuphiwa Mvuna.

**National Health Laboratory Services**

Cindy Hayes and Loyiso Mnqokoyi.

**National institute of Communicable Diseases**

Farzana Ismail and Shaheed Omar.

**USAID**

YaDiul Mukadi, Viktoriya Livchits, Phyllis Pholoholo, and Cindy Dlamini.

### 2 Supplementary Methods

#### 2.1 Background

BEAT Tuberculosis is a pragmatic strategy trial for most types of RR-TB. The inclusion criteria were drafted to be as inclusive as possible for the individuals who present to South African National TB program. The first version of the protocol did not include either pregnant women or children. However, in consultation with the Trial steering committee, in April 2020 the decision was made to include these population groups.

#### 2.2 Study Objectives

The objective of this study it to compare the efficacy and safety of a Study Strategy to a Control Strategy. Both the Study and the Control Strategies will be modified once fluoroquinolone susceptibility has been established.

##### 2.2.1 Primary Objectives:

###### 2.2.1.1 Efficacy Objective

To assess whether the proportion of participants with a successful outcome at the end of treatment and at the end of follow up at 76 weeks after randomization on the Study Strategy is not inferior to that on the Control Strategy.

###### 2.2.1.2 Safety Objective

To compare the proportion of participants who experience grade 3 or greater adverse events during treatment on the Study Strategy as compared to the Control Strategy.

##### 2.2.2 Secondary Objectives

###### 2.2.2.1 Composite efficacy and safety objective

To assess if the proportion of participants with a successful composite outcome at 76 weeks after randomization on the Study Strategy is superior to the Control Strategy.

###### 2.2.2.2 Pharmacokinetics/Pharmacodynamics (PK/PD)

- Develop a population PK model of clofazimine exposure.
- Estimate individual participant drug exposures for bedaquiline, delamanid, levofloxacin, and linezolid from sparse data using existing population PK models.
- Determine PK-PD relationships of toxicity (QT prolongation and increase in ALT from baseline) using estimated individual participant drug exposures of drugs/metabolites known to cause QT prolongation (clofazimine, bedaquiline M2 metabolite, delamanid DM6705 metabolite, and levofloxacin)

- Determine PK-PD relationships of linezolid toxicity (bone marrow suppression, peripheral neuropathy, and optic neuritis)
- Determine the change in the intervention strategy of free concentrations of bedaquiline, delamanid, levofloxacin, and linezolid over the first 8 weeks of therapy in a sub-group.
- Determine PK-PD relationships of efficacy (using change in time to sputum culture positivity as the efficacy parameter) in the intervention strategy of bedaquiline, delamanid, levofloxacin, and linezolid.

##### 2.3 Randomization

Participants were randomized in a ratio of 1:1 ratio to the Study Strategy or the Control Strategy. Randomization was stratified by HIV status and sites. Separate randomization lists for each combination of strata were prepared by an independent PHRU using permuted blocks of varying sizes. Participants were randomized using a web-based randomization system; if web access was not available at the time of randomization, a manual alternative using sealed envelopes was provided.

##### 2.4 Diagnostic Triage

Figure S2.1: Diagnostic triage flowchart.

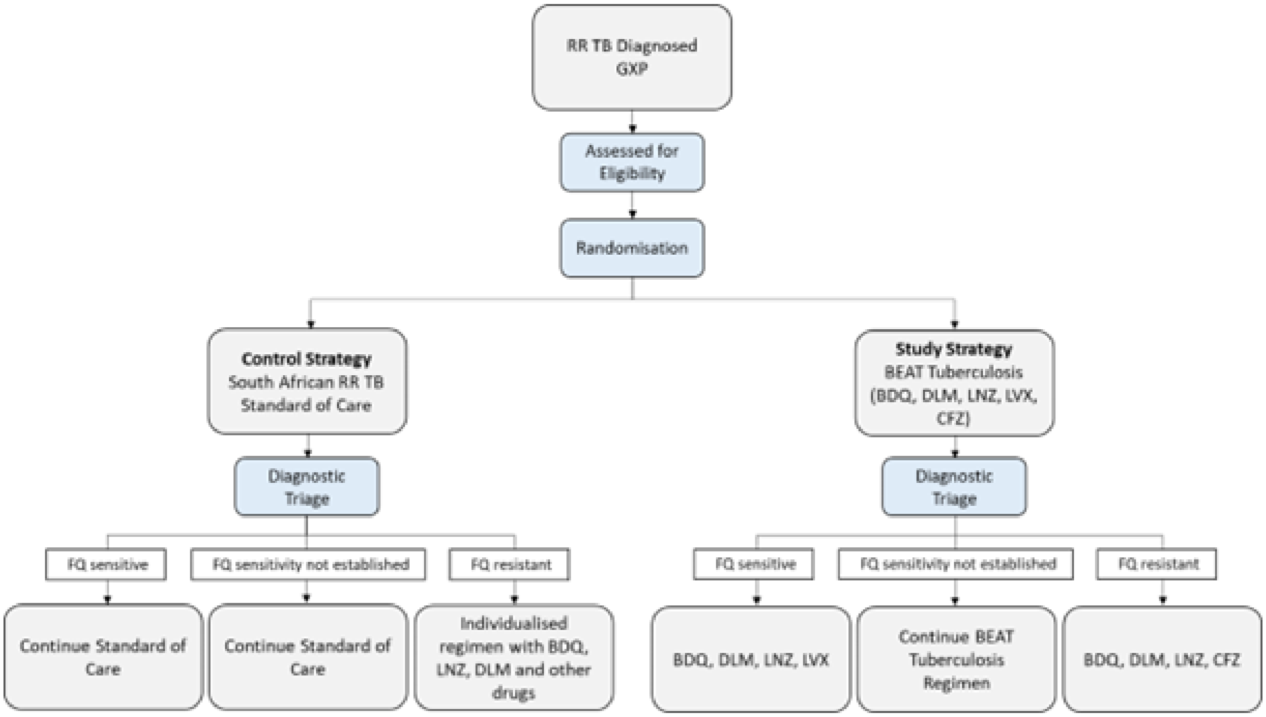

##### 2.5 Interventions

###### 2.5.1 Study strategy

The study strategy consists of bedaquiline, delamanid, linezolid, levofloxacin and clofazimine at doses shown below. All treatment in the Study Strategy is given orally for 24 weeks.

##### 2.5.2 Control Strategy

The Control Strategy is the South African Standard of care for RR-TB. This will be based on the most up to date, approved SA NTP guidance. Should this guidance change during the course of the study, the Control Strategy will also change. The site/s must follow the latest approved SA NTP guidelines as the Control Strategy.

At study start and throughout, the South African standard treatment for RR-TB was a 9-month, all-oral regimen consisting of bedaquiline (used for 6 months), linezolid (used for first 2 months), levofloxacin, ethambutol, high-dose isoniazid (for 4 months, extended to 6 months if the patient remains sputum smear positive at the end of 4 months), pyrazinamide and clofazimine, followed by treatment with levofloxacin/moxifloxacin, clofazimine, ethambutol and pyrazinamide (for 5 months). If fluoroquinolone resistance is diagnosed, then an 18-month regimen of bedaquiline, linezolid, clofazimine, terizidone and delamanid is given.

All treatment in the Control Strategy is given orally.

As per standard practice, at the time of treatment initiation, a sputum sample will be sent for ascertainment of fluoroquinolone susceptibility.

- If fluoroquinolone susceptibility is confirmed, the regimen will be continued unchanged.
- If fluoroquinolone resistance is confirmed, the regimen will be modified as per the latest NTP guidelines.
- If the fluoroquinolone resistance is known prior to treatment initiation, an individualized regimen will be given.
- If fluoroquinolone susceptibility is confirmed by genotype, the regimen will remain unchanged. Additionally, the genotypic result will be confirmed by phenotype as per the national testing guidelines and treatment modified as required, if there is a discrepancy. Discuss with the PI or Clinical Medical Monitors.

##### 2.5.3 Drug Doses (both strategies)

- Bedaquiline (BDQ)
  - *16-29.9kg*:
    - Participants ≥6 years old: 200mg daily for two weeks followed by 100mg three times weekly for weeks 3 to 24
    - Participants <6 years old: lower dose from 15-29.9kg, higher dose from ≥29 kg).
  - *≥30kg*: 400mg once daily for 14 days followed by 200mg three times weekly for weeks 3 to 24.
- Delamanid (DLD)
  - *16-23kg*: 25mg twice daily for 24 weeks.
  - *23.1-33.9kg*: 50mg twice daily for 24 weeks.
  - *≥34kg*: 100mg twice daily for 8 weeks followed by 200 mg daily for 16 weeks.
- Linezolid (LNZ)
  - *16-23kg*: 180-210mg (crush 1 tab and mix in 10ml water, administer 3-3.5ml. Discard rest).
  - *23.1-29.9kg*: 300mg daily.
  - *30-33.9kg*: 450mg daily.

    - *≥34kg*: 600mg daily.

- Levofloxacin (LVX)
    - *16-23kg*: 375-500mg daily.
    - *23.1-33.9kg*: 500mg daily.
    - *34-50kg*: 750mg daily.
    - *>50kg*: 1000mg daily.
- Clofazimine (CFZ)
    - *16-23kg*: 100mg three times a week or 50mg daily.
    - *>23kg*: 100mg daily.
- Isoniazid (INH)
    - *16-23kg*: 300mg daily.
    - *23.1-50kg*: 400mg daily.
    - *>50kg*: 600mg daily.
- Ethambutol (E)
    - *16-23kg*: 400mg daily.
    - *23.1-29.9kg*: 600mg daily.
    - *30-50kg:* 800mg daily.
    - *>50kg*: 1200mg daily.
- Pyrazinamide (Z)
    - *16-23kg*: 750mg daily.
    - *23.1-29.9kg*: 1000mg daily.
    - *30-33.9kg:* 1250mg daily.
    - *34-50mg*: 1500mg daily.
    - *>50kg*: 2000mg daily.

#### 2.6 Full Eligibility Criteria

##### 2.6.1 Inclusion Criteria

Potential participants are required to meet all the following inclusion criteria during the screening period to be randomized:

1. Willing and able to give informed consent to be enrolled in the research study prior to any study related procedures (signed or witnessed consent if the participant is unable to read and understand the informed consent document; signed or witnessed consent from a child's biological parent, legal guardian or primary caregiver) and if the participant is a child (6-17 years) is willing to sign assent.
2. Willing and able to adhere to the complete follow-up schedule and to study procedures.
3. Male or female, aged 6 years or older, including breastfeeding and/or pregnant women.
4. Weigh more than or equal to 16kg.
5. Participants above the age of 12 years, must have confirmed pulmonary TB with initial laboratory result of resistance to at least rifampicin as confirmed by genotypic or phenotypic susceptibility testing in the last three months.

6. Willing to use effective contraception for females of childbearing potential if sexually active; must be willing to use either an intrauterine contraceptive device or a hormonal method for the duration of the treatment regimen and for three months thereafter.
7. Willing to have an HIV test, and if positive, is willing to be treated with appropriate antiretroviral therapy.
8. Participants between the ages of 6 – 12 years, must have either confirmed pulmonary RR-TB or probable pulmonary RR-TB and a decision has been made by the referring clinician or investigator to treat the child for RR-TB
9. Participants who are pregnant, should have an ultrasound done to confirm a viable intrauterine pregnancy prior to enrolment.

##### 2.6.2 Exclusion Criteria

Potential participants will be excluded from participation if they meet any of the following criteria during the screening period:

1. Had taken more than 28 days, but less than 24 weeks, of second line TB drugs including BDQ, LNZ, CFZ, fluoroquinolones or DLM. Please note: Participants with prior successfully treated episodes of DR TB are permitted to enroll.
2. *For data purposes exclusion criterion no.2 has been omitted.*
3. Has complicated or severe extra-pulmonary manifestations of TB, including osteo-articular, pericardial and central nervous system infection as per investigator's opinion.
4. Is unable to take oral medication.
5. Is taking any prohibited medications (see section 5)
6. Has a known allergy or hypersensitivity to any of the medicines in the regimensIs currently taking part in another clinical trial of any medicinal product.
7. Has a QTcF interval of > 480 ms. Please note: If the QTcF interval is > 480 ms, it may be repeated if participant has reversible contributory factors, i.e. low potassium or to allow washout of previous QT prolonging drugs.
8. Has clinically significant ECG abnormality in the opinion of the site investigator within 60 days prior to entry, including but not limited to second or third degree atrioventricular (AV) block or clinically important arrhythmia.
9. Participants with the following laboratory abnormality at screening. Please note: These investigations may be repeated if abnormal, provided the results are available within the screening period.

    a) Haemoglobin level of < 8.0 g/dL
    b) Platelet count < 75,000/mm3
    c) Absolute neutrophil count (ANC) < 1000/ mm3
    d) An estimated creatinine clearance (CrCl) less than 30 mL/min as calculated by the National Health Laboratory Service (NHLS) equation.
    e) Alanine aminotransferase (ALT) ≥3 x ULN
    f) Total bilirubin grade 3 or greater (>2.0 x ULN, or >1.50 x ULN when accompanied by any increase in other liver function test)
    g) Serum potassium less than 3.2 mmol/l

10. Peripheral neuropathy of grade 3 or 4 using the Division of AIDS (DAIDS) Table for Grading the Severity of Adult and Pediatric Adverse Events
11. If in the investigator's opinion, the participant is unable to commit to study related procedures or it is unsafe for the participant to take part in the study.

#### 2.7 Schedule of Events

##### 2.7.1 On Treatment

Figure S2.2: Schedule of events: On Treatment.

| **Window period** | N/A | N/A | +/- 3 days | | | | | +/- 14 days | | | | | | | | | | | | | | |
|---|---|---|---|---|---|---|---|---|---|---|---|---|---|---|---|---|---|---|---|---|---|---|
| **Evaluation** | **Screening** | **Randomizatio** | **Week 2** | **Week 4** | **Week 8** | **Week 12** | **Week 16** | **Week 20** | **Week 24** | **Week 28** | **Week 32** | **Week 36** | **Week 40** | **Week 44** | **Week 48** | **Week 52** | **Week 56** | **Week 60** | **Week 64** | **Week 68** | **Week 72** | **Week 76** |
| Signed informed consent | X | | | | | | | | | | | | | | | | | | | | | |
| Demography | X | | | | | | | | | | | | | | | | | | | | | |
| Medical & Treatment History | X | X | | | | | | | | | | | | | | | | | | | | |
| Concomitant medication | X | X | X | X | X | X | X | X | X | X | X | X | X | X | X | X | X | X | X | X | X | X |
| HIV status[1] | X | | | | | | | | | As clinically indicated | | | | | | | | | | | | |
| Inclusion & exclusion criteria | X | X | | | | | | | | | | | | | | | | | | | | |
| Signs/Symptoms & Diagnoses | X | X | X | X | X | X | X | X | X | X | X | X | X | X | X | X | X | X | X | X | X | X |
| Trial medications dispensing | | X | X | X | X | X | X | X | X | X | X | X | X | X | X | X | X | X | X | X | X | X |
| Adverse events | | X | X | X | X | X | X | X | X | X | X | X | X | X | X | X | X | X | X | X | X | X |
| Physical Examination | X | X | X | X | X | X | X | X | X | X | X | X | X | X | X | X | X | X | X | X | X | X |
| Ophthalmic Exam[2] | X | | X | X | X | X | X | X | X | X | X | X | X | X | X | X | | | | | | |
| Haematology[3] | X | | X | X | X | X | X | X | X | X | | | | | | | | | | | | |
| Full Liver Function Tests[4] | X | | | As clinically indicated | | | | | | | | | | | | | | | | | | |
| ALT[5] | | | X | X | X | X | X | X | X | X | X | X | X | X | X | X | X | X | X | X | X | X |
| Chemistry[6] | X | | X | | X | X | As clinically indicated | | | | | | | | | | | | | | | |
| Targeted neurological examination[7] | X | | | | X | X | X | X | X | X | X | X | X | X | X | X | | | | | | |
| 12 lead ECG in triplicate[8] | X | X | X | X | X | X | X | X | X | X | X | X | X | X | X | | | | | | | |
| Triangle ECG measurement[9] | | | X | X | | X | | | X | | | | | | | | | | | | | |
| Pregnancy Testing[10] | X | X | | As clinically indicated | | | | | | | | | | | | | | | | | | |
| Chest X-ray | X | | | As clinically indicated | | | | | | | | | | | | | | | | | | |
| HIV Viral Load (if HIV+)[11] | X | | | | | | | | | X | | | | | | | | | | | | |
| CD4 count (if HIV+)[12] | X | | | | | | | | | X | | | | | | | | | | | | |
| Sputum AFB Smear & Culture In Liquid Media[13] | X | X | X | X | X | X | X | X | X | X | X | X | X | X | X | X | X | X | X | X | X | X |
| Drug Susceptibility Testing (DST) | X | | | | | X | X | X | X | X | X | X | X | X | X | X | X | X | X | X | X | X |
| Adherence Assessment | | | X | X | X | X | X | X | X | X | X | X | X | X | X | X | X | X | X | X | X | X |
| Intensive PK (30 participants)[14] | | | | X | | | | | | | | | | | | | | | | | | |
| Dried Blood Spot sample (20 participants)[15] | | | | X | | | | | | | | | | | | | | | | | | |
| Sparse PK sampling[16] | | | | X | | X | | | X | | | | | | | | | | | | | |
| Free trough drug concentration analysis[17] | | | X | X | X | | | | | | | | | | | | | | | | | |
| Total blood volumes in ml (excl. intensive PK volume and DBS sample) | **25** | **0** | **15** | **23** | **20** | **28** | **20** | **20** | **28** | **30** | **15** | **15** | **15** | **15** | **15** | **15** | **15** | **15** | **15** | **15** | **15** | **15** |

1. Using the National HIV testing algorithm. If a participant has a documented HIV positive test or is currently on ARVs, a repeat test does not have to be performed.
2. Visual Acuity using the Snellen chart. Performed at each visit when the participant is on LNZ
3. Full Blood count with Haemoglobin, White cell, platelets and differential. Performed at each visit when the participant is on LNZ. A maximum of 5ml (1 tube) of blood will be drawn per visit
4. Total protein, Albumin, Bilirubin (conjugated and unconjugated), ALT. A maximum of 5ml (1 tube) of blood will be drawn per visit
5. ALT to be done at all visits while the participant is on TB treatment. If ALT >3xULN, to do full LFT panel. A maximum of 5ml (1 tube) of blood will be drawn per visit
6. Potassium, Urea Creatinine. A maximum of 5ml (1 tube) of blood will be drawn per visit
7. Targeted neurological examination including fine touch, reflexes and vibration sense
8. Investigator to read all ECG's and act according to cardiac safety guidelines (section 8.4). In addition, ECG's at weeks 4, 12 and 24 will be centrally read by a cardiology consultant
9. Additional ECG readings using the triangle device at weeks 2, 4, 12 and 24 in participants who consent to this. These ECG readings will not be used for patient management purposes.
10. Urine BHCG; if positive for serum BHCG
11. A maximum of 5ml (1 tube) of blood will be drawn for HIV Viral Load
12. A maximum of 5ml (1 tube) of blood will be drawn for CD4 Count
13. All positive isolates from screening onwards must be stored for later whole genome sequencing
14. For participants in both treatment strategies, provided the participant is taking clofazimine at the time of intensive sampling. Pre-dose, 2, 4, 6, 8, 10, and 24 hours post-dose. In these 30 intensively sample participants, ECGs will be done in triplicate at each PK sampling time point. A maximum of 30ml (6 tubes) of blood will be drawn in this PK visit. Perform on BDQ administration days. Participants need to sign a separate informed consent in order to take part in this
15. Additional samples taken for DBS validation in participants who consent to this procedure. Blood will be sampled for DBS at the same time-points as plasma from 20 participants who are scheduled for intensive sampling at 7 time-points over 24 hours (pre-dose, 2, 4, 6, 8, 10, and 24 hours post-dose). 50 microlitres of blood will be needed for this procedure
16. Pre-dose sparse sampling for all drugs for all participants (control and study strategies) will be done at weeks 4, 12, and 24. A maximum of 8ml (2 tubes) of blood will be drawn per PK visit. Perform on BDQ administration days
17. Additional PK performed in 20 participants randomized to the Study Strategy who consent to this procedure. Performed once a week for the first 8 weeks of treatment

##### 2.7.2 Off Treatment

These off-treatment visits can begin at any time point, starting 1 month from the date that the participant completed treatment. These visits will take place monthly for the first 3 months, followed by 3 monthly visits until end of study is reached. All participants must attend the end of study visit (week 76 / 19 months).

Figure S2.3: Schedule of events: Off Treatemnt.

| **Window period** | +/- 14 days | | | | | | | | |
|---|---|---|---|---|---|---|---|---|---|
| **Evaluation** | ***Month 1** | **Month 2** | **Month 3** | **Month 6** | **Month 9** | **Month 12** | **Month 15** | **Month 18** | **Month 19 / Week 76** |
| Signs/Symptoms & Diagnoses | X | X | X | X | X | X | X | X | X |
| Vital signs including weight | X | X | X | X | X | X | X | X | X |
| Adverse events | X | X | X | X | X | X | X | X | X |
| Sputum AFB Smear & Culture in Liquid Media^ | X | X | X | X | X | X | X | X | X |

* Month 1 is calculated as 4 weeks after the participant completed treatment. E.g. Participant completed treatment on 1 April 2020. Month 1 off-treatment visit will therefore be scheduled for 1 May 2020.

^ All positive isolates from screening onwards must be stored for later whole genome sequencing

#### 2.8 Bacteriological Procedures

All specimens were processed at the National Health Laboratory Service (NHLS) as routine specimens and followed the SANTP guidelines. One sputum sample was collected for liquid culture (Mycobacterial Growth Indicator Tube system, MGIT; BACTEC 960, Becton Dickinson) and smear microscopy for acid-fast bacilli at the screening visit; then every two weeks for the first 8 weeks and thereafter monthly. A GenoType MTBDRplus and MTBDRsl line-probe assay (LPA; Hain Lifescience, Nehren, Germany) was done at least once at baseline. If there was discordance in rifampicin susceptibility between the Xpert MTB/RIF Ultra, the first-line LPA or the phenotypic rifampicin DST results, the patient would receive treatment for RR-TB as per the National guidelines.

Most LPAs were conducted on a positive Mycobacterium tuberculosis culture on MGIT due to the high rate of smear negativity. In addition, during the trial, bedaquiline and linezolid phenotypic DST was introduced into the South African National TB program. This was performed when resistance was detected to fluroquinolone or at the request of the investigator. An extended drug susceptibility test (EDST) was done if culture conversion had not occurred by week 16, on relapse or at the request of the investigator. This includes testing for bedaquiline, clofazimine, ethambutol, pyrazinamide ethionamide, isoniazid, levofloxacin, linezolid, moxifloxacin (high and low dose), para-aminosalicylic acid, rifabutin and rifampicin.

##### 2.9 Primary Efficacy Outcome Definition

These are based on the WHO outcome definitions, slightly modified for use in a clinical trial and also includes post treatment follow up to 76 weeks after randomization.

A successful primary efficacy outcome requires a successful end of treatment and successful end of follow-up outconme.

###### 2.9.1 End of Treatment

A successful treatment outcome measured at the end of treatment is defined as one of the following:

- *Cured*: Adequate treatment adherence (at least 80% of dosages taken) as per protocol without evidence of failure AND last two negative sputum specimens at the end of treatment being culture negative. These specimens must be separated by at least 14 days.
- *Treatment Completed*: Adequate treatment adherence (at least 80% of dosages taken) as per protocol without evidence of failure BUT no record that two or more consecutive cultures taken at least 14 days apart are negative.

An unsuccessful treatment outcome measured at the end of treatment is defined as one of the following:

- *Treatment Failed*
  - Lack of sputum culture conversion by the end of the of treatment, OR
  - Bacteriological reversion after sputum culture conversion to negative, OR
  - If two or more anti-TB drugs are substituted due to Adverse Drug Reactions (ADRs).
- *Death*: Death during the treatment from any cause.
- *Lost to Follow-up*: A participant who missed 28 consecutive days of treatment not directed by the clinician.
- *Not Evaluated*: A participant for whom no treatment outcome is assigned (this includes cases "transferred out" to another treatment unit and whose treatment outcome is unknown).

###### 2.9.2 End of Follow-up

A successful end of follow up outcome measured at 76 weeks post treatment initiation is defined as one of the following:

- *Cured*: Culture negative at the end of follow up.
- *Culture Negative*: Culture negative when last seen, if the participant is lost before the end of follow up and provided they have a successful treatment outcome at the last study visit attended.

An unsuccessful end of follow up outcome measured at 76 weeks post treatment initiation is defined as one of the following:

- *Recurrence*
  - Two consecutive positive cultures separated by at least 14 days, OR
  - One positive culture after confirmed culture conversion with clinical signs and symptoms of TB or no improvement or worsening of radiological changes since baseline. Please note: an isolated positive smear or culture without clinical or radiographic deterioration after treatment completion provides insufficient evidence to define recurrent TB.

- *Death*: Death in follow up from any cause.
- *Lost to Follow-up*
  - With clinical signs and/or symptoms of TB when last seen, OR
  - Sputum culture positive when last seen, OR
  - Not sputum culture negative and with clinical signs and symptoms of TB when last seen.

Strain typing will be available to distinguish between relapse and reinfection. Participants who are re-infected with a different strain can still be considered a successful end of follow up outcome, provided they meet the criteria for this prior to reinfection.

### 3 Supplementary Results

#### 3.1 Screening and Randomization

Figure S3.1: Cumulative screening by month.

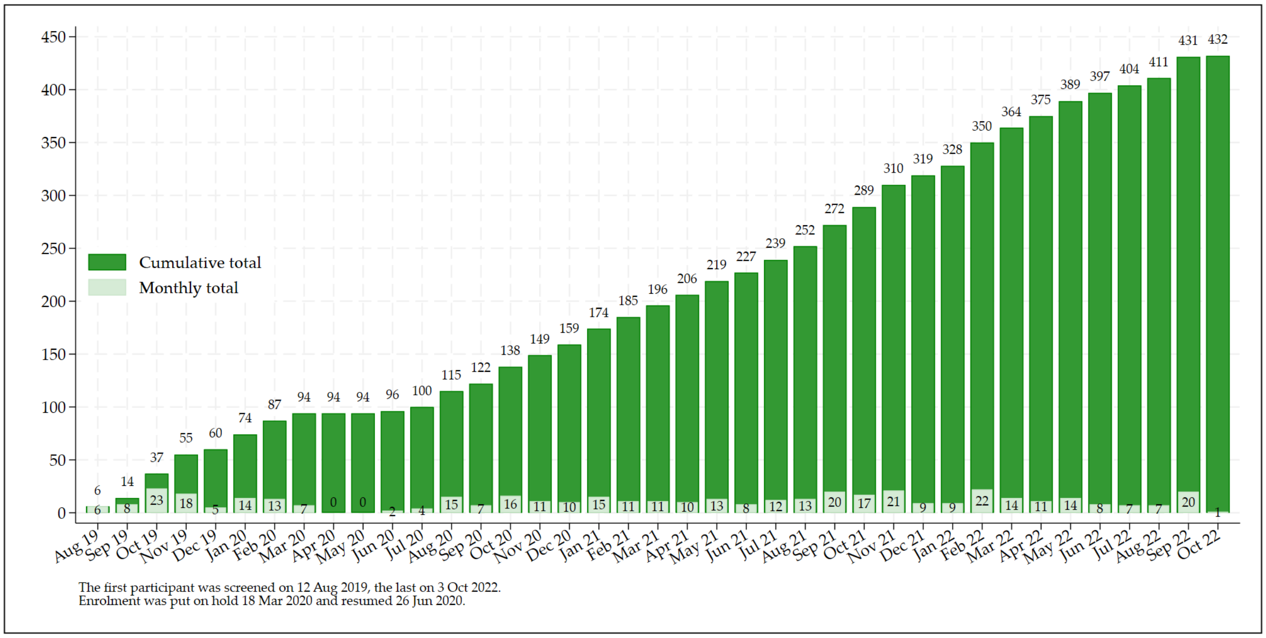

Figure S3.2: Cumulative randomization by month.

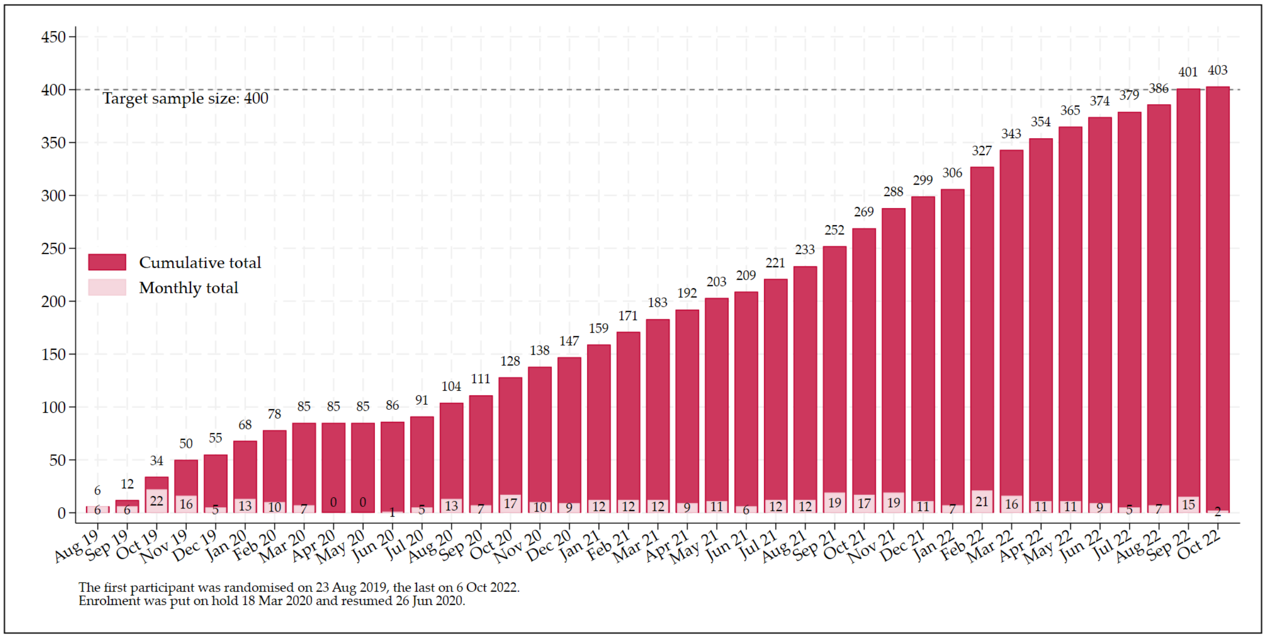

Figure S3.3: Screening and randomization over time.

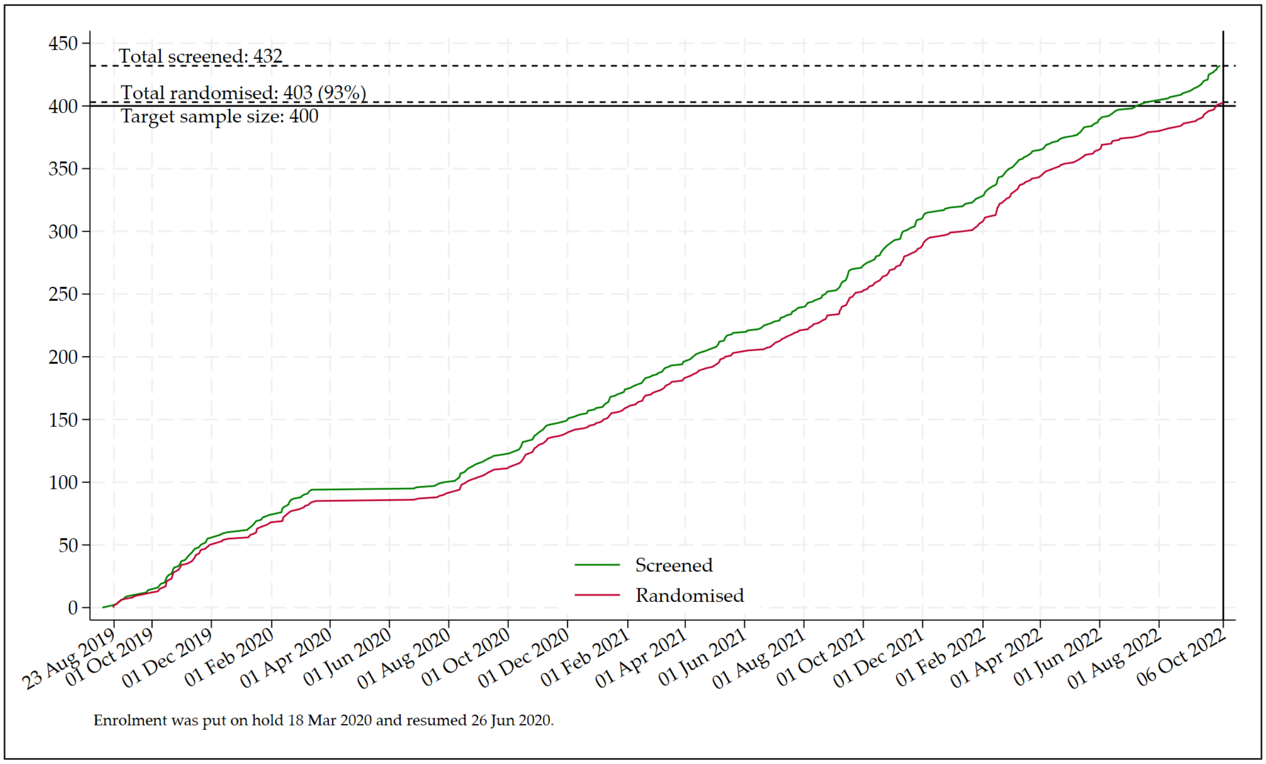

Table S3.1: Summary of reasons for not being randomized by study site

| | **Port Elizabeth** | **Durban** | **Total** |
|---|---|---|---|
| **Total screened** | **391** | **41** | **432** |
| **Total randomized** | **366 (93.6%)** | **37 (90.2%)** | **403 (93.3%)** |
| **Total not randomized** | **25 (6.4%)** | **4 (9.8%)** | **29 (6.7%)** |
| Safety lab abnormality | 8 (2.0%) | 1 (2.4%) | 9 (2.1%) |
| Investigator's opinion unable to commit to procedures | 4 (1.0%) | 1 (2.4%) | 5 (1.2%) |
| Complicated or severe extra-pulmonary TB | 1 (0.3%) | 2 (4.9%) | 3 (0.7%) |
| QTcF >450 ms | 3 (0.8%) | 0 | 3 (0.7%) |
| Taking prohibited medications | 3 (0.8%) | 0 | 3 (0.7%) |
| Peripheral neuropathy, grade 3-4 | 2 (0.5%) | 0 | 2 (0.5%) |
| Second line TB drugs for >28 days | 2 (0.5%) | 0 | 2 (0.5%) |
| No confirmation of pulmonary TB | 1 (0.3%) | 0 | 1 (0.2%) |
| Withdrew consent | 1 (0.3%) | 0 | 1 (0.2%) |

Figure S3.4: CONSORT participant flowchart.

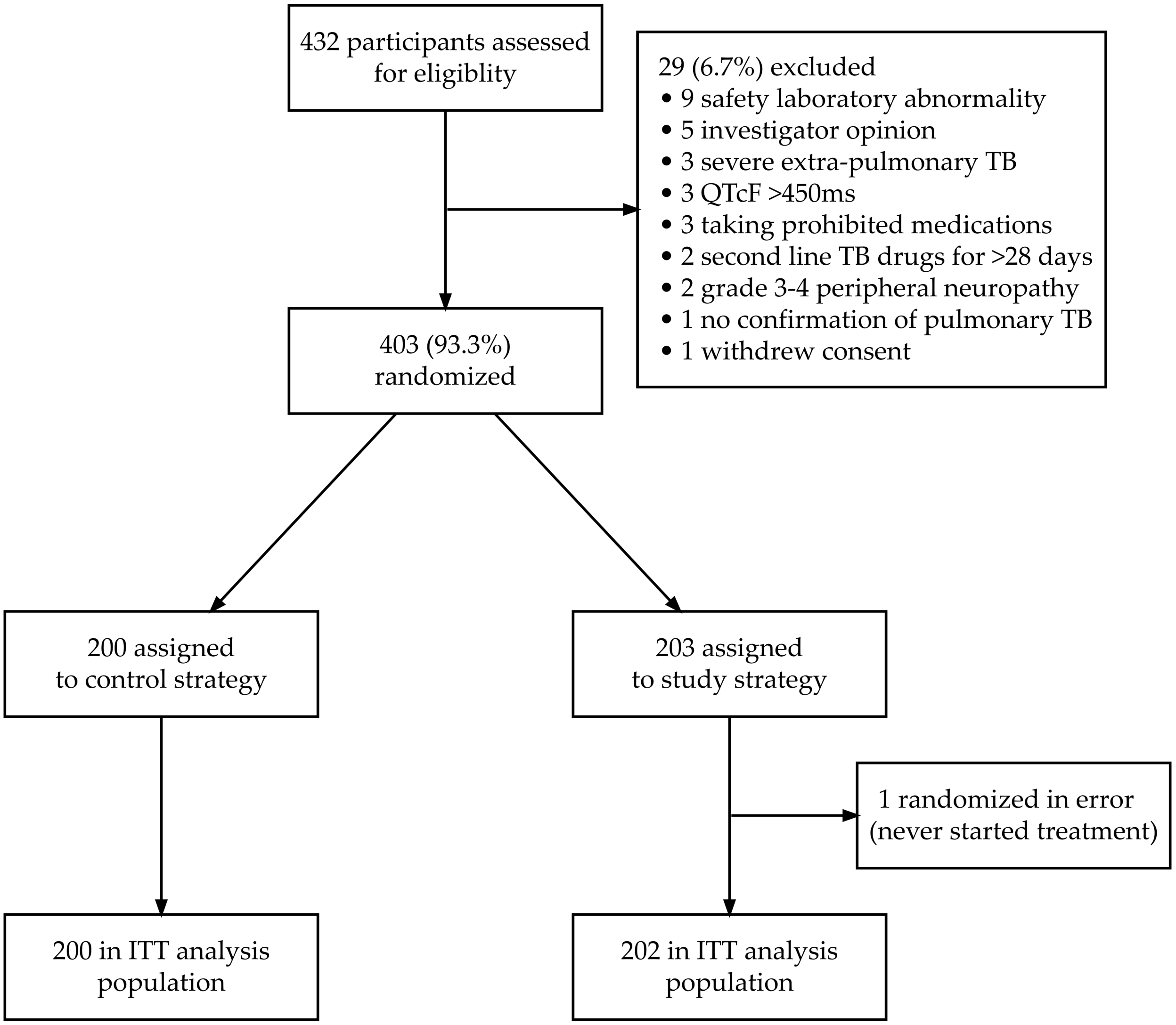

#### 3.2 Baseline Characteristics

Table S3.2: Summary of baseline characteristics by strategy.

| | Control Strategy | Study Strategy | Total |
|---|---|---|---|
| **Total randomised** | | | |
| | 200 | 203 | 403 |
| **Age (years)** | | | |
| Median (IQR) | 34.5 (27.0, 44.0) | 35.0 (28.0, 43.0) | 35.0 (28.0, 43.0) |
| Min, Max | 8.0, 66.0 | 10.0, 69.0 | 8.0, 69.0 |
| **Weight (kg)** | | | |
| Median (IQR) | 54.5 (47.9, 62.1) | 52.9 (45.5, 62.2) | 54.1 (46.9, 62.2) |
| Min, Max | 25.1, 114.7 | 28.0, 158.5 | 25.1, 158.5 |
| **BMI (kg/m²)** | | | |
| Median (IQR) | 19.3 (17.2, 22.4) | 19.1 (17.0, 22.0) | 19.2 (17.1, 22.2) |
| Min, Max | 12.7, 45.4 | 12.5, 50.6 | 12.5, 50.6 |
| Number missing | 0 | 1 | 1 |
| **QTcF at screening (ms)** | | | |
| Median (IQR) | 404.7 (392.7, 418.8) | 409.3 (393.3, 423.7) | 406.7 (392.7, 421.3) |
| Min, Max | 328.3, 499.3 | 348.3, 475.7 | 328.3, 499.3 |
| Number missing | 0 | 2 | 2 |
| **Heart Rate (HR) at screening** | | | |
| Median (IQR) | 86.2 (73.7, 99.5) | 86.3 (77.0, 98.7) | 86.3 (75.3, 99.3) |
| Min, Max | 48.7, 139.0 | 49.0, 142.3 | 48.7, 142.3 |
| Number missing | 0 | 2 | 2 |
| **Age, categorical (years)** | | | |
| 6 - <18 | 17 (8%) | 13 (6%) | 30 (7%) |
| 18 - <35 | 83 (42%) | 84 (41%) | 167 (41%) |
| 35 - <45 | 57 (28%) | 60 (30%) | 117 (29%) |
| 45 - <55 | 30 (15%) | 29 (14%) | 59 (15%) |
| ≥55 | 13 (6%) | 17 (8%) | 30 (7%) |
| **Gender** | | | |
| Male | 115 (57%) | 118 (58%) | 233 (58%) |
| Female | 85 (42%) | 85 (42%) | 170 (42%) |
| **Highest level of education** | | | |
| No schooling | 0 | 1 (<0.5%) | 1 (<0.5%) |
| Primary school not complete | 16 (8%) | 20 (10%) | 36 (9%) |
| Primary school complete | 11 (6%) | 12 (6%) | 23 (6%) |
| High school not complete | 126 (63%) | 125 (62%) | 251 (62%) |
| High school complete | 45 (22%) | 32 (16%) | 77 (19%) |
| Tertiary education not complete | 2 (1%) | 6 (3%) | 8 (2%) |
| Tertiary education complete | 0 | 7 (3%) | 7 (2%) |
| **HIV Status** | | | |
| HIV Negative | 100 (50%) | 98 (48%) | 198 (49%) |
| HIV Positive | 100 (50%) | 105 (52%) | 205 (51%) |
| **CD4 Count, categorical** | | | |
| <200 | 38 (19%) | 50 (25%) | 88 (22%) |
| ≥200 | 43 (22%) | 38 (19%) | 81 (20%) |
| HIV Neg | 100 (50%) | 98 (48%) | 198 (49%) |
| Missing | 19 (10%) | 17 (8%) | 36 (9%) |
| **Previous diagnosis of TB** | | | |
| None | 98 (49%) | 92 (45%) | 190 (47%) |

Table S3.2: Summary of baseline characteristics by strategy. *(continued)*

| | **Control Strategy** | **Study Strategy** | **Total** |
|---|---|---|---|
| DS-TB | 99 (50%) | 105 (52%) | 204 (51%) |
| MDR-TB | 3 (2%) | 6 (3%) | 9 (2%) |
| **Duration of treatment at previous TB episode** | | | |
| <6 months | 20 (10%) | 23 (11%) | 43 (11%) |
| 6-12 months | 82 (41%) | 83 (41%) | 165 (41%) |
| >12 months | 0 | 4 (2%) | 4 (1%) |
| No previous diagnosis | 98 (49%) | 92 (45%) | 190 (47%) |
| Missing | 0 | 1 (<0.5%) | 1 (<0.5%) |
| **Duration of injectable at previous TB episode** | | | |
| Not received Inj. | 85 (42%) | 105 (52%) | 190 (47%) |
| <=2 months | 4 (2%) | 1 (<0.5%) | 5 (1%) |
| >2 & <4 months | 3 (2%) | 2 (1%) | 5 (1%) |
| No previous diagnosis | 98 (49%) | 92 (45%) | 190 (47%) |
| Missing | 10 (5%) | 3 (1%) | 13 (3%) |
| **Outcome of previous TB episode** | | | |
| Treatment completed | 51 (26%) | 62 (31%) | 113 (28%) |
| Cured | 33 (16%) | 27 (13%) | 60 (15%) |
| Treatment failed | 5 (2%) | 9 (4%) | 14 (3%) |
| Lost to follow-up | 11 (6%) | 10 (5%) | 21 (5%) |
| Unknown | 0 | 2 (1%) | 2 (<0.5%) |
| No previous diagnosis | 98 (49%) | 92 (45%) | 190 (47%) |
| Missing | 2 (1%) | 1 (<0.5%) | 3 (1%) |
| **BMI, categorical (kg/m²)** | | | |
| <18.5 | 82 (41%) | 85 (42%) | 167 (41%) |
| 18.5 - <25.0 | 87 (44%) | 90 (44%) | 177 (44%) |
| ≥25.0 | 31 (16%) | 27 (13%) | 58 (14%) |
| Missing | 0 | 1 (<0.5%) | 1 (<0.5%) |
| **CD4 Count** | | | |
| Median (IQR) | 229.0 (87.0, 395.0) | 168.0 (85.0, 298.5) | 194.0 (87.0, 362.0) |
| Min, Max | 3.0, 944.0 | 15.0, 881.0 | 3.0, 944.0 |
| Number missing | 119 | 115 | 234 |

Figure S3.5: Histogram of age by gender.

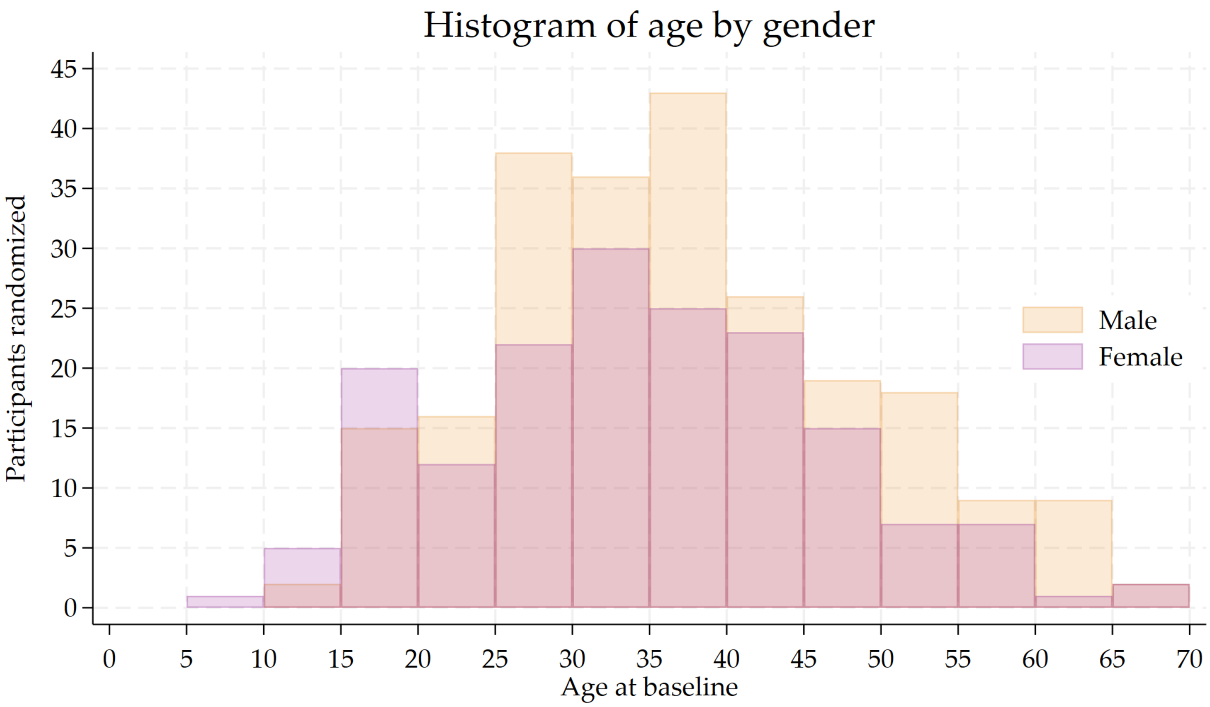

Table S3.3: Summary of baseline bacteriology by strategy.

| | Control Strategy | Study Strategy | Total |
|---|---|---|---|
| **Total randomised** | | | |
| | 200 | 203 | 403 |
| **Smear grading** | | | |
| Neg | 116 (58%) | 115 (57%) | 231 (57%) |
| Scanty | 2 (1%) | 4 (2%) | 6 (1%) |
| 1+ | 39 (20%) | 38 (19%) | 77 (19%) |
| 2+ | 23 (12%) | 23 (11%) | 46 (11%) |
| 3+ | 20 (10%) | 23 (11%) | 43 (11%) |
| **MGIT culture result** | | | |
| Negative | 35 (18%) | 32 (16%) | 67 (17%) |
| Mtb complex | 165 (82%) | 171 (84%) | 336 (83%) |
| **Rifampicin Resistance** | | | |
| Resistant | 200 (100%) | 203 (100%) | 403 (100%) |
| **Isoniazid Resistance** | | | |
| Sensitive | 52 (26%) | 62 (31%) | 114 (28%) |
| Resistant | 105 (52%) | 102 (50%) | 207 (51%) |
| Indeterminate | 2 (1%) | 0 | 2 (<0.5%) |
| Missing | 41 (20%) | 39 (19%) | 80 (20%) |
| **Fluoroquinolone Resistance** | | | |
| Sensitive | 123 (62%) | 123 (61%) | 246 (61%) |
| Resistant | 43 (22%) | 42 (21%) | 85 (21%) |
| Indeterminate | 3 (2%) | 3 (1%) | 6 (1%) |
| Missing | 31 (16%) | 35 (17%) | 66 (16%) |
| **Linezolid Resistance** | | | |
| Sensitive | 28 (14%) | 35 (17%) | 63 (16%) |
| Resistant | 1 (0%) | 1 (<0.5%) | 2 (<0.5%) |

| | | | |
|---|---|---|---|
| Missing | 171 (86%) | 167 (82%) | 338 (84%) |
| **Bedaquiline Resistance** | | | |
| Sensitive | 24 (12%) | 35 (17%) | 59 (15%) |
| Resistant | 4 (2%) | 0 | 4 (1%) |
| Missing | 172 (86%) | 168 (83%) | 340 (84%) |
| **Clofazimine Resistance** | | | |
| Sensitive | 13 (6%) | 23 (11%) | 36 (9%) |
| Resistant | 3 (2%) | 0 | 3 (1%) |
| Missing | 184 (92%) | 180 (89%) | 364 (90%) |
| **Injectables Resistance** | | | |
| Sensitive | 101 (50%) | 92 (45%) | 193 (48%) |
| Resistant | 44 (22%) | 51 (25%) | 95 (24%) |
| Indeterminate | 5 (2%) | 10 (5%) | 15 (4%) |
| Missing | 50 (25%) | 50 (25%) | 100 (25%) |

Table S3.4: Summary of baseline safety laboratory parameters by strategy.

| | Control Strategy | Study Strategy | Total |
|---|---|---|---|
| **Haemoglobin (g/dL)** | | | |
| Median (IQR) | 11.6 (10.3, 13.1) | 11.6 (10.4, 12.7) | 11.6 (10.4, 12.9) |
| Min, Max | 8.4, 15.9 | 8.0, 15.8 | 8.0, 15.9 |
| Number missing | 1 | 1 | 2 |
| **White blood cell (x10$^9$/L** | | | |
| Median (IQR) | 7.6 (5.5, 10.1) | 8.1 (5.8, 10.9) | 7.9 (5.6, 10.5) |
| Min, Max | 2.2, 22.2 | 2.3, 32.0 | 2.2, 32.0 |
| Number missing | 1 | 1 | 2 |
| **Platelets (x10$^9$/L)** | | | |
| Median (IQR) | 381.5 (297.0, 487.0) | 399.5 (299.0, 502.0) | 387.5 (298.0, 494.5) |
| Min, Max | 111.0, 942.0 | 85.0, 980.0 | 85.0, 980.0 |
| Number missing | 2 | 1 | 3 |
| **Neutrophils (x10$^9$/L)** | | | |
| Median (IQR) | 4.9 (3.3, 7.0) | 5.2 (3.5, 7.9) | 5.0 (3.4, 7.5) |
| Min, Max | 1.0, 18.0 | 1.0, 62.6 | 1.0, 62.6 |
| Number missing | 5 | 3 | 8 |
| **Potassium (mmol/L)** | | | |
| Median (IQR) | 4.5 (4.2, 5.0) | 4.5 (4.2, 5.0) | 4.5 (4.2, 5.0) |
| Min, Max | 3.2, 7.8 | 3.2, 6.1 | 3.2, 7.8 |
| Number missing | 1 | 0 | 1 |
| **Urea (mmol/L)** | | | |
| Median (IQR) | 2.8 (2.1, 3.8) | 2.9 (2.1, 3.8) | 2.9 (2.1, 3.8) |
| Min, Max | 1.0, 7.8 | 1.0, 15.4 | 1.0, 15.4 |
| Number missing | 8 | 4 | 12 |
| **Creatinine (mmol/L)** | | | |
| Median (IQR) | 61.0 (51.0, 72.0) | 62.0 (54.0, 74.0) | 61.0 (52.0, 73.0) |
| Min, Max | 32.0, 156.0 | 27.0, 163.0 | 27.0, 163.0 |
| Number missing | 1 | 1 | 2 |
| **ALT (U/L)** | | | |
| Median (IQR) | 19.0 (14.0, 27.0) | 19.0 (14.0, 32.0) | 19.0 (14.0, 30.0) |
| Min, Max | 5.0, 130.0 | 5.0, 148.0 | 5.0, 148.0 |
| Number missing | 8 | 6 | 14 |
| **Total bilirubin (umol/L)** | | | |
| Median (IQR) | 7.0 (5.0, 10.0) | 7.0 (5.0, 10.0) | 7.0 (5.0, 10.0) |
| Min, Max | 0.6, 80.0 | 2.0, 111.0 | 0.6, 111.0 |
| Number missing | 2 | 5 | 7 |
| **Albumin (g/L)** | | | |
| Median (IQR) | 31.0 (27.0, 36.0) | 30.0 (26.0, 35.0) | 30.0 (26.0, 36.0) |
| Min, Max | 17.0, 49.0 | 13.0, 47.0 | 13.0, 49.0 |
| Number missing | 7 | 5 | 12 |
| **Total protein (g/L)** | | | |
| Median (IQR) | 81.0 (76.0, 86.0) | 81.0 (75.0, 86.0) | 81.0 (75.0, 86.0) |
| Min, Max | 57.0, 118.0 | 6.0, 123.0 | 6.0, 123.0 |
| Number missing | 18 | 16 | 34 |
| **Viral load (copies/mm$^3$)** | | | |
| Median (IQR) | 13977.5 (279.0, 68500.0) | 2960.5 (92.5, 114411.0) | 10245.0 (115.0, 86300.0) |
| Min, Max | 0.2, 2.2e+06 | 12.3, 2.0e+06 | 0.2, 2.2e+06 |
| Number missing | 146 | 135 | 281 |

#### 3.3 Treatment Regimens

##### 3.3.1 Treatment prior to Randomization

Table S3.5: Number of weeks of TB treatment prior to randomization.

| Drug | None | <1 wks | 1-<2 wks | 2-<3 wks | 3-<4 wks | ≥4 wks | Total |
|---|---|---|---|---|---|---|---|
| **Study Strategy** | | | | | | | |
| **Any drug** | **50 (24.8%)** | **67 (33.2%)** | **50 (24.8%)** | **23 (11.4%)** | **10 (5.0%)** | **2 (1.0%)** | **202** |
| Bedaquiline | 50 (24.8%) | 68 (33.7%) | 50 (24.8%) | 23 (11.4%) | 10 (5.0%) | 1 (0.5%) | 202 |
| Clofazimine | 54 (26.7%) | 68 (33.7%) | 48 (23.8%) | 21 (10.4%) | 10 (5.0%) | 1 (0.5%) | 202 |
| Delamanid | 189 (93.6%) | 7 (3.5%) | 5 (2.5%) | 1 (0.5%) | 0 (0.0%) | 0 (0.0%) | 202 |
| Ethambutol | 78 (38.6%) | 60 (29.7%) | 39 (19.3%) | 16 (7.9%) | 8 (4.0%) | 1 (0.5%) | 202 |
| Isoniazid | 75 (37.1%) | 59 (29.2%) | 42 (20.8%) | 16 (7.9%) | 9 (4.5%) | 1 (0.5%) | 202 |
| Levofloxacin | 60 (29.7%) | 65 (32.2%) | 45 (22.3%) | 21 (10.4%) | 10 (5.0%) | 1 (0.5%) | 202 |
| Linezolid | 52 (25.7%) | 69 (34.2%) | 49 (24.3%) | 21 (10.4%) | 9 (4.5%) | 2 (1.0%) | 202 |
| Pyrazinamide | 79 (39.1%) | 57 (28.2%) | 39 (19.3%) | 17 (8.4%) | 9 (4.5%) | 1 (0.5%) | 202 |
| **Control Strategy** | | | | | | | |
| **Any drug** | **55 (27.5%)** | **71 (35.5%)** | **38 (19.0%)** | **21 (10.5%)** | **13 (6.5%)** | **2 (1.0%)** | **200** |
| Bedaquiline | 56 (28.0%) | 72 (36.0%) | 38 (19.0%) | 20 (10.0%) | 13 (6.5%) | 1 (0.5%) | 200 |
| Clofazimine | 60 (30.0%) | 69 (34.5%) | 36 (18.0%) | 22 (11.0%) | 12 (6.0%) | 1 (0.5%) | 200 |
| Delamanid | 189 (94.5%) | 4 (2.0%) | 4 (2.0%) | 3 (1.5%) | 0 (0.0%) | 0 (0.0%) | 200 |
| Ethambutol | 75 (37.5%) | 64 (32.0%) | 33 (16.5%) | 15 (7.5%) | 12 (6.0%) | 1 (0.5%) | 200 |
| Isoniazid | 77 (38.5%) | 63 (31.5%) | 32 (16.0%) | 15 (7.5%) | 12 (6.0%) | 1 (0.5%) | 200 |
| Levofloxacin | 65 (32.5%) | 69 (34.5%) | 36 (18.0%) | 16 (8.0%) | 13 (6.5%) | 1 (0.5%) | 200 |
| Linezolid | 58 (29.0%) | 71 (35.5%) | 37 (18.5%) | 20 (10.0%) | 13 (6.5%) | 1 (0.5%) | 200 |
| Pyrazinamide | 76 (38.0%) | 63 (31.5%) | 33 (16.5%) | 15 (7.5%) | 12 (6.0%) | 1 (0.5%) | 200 |

##### 3.3.2 Initial regimen at baseline

Table S3.6: Summary of drugs in initial regimen at randomization. This includes all drugs taken up to 10 days after randomization; does not include drugs taken prior to randomization that were stopped on or before randomization.

| Regimen, N(%) | Control Strategy | Study Strategy |
|---|---|---|
| **Total** | **200** | **202** |
| Bedaquiline, Delamanid, Linezolid, Levofloxacin, Clofazimine | 0 | 145 (71.8%) |
| Bedaquiline, Delamanid, Linezolid, Levofloxacin | 0 | 42 (20.8%) |
| Bedaquiline, Delamanid, Linezolid, Clofazimine | 0 | 14 (6.9%) |
| Bedaquiline, Delamanid, Clofazimine, Ethambutol, Linezolid, Levofloxacin, Pyrazinamide, Rifafour | 0 | 1 (0.5%) |
| Bedaquiline, Clofazimine, Ethambutol, Isoniazid, Linezolid, Levofloxacin, Pyrazinamide | 168 (84.0%) | 0 |
| Bedaquiline, Delamanid, Linezolid, Clofazimine, Terizidone | 16 (8.0%) | 0 |
| Bedaquiline, Linezolid, Levofloxacin, Clofazimine, Terizidone | 8 (4.0%) | 0 |
| Bedaquiline, Delamanid, Linezolid, Clofazimine, PAS | 5 (2.5%) | 0 |
| Bedaquiline, Clofazimine, Ethambutol, Linezolid, Levofloxacin, Pyrazinamide | 2 (1.0%) | 0 |
| Bedaquiline, Ethambutol, Isoniazid, Linezolid, Levofloxacin, Pyrazinamide | 1 (0.5%) | 0 |

##### 3.3.3 Summary of duration of treatment (not including doses reported prior to randomization)

These figures and tables *do not* include any reported treatment prior to randomization.

Figure S3.6: Total cumulative duration of treatment, including any extension and retreatment (not including doses reported prior to randomization).

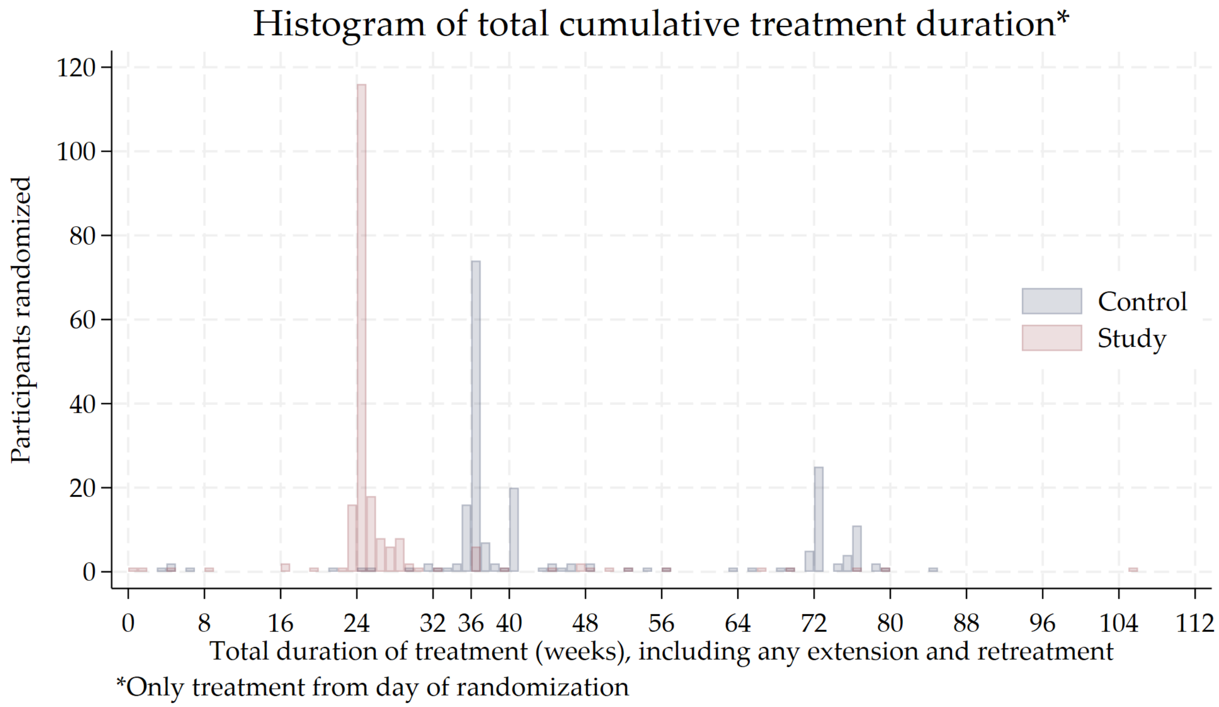

Figure S3.7: Ridge plot of cumulative weeks of treatment by drug and strategy (not including doses reported prior to randomization).

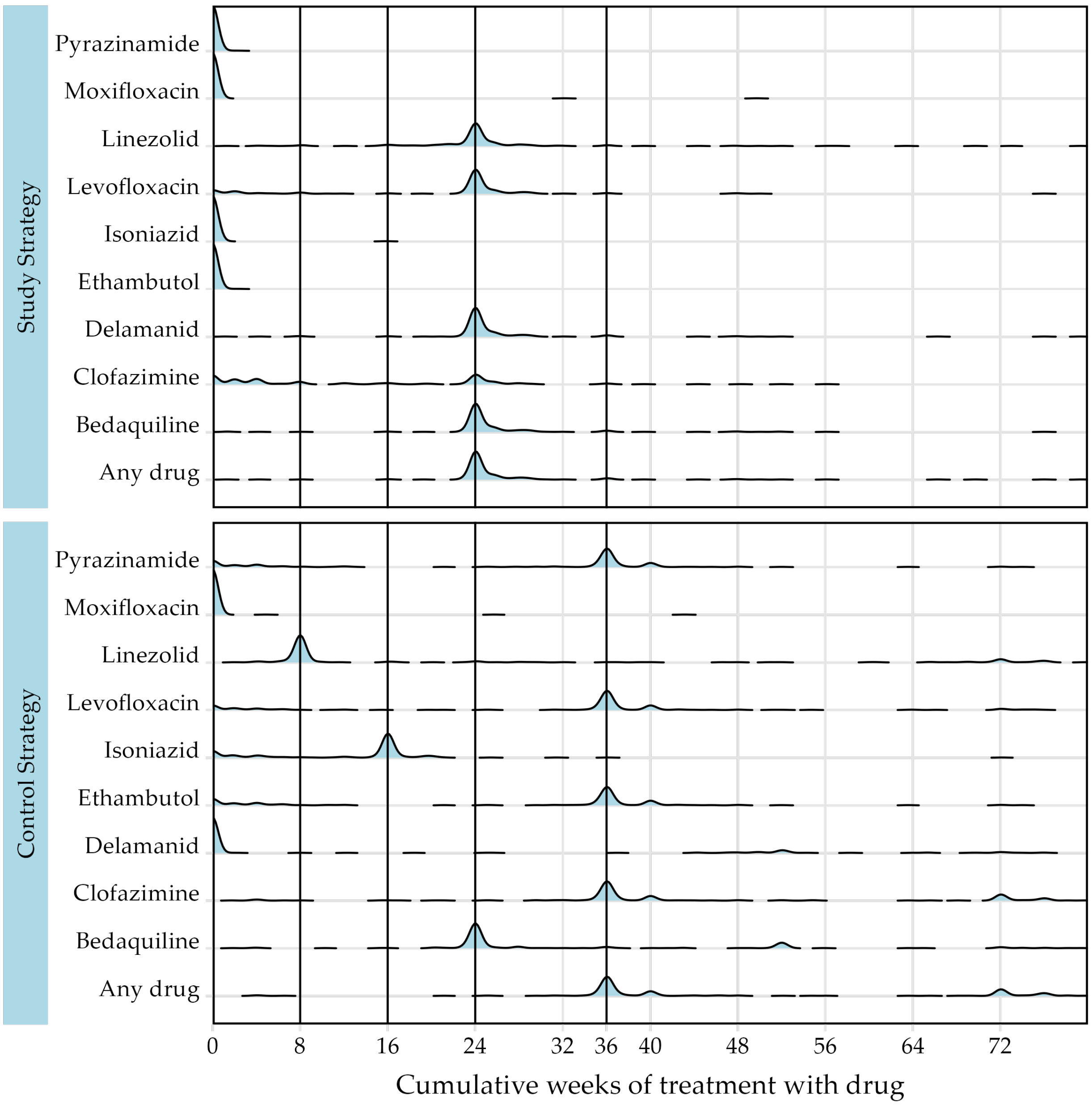

Figure S3.8: Grouped bar chart of cumulative weeks of treatment by drug and strategy (not including doses reported prior to randomization).

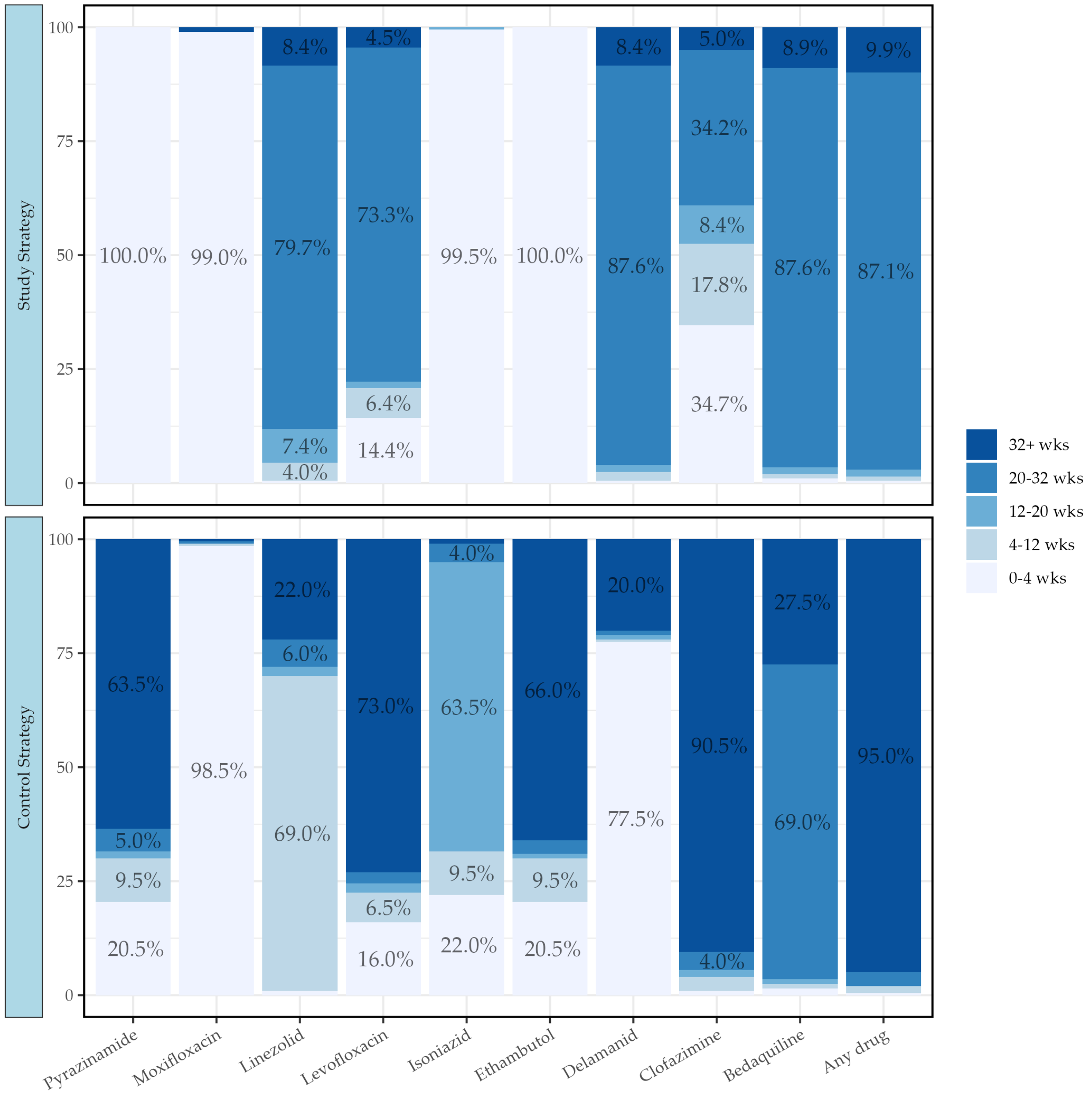

Table S3.7: Grouping of participants by weeks of treatment by category, drug and strategy (not including doses reported prior to randomization).

| Drug | 0-<4 wks | 4-<12 wks | 12-<20 wks | 20-<32 wks | ≥32 wks | Total |
|---|---|---|---|---|---|---|
| **Study Strategy** | | | | | | |
| **Any drug** | **1 (0.5%)** | **2 (1.0%)** | **3 (1.5%)** | **176 (87.1%)** | **20 (9.9%)** | **202** |
| Bedaquiline | 2 (1.0%) | 2 (1.0%) | 3 (1.5%) | 177 (87.6%) | 18 (8.9%) | 202 |
| Clofazimine | 70 (34.7%) | 36 (17.8%) | 17 (8.4%) | 69 (34.2%) | 10 (5.0%) | 202 |
| Delamanid | 1 (0.5%) | 4 (2.0%) | 3 (1.5%) | 177 (87.6%) | 17 (8.4%) | 202 |
| Ethambutol | 202 (100.0%) | 0 (0.0%) | 0 (0.0%) | 0 (0.0%) | 0 (0.0%) | 202 |
| Isoniazid | 201 (99.5%) | 0 (0.0%) | 1 (0.5%) | 0 (0.0%) | 0 (0.0%) | 202 |
| Levofloxacin | 29 (14.4%) | 13 (6.4%) | 3 (1.5%) | 148 (73.3%) | 9 (4.5%) | 202 |
| Linezolid | 1 (0.5%) | 8 (4.0%) | 15 (7.4%) | 161 (79.7%) | 17 (8.4%) | 202 |
| Moxifloxacin | 200 (99.0%) | 0 (0.0%) | 0 (0.0%) | 0 (0.0%) | 2 (1.0%) | 202 |
| Pyrazinamide | 202 (100.0%) | 0 (0.0%) | 0 (0.0%) | 0 (0.0%) | 0 (0.0%) | 202 |
| **Control Strategy** | | | | | | |
| **Any drug** | **1 (0.5%)** | **3 (1.5%)** | **0 (0.0%)** | **6 (3.0%)** | **190 (95.0%)** | **200** |
| Bedaquiline | 3 (1.5%) | 2 (1.0%) | 2 (1.0%) | 138 (69.0%) | 55 (27.5%) | 200 |
| Clofazimine | 2 (1.0%) | 6 (3.0%) | 3 (1.5%) | 8 (4.0%) | 181 (90.5%) | 200 |
| Delamanid | 155 (77.5%) | 1 (0.5%) | 2 (1.0%) | 2 (1.0%) | 40 (20.0%) | 200 |
| Ethambutol | 41 (20.5%) | 19 (9.5%) | 2 (1.0%) | 6 (3.0%) | 132 (66.0%) | 200 |
| Isoniazid | 44 (22.0%) | 19 (9.5%) | 127 (63.5%) | 8 (4.0%) | 2 (1.0%) | 200 |
| Levofloxacin | 32 (16.0%) | 13 (6.5%) | 4 (2.0%) | 5 (2.5%) | 146 (73.0%) | 200 |
| Linezolid | 2 (1.0%) | 138 (69.0%) | 4 (2.0%) | 12 (6.0%) | 44 (22.0%) | 200 |
| Moxifloxacin | 197 (98.5%) | 1 (0.5%) | 0 (0.0%) | 1 (0.5%) | 1 (0.5%) | 200 |
| Pyrazinamide | 41 (20.5%) | 19 (9.5%) | 3 (1.5%) | 10 (5.0%) | 127 (63.5%) | 200 |

Table S3.8: Summary of cumulative weeks of treatment by drug and strategy (not including doses reported prior to randomization).

| | Control: Total | Control: Med (IQR) | Study: Total | Study: Med (IQR) | Overall: Total | Overall: Med (IQR) |
|---|---|---|---|---|---|---|
| **Total Randomised** | **200 (100.0%)** | | **203 (100.0%)** | | **403 (100.0%)** | |
| **Any treatment** | **200 (100.0%)** | **36.4 (36.0, 71.7)** | **202 (99.5%)** | **24.1 (24.0, 25.6)** | **402 (99.8%)** | **35.3 (24.1, 38.4)** |
| Bedaquiline | 200 (100.0%) | 24.1 (24.0, 36.0) | 202 (99.5%) | 24.1 (24.0, 25.4) | 402 (99.8%) | 24.1 (24.0, 26.3) |
| Linezolid | 200 (100.0%) | 8.1 (8.0, 24.3) | 202 (99.5%) | 24.0 (23.7, 25.0) | 402 (99.8%) | 23.9 (8.1, 25.0) |
| Delamanid | 46 (23.0%) | 52.0 (48.0, 65.3) | 202 (99.5%) | 24.0 (24.0, 25.1) | 248 (61.5%) | 24.1 (24.0, 28.6) |
| Clofazimine | 200 (100.0%) | 36.1 (36.0, 52.0) | 163 (80.3%) | 19.1 (4.0, 24.1) | 363 (90.1%) | 35.3 (17.0, 37.0) |
| Levofloxacin | 180 (90.0%) | 36.1 (35.4, 39.1) | 187 (92.1%) | 24.0 (24.0, 24.4) | 367 (91.1%) | 25.4 (24.0, 36.1) |
| Moxifloxacin | 3 (1.5%) | 25.7 (4.9, 43.1) | 2 (1.0%) | 40.9 (32.1, 49.7) | 5 (1.2%) | 32.1 (25.7, 43.1) |
| Isoniazid | 171 (85.5%) | 16.0 (15.4, 16.1) | 2 (1.0%) | 8.4 (1.0, 15.9) | 173 (42.9%) | 16.0 (15.4, 16.1) |
| PAS | 37 (18.5%) | 22.6 (4.9, 48.3) | 5 (2.5%) | 32.1 (14.6, 54.6) | 42 (10.4%) | 24.1 (4.9, 48.4) |
| Terizidone | 56 (28.0%) | 64.1 (30.9, 71.4) | 6 (3.0%) | 27.9 (15.0, 50.1) | 62 (15.4%) | 62.3 (24.3, 71.1) |
| Ethambutol | 172 (86.0%) | 36.0 (34.1, 36.4) | 2 (1.0%) | 1.6 (1.0, 2.3) | 174 (43.2%) | 36.0 (33.1, 36.3) |
| Pyrazinamide | 172 (86.0%) | 36.0 (31.1, 36.3) | 2 (1.0%) | 1.6 (1.0, 2.3) | 174 (43.2%) | 36.0 (29.6, 36.3) |
| Meropenem | 6 (3.0%) | 26.2 (25.9, 26.3) | 5 (2.5%) | 26.1 (14.6, 32.1) | 11 (2.7%) | 26.1 (14.6, 27.1) |
| Augmentin | 6 (3.0%) | 26.3 (10.1, 43.1) | 5 (2.5%) | 32.1 (14.6, 52.7) | 11 (2.7%) | 26.3 (10.1, 52.7) |
| Rifabutin | 1 (0.5%) | 26.3 (26.3, 26.3) | 1 (0.5%) | 60.3 (60.3, 60.3) | 2 (0.5%) | 43.3 (26.3, 60.3) |

Table S3.9: Among participants allocated to the Study Strategy, grouping by weeks of treatment with Clofazimine and Levofloxacin by baseline resistance to fluoroquinolones (not including doses reported prior to randomization).

| Drug | 0-<4 wks | 4-<12 wks | 12-<20 wks | 20-<32 wks | ≥32 wks | Total |
|---|---|---|---|---|---|---|
| **FQ-Sensitive** | | | | | | |
| Clofazimine | 68 (55.3%) | 34 (27.6%) | 13 (10.6%) | 8 (6.5%) | 0 | 123 |
| Levofloxacin | 1 (0.8%) | 1 (0.8%) | 3 (2.4%) | 110 (89.4%) | 8 (6.5%) | 123 |
| **FQ-Resistant** | | | | | | |
| Clofazimine | 1 (2.4%) | 1 (2.4%) | 1 (2.4%) | 30 (71.4%) | 9 (21.4%) | 42 |
| Levofloxacin | 26 (61.9%) | 12 (28.6%) | 0 | 4 (9.5%) | 0 | 42 |
| **FQ-Indeterminate** | | | | | | |
| Clofazimine | 0 | 0 | 1 (33.3%) | 2 (66.7%) | 0 | 3 |
| Levofloxacin | 0 | 0 | 0 | 3 (100.0%) | 0 | 3 |
| **FQ-Missing** | | | | | | |
| Clofazimine | 1 (2.9%) | 1 (2.9%) | 2 (5.9%) | 29 (85.3%) | 1 (2.9%) | 34 |
| Levofloxacin | 2 (5.9%) | 0 | 0 | 31 (91.2%) | 1 (2.9%) | 34 |
| **Total** | | | | | | |
| Clofazimine | 70 (34.7%) | 36 (17.8%) | 17 (8.4%) | 69 (34.2%) | 10 (5.0%) | 202 |
| Levofloxacin | 29 (14.4%) | 13 (6.4%) | 3 (1.5%) | 148 (73.3%) | 9 (4.5%) | 202 |

Table S3.10: Study regimen composition (total duration during study, not including doses prior to randomization).

| Regimen, N(%) | Sensitive | Resistant | Missing/Indeterminate | Total |
|---|---|---|---|---|
| 20-28wk BDLzC (Lf < 12wks) | 0 | 24 (54.5%) | 1 (2.8%) | 25 (12.4%) |
| 20-28wk BDLzLf (C < 12wks) | 79 (64.8%) | 3 (6.8%) | 1 (2.8%) | 83 (41.1%) |
| 20-28wk BDLzLf (C < 20wks) | 12 (9.8%) | 0 | 3 (8.3%) | 15 (7.4%) |
| 24wk BDLzLfC | 13 (10.7%) | 4 (9.1%) | 28 (77.8%) | 45 (22.3%) |
| <20wk treatment* | 3 (2.5%) | 2 (4.5%) | 1 (2.8%) | 6 (3.0%) |
| >28wk treatment | 15 (12.3%) | 11 (25.0%) | 2 (5.6%) | 28 (13.9%) |
| **Total** | **122** | **44** | **36** | **202** |

*Note:* B - Bedaquiline, D - Delamanid, Lz - Linezolid, Lf - Levofloxacin, C - Clofazimine.
* All 6 participants withdrew consent, were lost, or died prior to 20 weeks.

- 28 participants had treatment extension/retreatment beyond 28 weeks
  - Making up for missed doses and treatment interruptions: 12
  - Treatment failed: 7
  - Recurrence: 5
  - Late culture conversion: 3
  - N/A (2 days over 28 weeks) : 1

##### 3.3.4 Summary of duration of treatment (including doses reported prior to randomization)

These figures and tables *include any reported treatment prior to randomization* (as part of the same TB event).

Figure S3.9: Total cumulative duration of treatment, including any extension and retreatment (including doses reported prior to randomization).

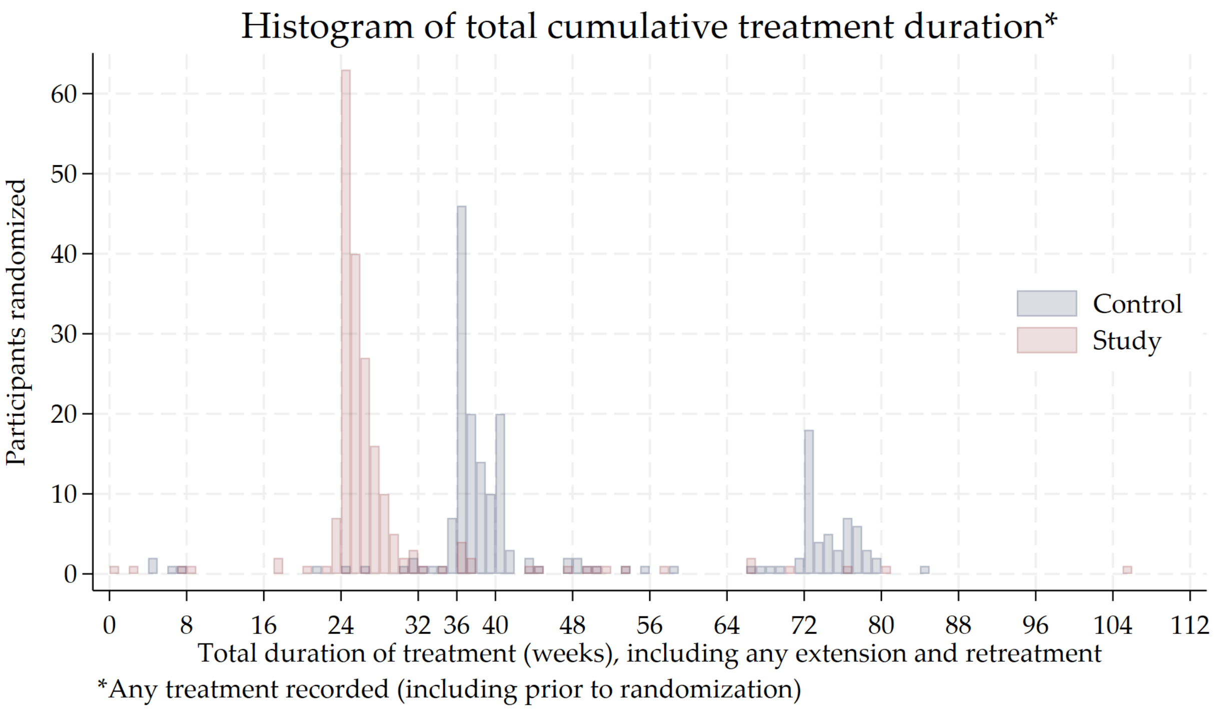

Figure S3.10: Ridge plot of cumulative weeks of treatment by drug and strategy (including doses reported prior to randomization).

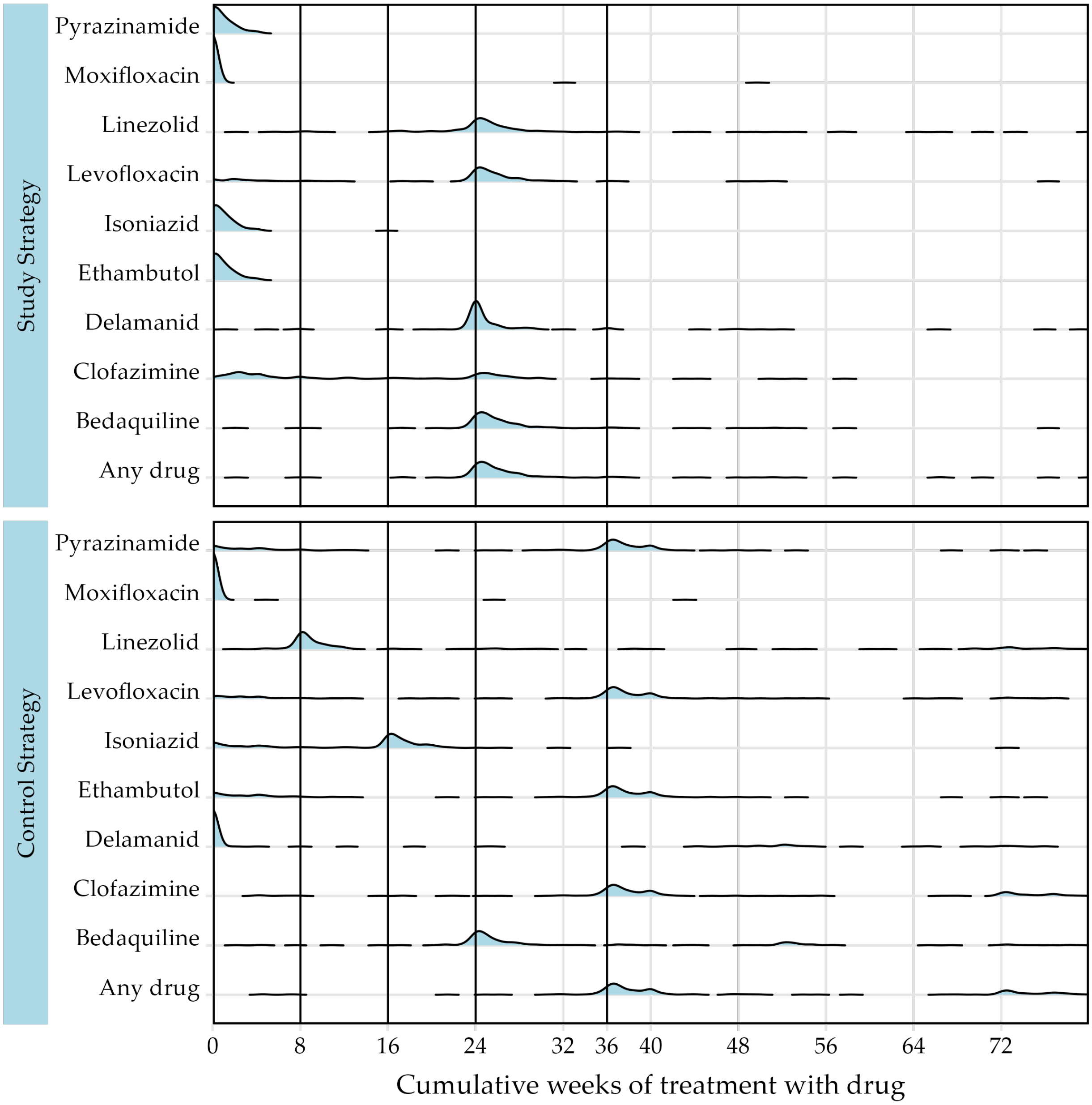

Figure S3.11: Grouped bar chart of cumulative weeks of treatment by drug and strategy (including doses reported prior to randomization).

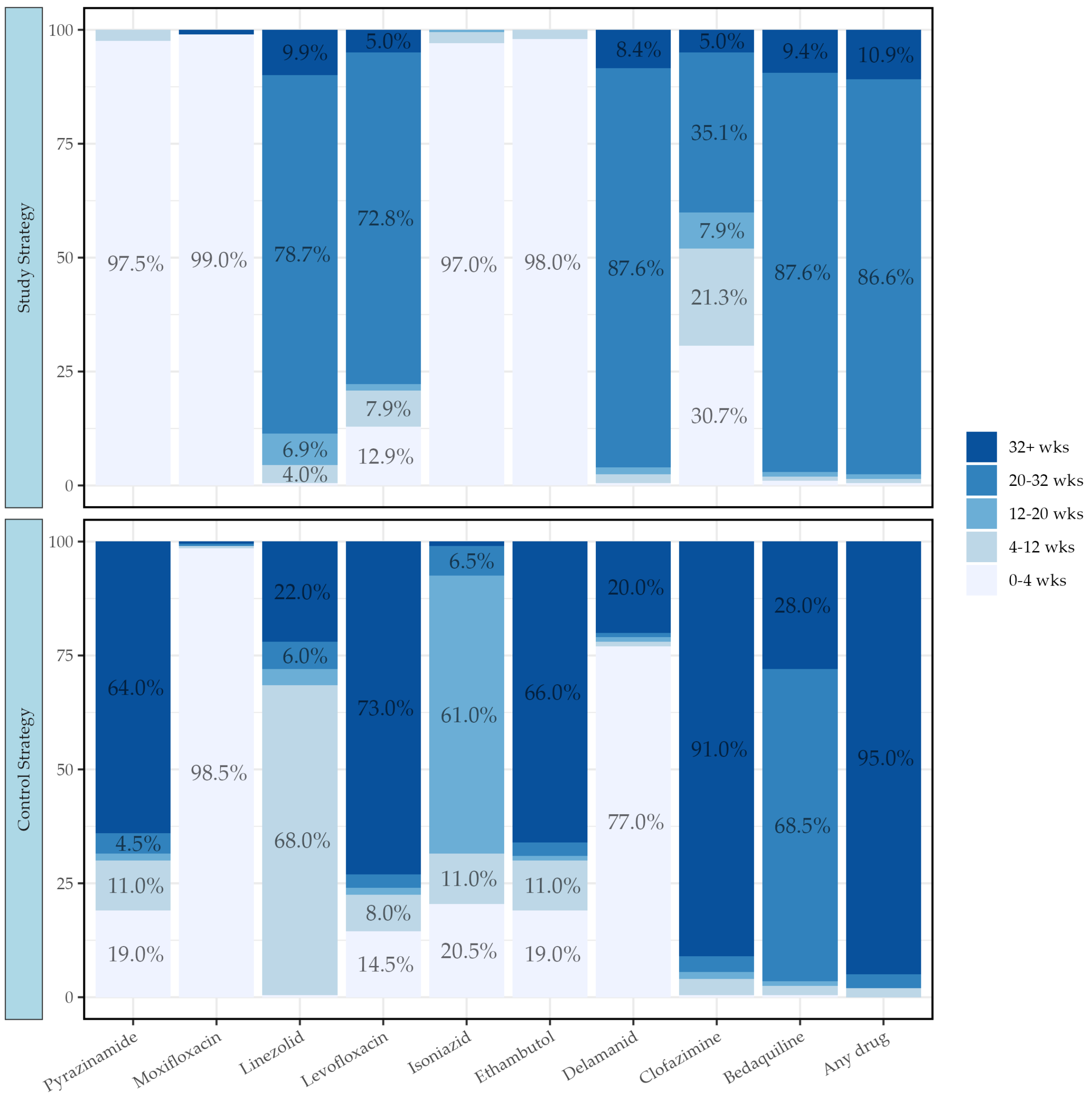

Table S3.11: Grouping of participants by weeks of treatment by category, drug and strategy (including doses reported prior to randomization).

| Drug | 0-<4 wks | 4-<12 wks | 12-<20 wks | 20-<32 wks | ≥32 wks | Total |
|---|---|---|---|---|---|---|
| **Study Strategy** | | | | | | |
| Any drug | 1 (0.5%) | 2 (1.0%) | 2 (1.0%) | 175 (86.6%) | 22 (10.9%) | 202 |
| Bedaquiline | 2 (1.0%) | 2 (1.0%) | 2 (1.0%) | 177 (87.6%) | 19 (9.4%) | 202 |
| Clofazimine | 62 (30.7%) | 43 (21.3%) | 16 (7.9%) | 71 (35.1%) | 10 (5.0%) | 202 |
| Delamanid | 1 (0.5%) | 4 (2.0%) | 3 (1.5%) | 177 (87.6%) | 17 (8.4%) | 202 |
| Ethambutol | 198 (98.0%) | 4 (2.0%) | 0 (0.0%) | 0 (0.0%) | 0 (0.0%) | 202 |
| Isoniazid | 196 (97.0%) | 5 (2.5%) | 1 (0.5%) | 0 (0.0%) | 0 (0.0%) | 202 |
| Levofloxacin | 26 (12.9%) | 16 (7.9%) | 3 (1.5%) | 147 (72.8%) | 10 (5.0%) | 202 |
| Linezolid | 1 (0.5%) | 8 (4.0%) | 14 (6.9%) | 159 (78.7%) | 20 (9.9%) | 202 |
| Moxifloxacin | 200 (99.0%) | 0 (0.0%) | 0 (0.0%) | 0 (0.0%) | 2 (1.0%) | 202 |
| Pyrazinamide | 197 (97.5%) | 5 (2.5%) | 0 (0.0%) | 0 (0.0%) | 0 (0.0%) | 202 |
| **Control Strategy** | | | | | | |
| Any drug | 0 (0.0%) | 4 (2.0%) | 0 (0.0%) | 6 (3.0%) | 190 (95.0%) | 200 |
| Bedaquiline | 1 (0.5%) | 4 (2.0%) | 2 (1.0%) | 137 (68.5%) | 56 (28.0%) | 200 |
| Clofazimine | 1 (0.5%) | 7 (3.5%) | 3 (1.5%) | 7 (3.5%) | 182 (91.0%) | 200 |
| Delamanid | 154 (77.0%) | 2 (1.0%) | 2 (1.0%) | 2 (1.0%) | 40 (20.0%) | 200 |
| Ethambutol | 38 (19.0%) | 22 (11.0%) | 2 (1.0%) | 6 (3.0%) | 132 (66.0%) | 200 |
| Isoniazid | 41 (20.5%) | 22 (11.0%) | 122 (61.0%) | 13 (6.5%) | 2 (1.0%) | 200 |
| Levofloxacin | 29 (14.5%) | 16 (8.0%) | 3 (1.5%) | 6 (3.0%) | 146 (73.0%) | 200 |
| Linezolid | 1 (0.5%) | 136 (68.0%) | 7 (3.5%) | 12 (6.0%) | 44 (22.0%) | 200 |
| Moxifloxacin | 197 (98.5%) | 1 (0.5%) | 0 (0.0%) | 1 (0.5%) | 1 (0.5%) | 200 |
| Pyrazinamide | 38 (19.0%) | 22 (11.0%) | 3 (1.5%) | 9 (4.5%) | 128 (64.0%) | 200 |

Table S3.12: Summary of cumulative weeks of treatment by drug and strategy (including doses reported prior to randomization).

| | Control: Total | Control: Med (IQR) | Study: Total | Study: Med (IQR) | Overall: Total | Overall: Med (IQR) |
|---|---|---|---|---|---|---|
| **Total Randomised** | **200 (100.0%)** | | **203 (100.0%)** | | **403 (100.0%)** | |
| **Any treatment** | **200 (100.0%)** | **38.9 (36.6, 71.8)** | **202 (99.5%)** | **25.2 (24.3, 27.3)** | **402 (99.8%)** | **35.8 (25.1, 40.0)** |
| Bedaquiline | 200 (100.0%) | 25.3 (24.1, 37.1) | 202 (99.5%) | 25.1 (24.3, 27.1) | 402 (99.8%) | 25.2 (24.1, 28.0) |
| Linezolid | 200 (100.0%) | 9.3 (8.1, 25.9) | 202 (99.5%) | 24.9 (24.0, 26.7) | 402 (99.8%) | 24.0 (8.9, 26.6) |
| Delamanid | 47 (23.5%) | 52.1 (47.3, 65.3) | 202 (99.5%) | 24.1 (24.0, 25.4) | 249 (61.8%) | 24.1 (24.0, 28.7) |
| Clofazimine | 200 (100.0%) | 38.0 (36.3, 52.9) | 198 (97.5%) | 9.4 (3.0, 25.0) | 398 (98.8%) | 29.5 (8.0, 38.4) |
| Levofloxacin | 193 (96.5%) | 36.9 (33.9, 39.6) | 193 (95.1%) | 24.7 (24.0, 26.1) | 386 (95.8%) | 26.3 (24.0, 36.9) |
| Moxifloxacin | 3 (1.5%) | 25.7 (4.9, 43.1) | 2 (1.0%) | 40.9 (32.1, 49.7) | 5 (1.2%) | 32.1 (25.7, 43.1) |
| Isoniazid | 182 (91.0%) | 16.2 (12.0, 17.6) | 129 (63.5%) | 1.1 (0.7, 1.9) | 311 (77.2%) | 4.3 (1.1, 16.6) |
| PAS | 37 (18.5%) | 22.6 (4.9, 48.3) | 16 (7.9%) | 2.1 (1.0, 9.1) | 53 (13.2%) | 12.3 (3.6, 35.9) |
| Terizidone | 59 (29.5%) | 64.0 (24.1, 72.0) | 19 (9.4%) | 1.9 (1.0, 15.0) | 78 (19.4%) | 51.6 (5.0, 68.9) |
| Ethambutol | 184 (92.0%) | 36.6 (17.1, 38.7) | 125 (61.6%) | 1.0 (0.7, 1.9) | 309 (76.7%) | 4.6 (1.1, 36.9) |
| Pyrazinamide | 184 (92.0%) | 36.4 (12.9, 38.2) | 125 (61.6%) | 1.1 (0.7, 2.0) | 309 (76.7%) | 4.6 (1.3, 36.9) |
| Meropenem | 6 (3.0%) | 26.2 (25.9, 26.3) | 5 (2.5%) | 26.1 (14.6, 32.1) | 11 (2.7%) | 26.1 (14.6, 27.1) |
| Augmentin | 6 (3.0%) | 26.3 (10.1, 43.1) | 5 (2.5%) | 32.1 (14.6, 52.7) | 11 (2.7%) | 26.3 (10.1, 52.7) |
| Rifabutin | 1 (0.5%) | 26.3 (26.3, 26.3) | 1 (0.5%) | 60.3 (60.3, 60.3) | 2 (0.5%) | 43.3 (26.3, 60.3) |

Table S3.13: Among participants allocated to the Study Strategy, grouping by weeks of treatment with Clofazimine and Levofloxacin by baseline resistance to fluoroquinolones (including doses reported prior to randomization).

| Drug | 0-<4 wks | 4-<12 wks | 12-<20 wks | 20-<32 wks | ≥32 wks | Total |
|---|---|---|---|---|---|---|
| **FQ-Sensitive** | | | | | | |
| Clofazimine | 60 (48.8%) | 41 (33.3%) | 14 (11.4%) | 8 (6.5%) | 0 | 123 |
| Levofloxacin | 1 (0.8%) | 1 (0.8%) | 3 (2.4%) | 109 (88.6%) | 9 (7.3%) | 123 |
| **FQ-Resistant** | | | | | | |
| Clofazimine | 1 (2.4%) | 1 (2.4%) | 0 | 31 (73.8%) | 9 (21.4%) | 42 |
| Levofloxacin | 23 (54.8%) | 15 (35.7%) | 0 | 4 (9.5%) | 0 | 42 |
| **FQ-Indeterminate** | | | | | | |
| Clofazimine | 0 | 0 | 1 (33.3%) | 2 (66.7%) | 0 | 3 |
| Levofloxacin | 0 | 0 | 0 | 3 (100.0%) | 0 | 3 |
| **FQ-Missing** | | | | | | |
| Clofazimine | 1 (2.9%) | 1 (2.9%) | 1 (2.9%) | 30 (88.2%) | 1 (2.9%) | 34 |
| Levofloxacin | 2 (5.9%) | 0 | 0 | 31 (91.2%) | 1 (2.9%) | 34 |
| **Total** | | | | | | |
| Clofazimine | 62 (30.7%) | 43 (21.3%) | 16 (7.9%) | 71 (35.1%) | 10 (5.0%) | 202 |
| Levofloxacin | 26 (12.9%) | 16 (7.9%) | 3 (1.5%) | 147 (72.8%) | 10 (5.0%) | 202 |

##### 3.4 Safety: Adverse Events

Table S3.14: Overall safety summary by strategy.

| | Control Strategy | Study Strategy | Total | RD (95% CI) |
|---|---|---|---|---|
| **Randomised and starting treatment** | **200** | **202** | **402** | |
| Grade 3-5 AEs | 76 (38.0%) | 69 (34.2%) | 145 (36.1%) | 3.8% (-5.5%, 13.2%) |
| Grade 3-5 AEs during treatment | 74 (37.0%) | 63 (31.2%) | 137 (34.1%) | 5.8% (-3.4%, 15.1%) |
| Grade 3-5 AEs during treatment, at least possibly related | 56 (28.0%) | 52 (25.7%) | 108 (26.9%) | 2.3% (-6.4%, 10.9%) |
| SAEs | 44 (22.0%) | 45 (22.3%) | 89 (22.1%) | -0.3% (-8.4%, 7.8%) |
| SAEs during treatment | 42 (21.0%) | 38 (18.8%) | 80 (19.9%) | 2.2% (-5.6%, 10.0%) |
| Notable Events (NEs) | 31 (15.5%) | 36 (17.8%) | 67 (16.7%) | -2.3% (-9.6%, 5.0%) |
| Death at any time | 10 (5.0%) | 10 (5.0%) | 20 (5.0%) | 0.0% (-4.2%, 4.3%) |
| Death during treatment | 8 (4.0%) | 6 (3.0%) | 14 (3.5%) | 1.0% (-2.6%, 4.6%) |
| Death after treatment | 2 (1.0%) | 4 (2.0%) | 6 (1.5%) | -1.0% (-3.3%, 1.4%) |
| Anaemia leading to treatment discontinuation | 8 (4.0%) | 10 (5.0%) | 18 (4.5%) | -1.0% (-5.0%, 3.1%) |
| Anaemia leading to blood transfusion | 11 (5.5%) | 20 (9.9%) | 31 (7.7%) | -4.4% (-9.6%, 0.8%) |
| Grade 3-5 liver abnormality | 9 (4.5%) | 4 (2.0%) | 13 (3.2%) | 2.5% (-0.9%, 6.0%) |
| Perihperal neuropathy leading to treatment discontinuation | 7 (3.5%) | 10 (5.0%) | 17 (4.2%) | -1.5% (-5.4%, 2.5%) |
| Optic neuropathy leading to treatment discontinuation | 2 (1.0%) | 7 (3.5%) | 9 (2.2%) | -2.5% (-5.3%, 0.4%) |
| QTcF $\geq$ 480ms | 19 (9.5%) | 12 (5.9%) | 31 (7.7%) | 3.6% (-1.7%, 8.8%) |
| QTcF $\geq$ 500ms | 7 (3.5%) | 5 (2.5%) | 12 (3.0%) | 1.0% (-2.3%, 4.4%) |

- A **Notable event (NE)** is any one of the following:
  - Grade 3 and 4 cardiac adverse event
  - Grade 3 or 4 liver toxicity
  - One or more drugs may need to be suspended permanently due to severe toxicity
  - Pregnancy occurs in female participants during the trial (both on or off treatment)

Figure S3.12: Kaplan-Meier of time to first grade 3-5 AE by strategy.

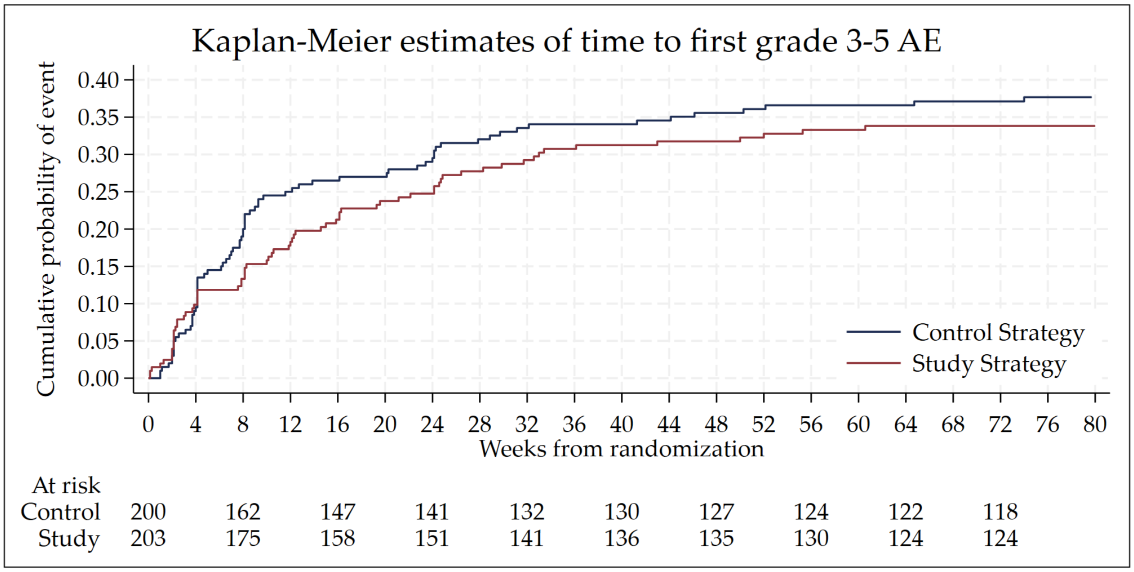

Figure S3.13: Kaplan-Meier of time to death by strategy.

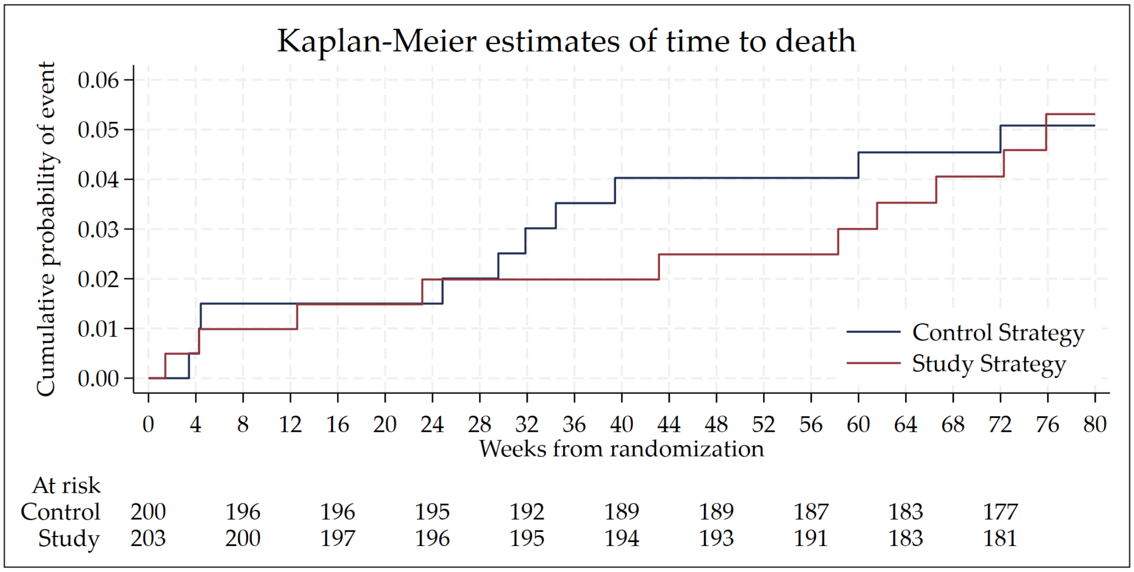

Table S3.15: Listing of all deaths: Control strategy.

| | Primary AE | Weeks from randomization |
|---|---|---|
| 1 | Sepsis | 3.4 |
| 2 | Dyspnoea | 4.3 |
| 3 | Hiv Related Iris | 4.4 |
| 4 | Meningitis cryptococcal | 24.9 |
| 5 | Gastrointestinal disorder | 29.6 |
| 6 | Meningitis tuberculous | 31.9 |
| 7 | Hemiplegia, Central nervous system lesion | 34.4 |
| 8 | Seizure | 39.4 |
| 9 | Acute Diarrhoea | 60.0 |
| 10 | Sudden Death | 72.0 |

Table S3.16: Listing of all deaths: Study strategy.

| | Primary AE | Weeks from randomization |
|---|---|---|
| 11 | Respiratory distress | 1.4 |
| 12 | SARS-CoV-2 test positive, Anaemia | 4.3 |
| 13 | Anaemia | 12.6 |
| 14 | Gun shot wound | 23.1 |
| 15 | Angiopathy | 43.1 |
| 16 | Chronic respiratory disease | 58.3 |
| 17 | 11-beta-hydroxylase deficiency | 61.6 |
| 18 | Pancreatitis, Anaemia | 66.6 |
| 19 | Sudden Death | 72.3 |
| 20 | Dyspnoea | 75.9 |

Table S3.17: Summary of grade 3-5 AEs by MedDRA coding and strategy sorted by decreasing frequency.

| | Control Strategy | Study Strategy | Total |
|---|---|---|---|
| **Total randomized** | **200** | **202** | **402** |
| **No Grade 3-5 AE** | **124 (62.0%)** | **133 (65.8%)** | **257 (63.9%)** |
| **Any Grade 3-5 AE** | **76 (38.0%)** | **69 (34.2%)** | **145 (36.1%)** |
| Anaemia | 29 (14.5%) | 33 (16.3%) | 62 (15.4%) |
| Neuropathy peripheral | 12 (6.0%) | 16 (7.9%) | 28 (7.0%) |
| Alanine aminotransferase increased | 9 (4.5%) | 4 (2.0%) | 13 (3.2%) |
| Electrocardiogram QT prolonged | 7 (3.5%) | 6 (3.0%) | 13 (3.2%) |
| Optic neuritis | 2 (1.0%) | 5 (2.5%) | 7 (1.7%) |
| Dyspnoea | 2 (1.0%) | 3 (1.5%) | 5 (1.2%) |
| Treatment failure | 4 (2.0%) | 1 (0.5%) | 5 (1.2%) |
| Hepatotoxicity | 2 (1.0%) | 1 (0.5%) | 3 (0.7%) |
| Not yet coded | 2 (1.0%) | 1 (0.5%) | 3 (0.7%) |
| Pneumonia | 1 (0.5%) | 2 (1.0%) | 3 (0.7%) |
| 11-beta-hydroxylase deficiency | 0 | 2 (1.0%) | 2 (0.5%) |
| Acute kidney injury | 0 | 2 (1.0%) | 2 (0.5%) |
| Arthralgia | 1 (0.5%) | 1 (0.5%) | 2 (0.5%) |
| Back pain | 0 | 2 (1.0%) | 2 (0.5%) |
| Gastritis | 2 (1.0%) | 0 | 2 (0.5%) |
| Gastroenteritis | 2 (1.0%) | 0 | 2 (0.5%) |
| Meningitis cryptococcal | 1 (0.5%) | 1 (0.5%) | 2 (0.5%) |
| Myelosuppression | 1 (0.5%) | 1 (0.5%) | 2 (0.5%) |
| Neutropenia | 1 (0.5%) | 1 (0.5%) | 2 (0.5%) |
| Renal impairment | 1 (0.5%) | 1 (0.5%) | 2 (0.5%) |
| Respiratory distress | 0 | 2 (1.0%) | 2 (0.5%) |
| Sepsis | 1 (0.5%) | 1 (0.5%) | 2 (0.5%) |
| Thrombocytopenia | 1 (0.5%) | 1 (0.5%) | 2 (0.5%) |
| Weight decreased | 2 (1.0%) | 0 | 2 (0.5%) |
| Abdominal discomfort | 1 (0.5%) | 0 | 1 (0.2%) |
| Acarodermatitis | 1 (0.5%) | 0 | 1 (0.2%) |
| Acute abdomen | 1 (0.5%) | 0 | 1 (0.2%) |
| Acute coronary syndrome | 0 | 1 (0.5%) | 1 (0.2%) |
| Acute hepatitis B | 1 (0.5%) | 0 | 1 (0.2%) |
| Angiopathy | 0 | 1 (0.5%) | 1 (0.2%) |
| Appendicitis | 1 (0.5%) | 0 | 1 (0.2%) |
| Arthritis | 0 | 1 (0.5%) | 1 (0.2%) |
| Arthritis bacterial | 0 | 1 (0.5%) | 1 (0.2%) |
| Blood creatinine increased | 1 (0.5%) | 0 | 1 (0.2%) |
| Cardiotoxicity | 1 (0.5%) | 0 | 1 (0.2%) |
| Cataract operation | 0 | 1 (0.5%) | 1 (0.2%) |
| Central nervous system lesion | 1 (0.5%) | 0 | 1 (0.2%) |
| Chronic obstructive pulmonary disease | 0 | 1 (0.5%) | 1 (0.2%) |
| Chronic respiratory disease | 0 | 1 (0.5%) | 1 (0.2%) |
| Drug-induced liver injury | 1 (0.5%) | 0 | 1 (0.2%) |
| Erythema | 1 (0.5%) | 0 | 1 (0.2%) |
| Facial bones fracture | 0 | 1 (0.5%) | 1 (0.2%) |
| Fixed eruption | 1 (0.5%) | 0 | 1 (0.2%) |
| Gastrointestinal disorder | 1 (0.5%) | 0 | 1 (0.2%) |
| General physical health deterioration | 1 (0.5%) | 0 | 1 (0.2%) |
| Glomerular filtration rate decreased | 1 (0.5%) | 0 | 1 (0.2%) |
| Gun shot wound | 0 | 1 (0.5%) | 1 (0.2%) |
| Haemoptysis | 0 | 1 (0.5%) | 1 (0.2%) |
| Headache | 1 (0.5%) | 0 | 1 (0.2%) |

Table S3.17: Summary of grade 3-5 AEs by MedDRA coding and strategy sorted by decreasing frequency. *(continued)*

| | Control Strategy | Study Strategy | Total |
|---|---|---|---|
| Hemiplegia | 1 (0.5%) | 0 | 1 (0.2%) |
| Hepatic failure | 1 (0.5%) | 0 | 1 (0.2%) |
| Hyperglycaemia | 0 | 1 (0.5%) | 1 (0.2%) |
| Hypotension | 0 | 1 (0.5%) | 1 (0.2%) |
| IRIS associated TB | 1 (0.5%) | 0 | 1 (0.2%) |
| Immunosuppression | 1 (0.5%) | 0 | 1 (0.2%) |
| Keratitis | 1 (0.5%) | 0 | 1 (0.2%) |
| Lip lesion excision | 1 (0.5%) | 0 | 1 (0.2%) |
| Liver injury | 1 (0.5%) | 0 | 1 (0.2%) |
| Lower respiratory tract infection | 1 (0.5%) | 0 | 1 (0.2%) |
| Lung abscess | 0 | 1 (0.5%) | 1 (0.2%) |
| Meningitis tuberculous | 1 (0.5%) | 0 | 1 (0.2%) |
| Multiple fractures | 0 | 1 (0.5%) | 1 (0.2%) |
| Multiple injuries | 0 | 1 (0.5%) | 1 (0.2%) |
| Myalgia | 0 | 1 (0.5%) | 1 (0.2%) |
| Nausea | 1 (0.5%) | 0 | 1 (0.2%) |
| Pancreatitis | 0 | 1 (0.5%) | 1 (0.2%) |
| Physical assault | 0 | 1 (0.5%) | 1 (0.2%) |
| Premature rupture of membranes | 1 (0.5%) | 0 | 1 (0.2%) |
| Psychotic disorder | 0 | 1 (0.5%) | 1 (0.2%) |
| Pyelonephritis | 0 | 1 (0.5%) | 1 (0.2%) |
| Rash papular | 1 (0.5%) | 0 | 1 (0.2%) |
| Renal stone removal | 0 | 1 (0.5%) | 1 (0.2%) |
| SARS-CoV-2 test positive | 0 | 1 (0.5%) | 1 (0.2%) |
| Seizure | 1 (0.5%) | 0 | 1 (0.2%) |
| Skin mass | 0 | 1 (0.5%) | 1 (0.2%) |
| Stab wound | 0 | 1 (0.5%) | 1 (0.2%) |
| Substance abuse | 1 (0.5%) | 0 | 1 (0.2%) |
| Traumatic haemothorax | 1 (0.5%) | 0 | 1 (0.2%) |
| Treatment noncompliance | 0 | 1 (0.5%) | 1 (0.2%) |
| Tuberculosis | 0 | 1 (0.5%) | 1 (0.2%) |
| Vasculitic rash | 1 (0.5%) | 0 | 1 (0.2%) |
| Vitamin B12 deficiency | 1 (0.5%) | 0 | 1 (0.2%) |
| Vomiting | 1 (0.5%) | 0 | 1 (0.2%) |

#### 3.5 Safety laboratory parameter summaries

Figure S3.14: Mean ALT over time.

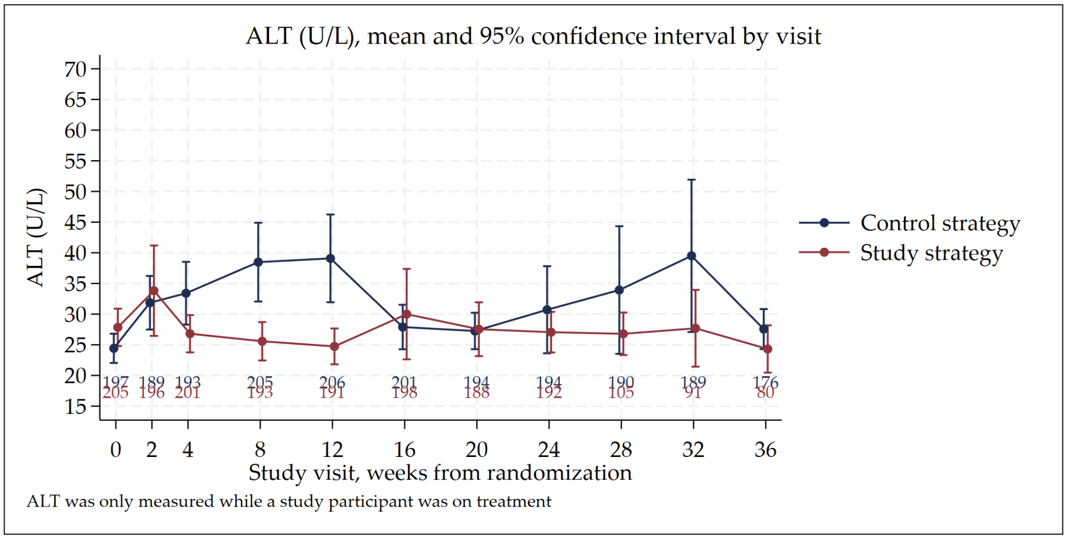

Figure S3.15: Mean creatinine over time.

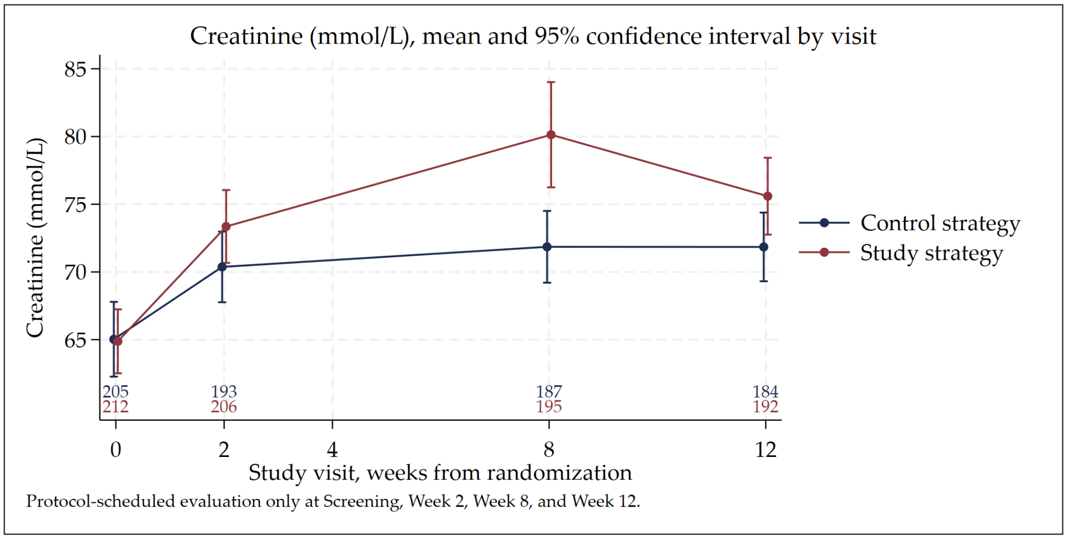

Figure S3.16: Mean haemoglobin over time.

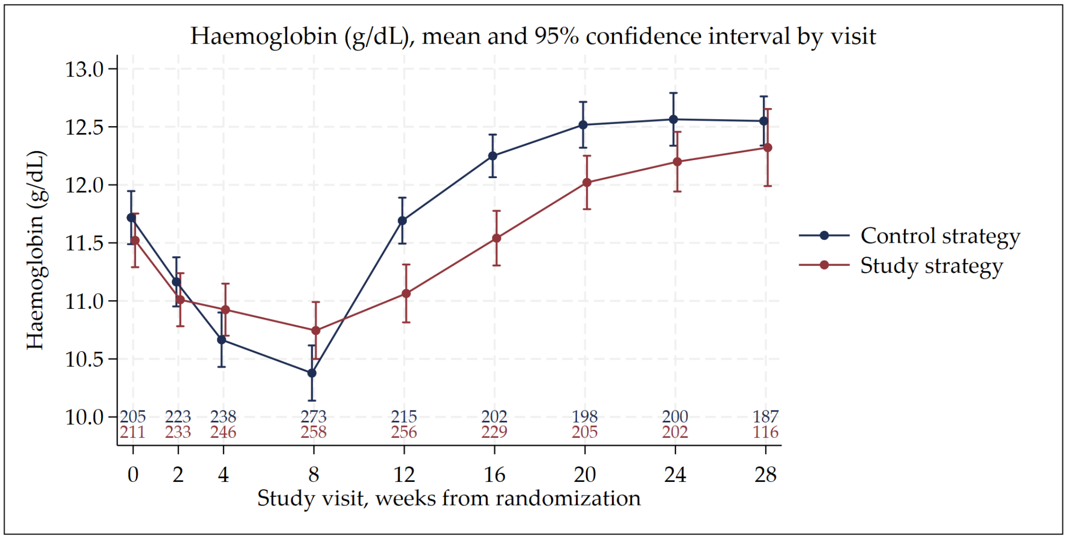

Figure S3.17: Kaplan-Meier of time to onset of anaemia.

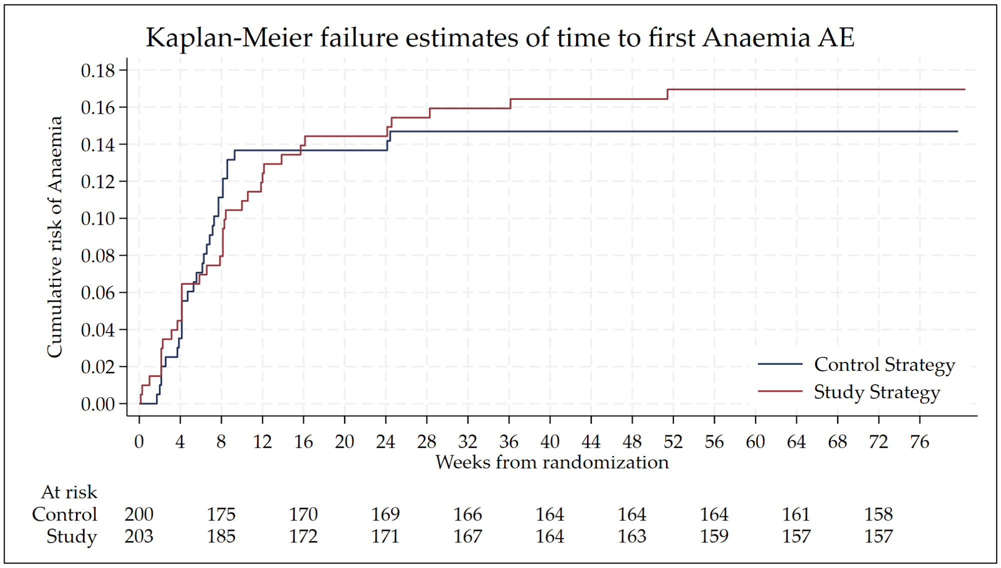

Figure S3.18: Mean neutrophils over time.

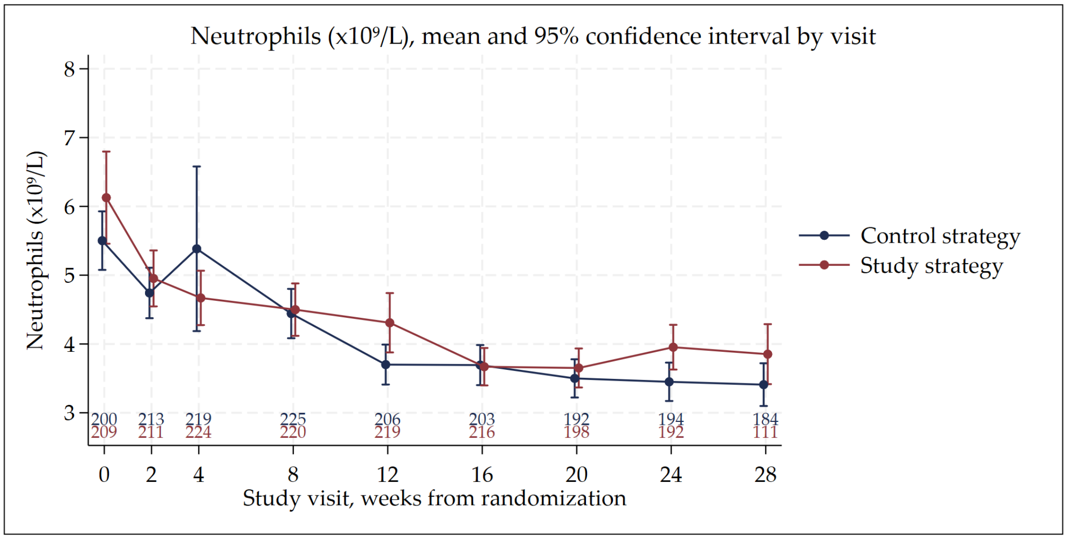

Figure S3.19: Mean platelets over time.

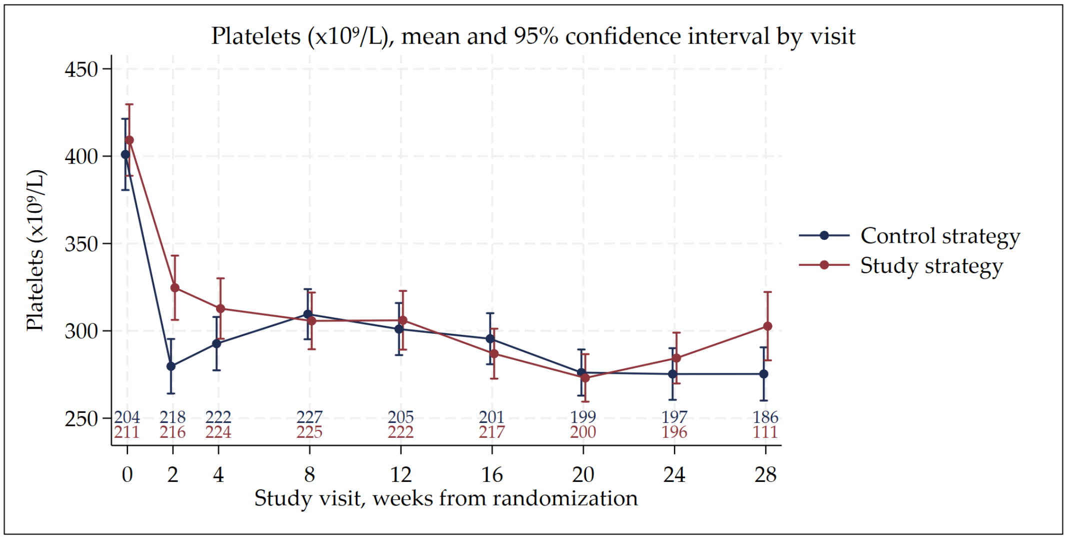

Figure S3.20: Mean potassium over time.

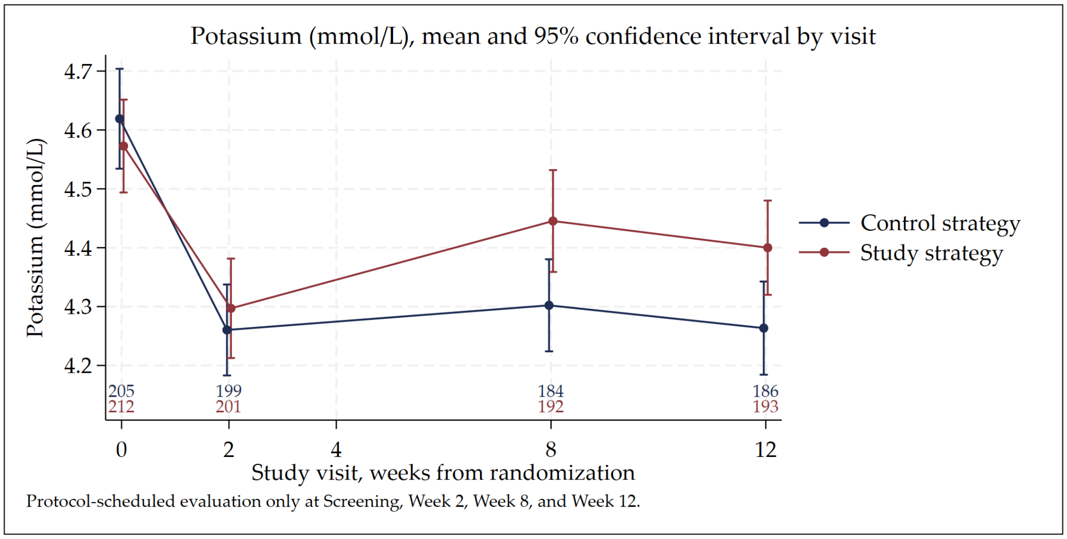

##### 3.6 ECG Results

Figure S3.21: Mean Fridericia-correct QT (QTcF) over time.

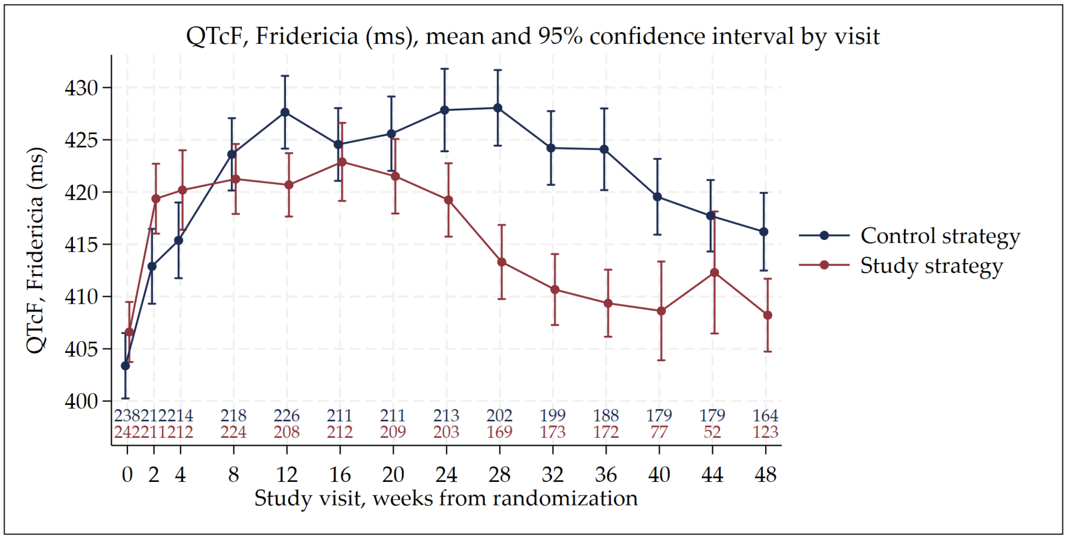

Figure S3.22: Mean change in Fridericia-correct QT (QTcF) from baseline over time.

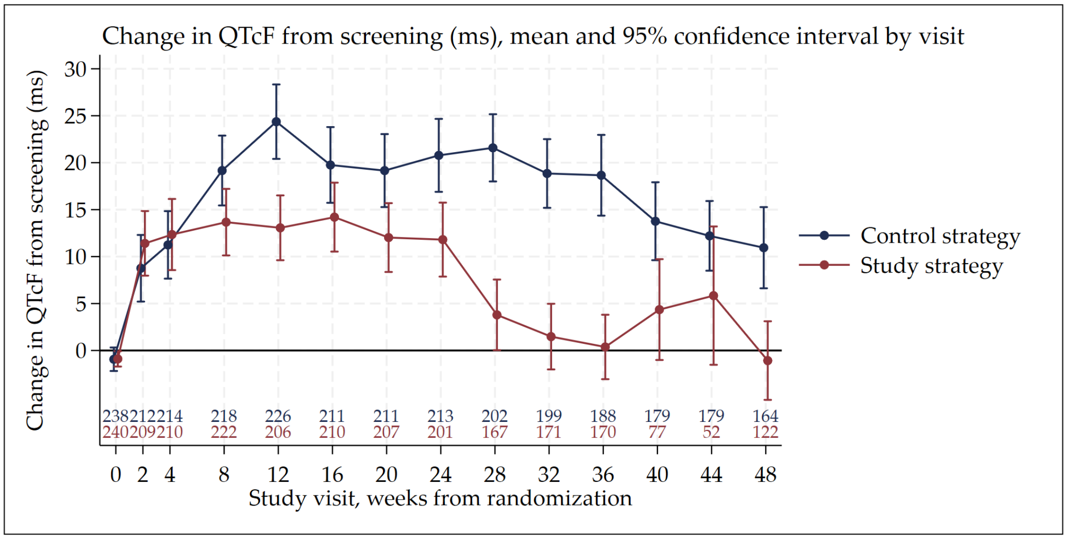

##### 3.7 Change in Weight

Figure S3.23: Mean change in weight from baseline over time.

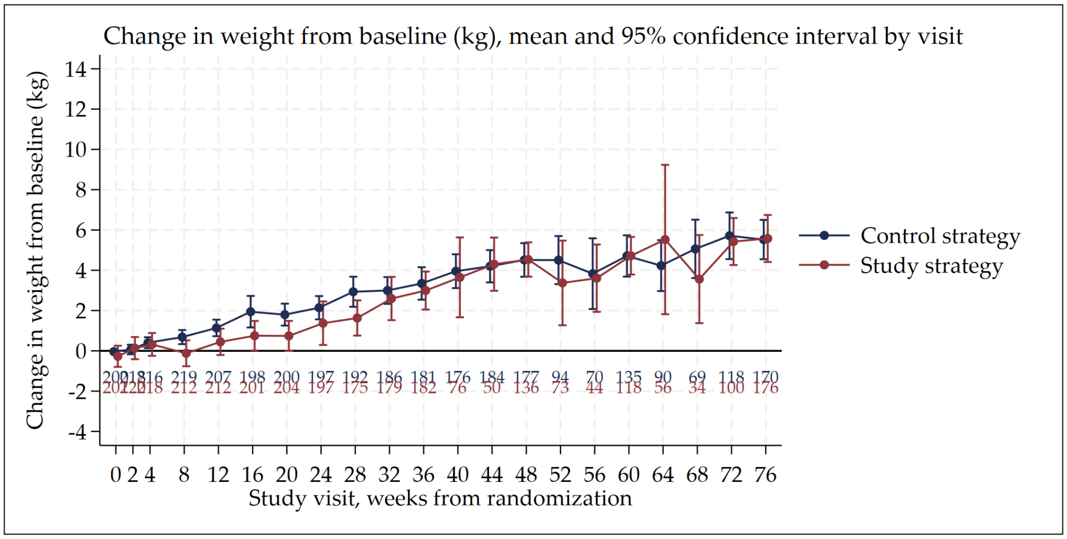

#### 3.8 Pregnancy

Table S3.18: Listing of all women that were pregnant at baseline or became pregnant during the study.

| | Arm | Outcome | Weeks from randomization and delivery |
|---|---|---|---|
| 1 | Study | Full term live birth | 6.3 |
| 2 | Control | Premature live birth | 8.9 |
| 3 | Control | Full term live birth | 12.0 |
| 4 | Control | Full term live birth | 12.9 |
| 5 | Study | Full term live birth | 17.6 |
| 6 | Control | Full term live birth | 18.7 |
| 7 | Study | Full term live birth | 19.4 |
| 8 | Control | Full term live birth | 26.0 |
| 9 | Control | Full term live birth | 37.4 |
| 10 | Study | Full term live birth | 37.9 |

Table S3.19: Primary week 76 efficacy outcome among pregnant women.

| Primary Outcome | Control Strategy | Study Strategy | Total |
|---|---|---|---|
| **Total randomized (ITT population)** | **6** | **4** | **10** |
| **Successful outcome at end of treatment and follow-up** | | | |
| **Total** | **6 (100.0%)** | **3 (75.0%)** | **9 (90.0%)** |
| Cured at end of treatment, and end of follow-up | 6 (100.0%) | 3 (75.0%) | 9 (90.0%) |
| **Unsuccessful end of follow-up** | | | |
| Total | 0 | 1 (25.0%) | 1 (10.0%) |
| Recurrence after cure at end of treatment* | 0 | 1 (25.0%) | 1 (10.0%) |

*Note:* One participant experienced recurrence 7 months after full term delivery of infant.

#### 3.9 Time to stable culture negative conversion

Figure S3.24: Kaplan Meier of time to stable culture conversion by strategy.

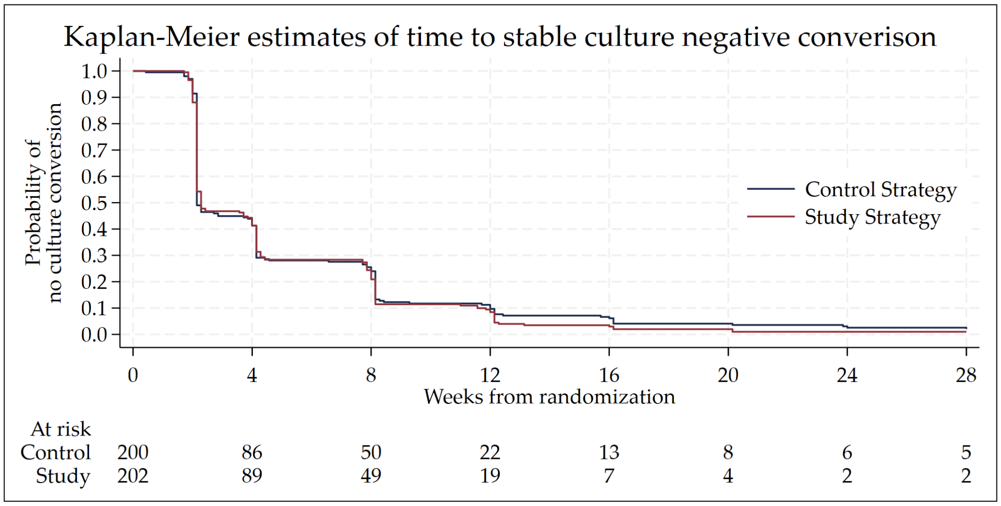

Figure S3.25: Kaplan Meier of time to stable culture conversion by strategy and fluoroquinlone sensitivity

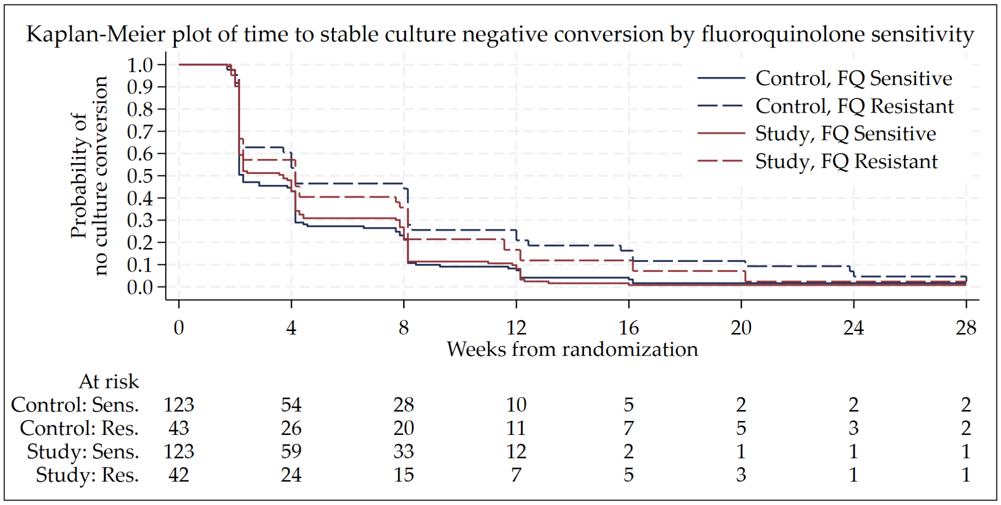

#### 3.10 Primary Efficacy

##### 3.10.1 Primary outcome: Combined end of treatment and end of follow-up outcome

Table S3.20: Summary of week 76 primary efficacy outcome.

| Primary Outcome | Control Strategy | Study Strategy | Total |
|---|---|---|---|
| **Total randomized (ITT population)** | **200** | **202** | **402** |
| **Successful outcome at end of treatment and follow-up** | | | |
| **Total** | **172 (86.0%)** | **174 (86.1%)** | **346 (86.1%)** |
| Cured at end of treatment, and end of follow-up | 162 (81.0%) | 160 (79.2%) | 322 (80.1%) |
| Cured at end of treatment, culture negative when last seen | 10 (5.0%) | 14 (6.9%) | 24 (6.0%) |
| **Unsuccessful end of treatment outcome** | | | |
| **Total** | **22 (11.0%)** | **14 (6.9%)** | **36 (9.0%)** |
| Treatment failed | 10 (5.0%) | 7 (3.5%) | 17 (4.2%) |
| Lost to follow-up on treatment | 4 (2.0%) | 2 (1.0%) | 6 (1.5%) |
| Died while on treatment | 7 (3.5%) | 4 (2.0%) | 11 (2.7%) |
| Not Evaluated (Participant withdrew consent) | 1 (0.5%) | 1 (<0.5%) | 2 (<0.5%) |
| **Unsuccessful end of follow-up** | | | |
| **Total** | **6 (3.0%)** | **14 (6.9%)** | **20 (5.0%)** |
| Recurrence after cure at end of treatment | 4 (2.0%) | 10 (5.0%) | 14 (3.5%) |
| Died after cure at end of treatment | 2 (1.0%) | 4 (2.0%) | 6 (1.5%) |

Table S3.21: Primary outcome: Risk difference

| | Risk difference* | p-value for non-inferiority test |
|---|---|---|
| Adjusted difference, HIV, site | -0.2% (-6.9%, 6.5%) | One-sided p = 0.0014 |
| Unadjusted difference | -0.1% (-6.9%, 6.6%) | One-sided p = 0.0017 |

* Compare upper bound with 10% margin of non-inferiority

Figure S3.26: Risk difference plot of the primary safety outcome for primary outcome

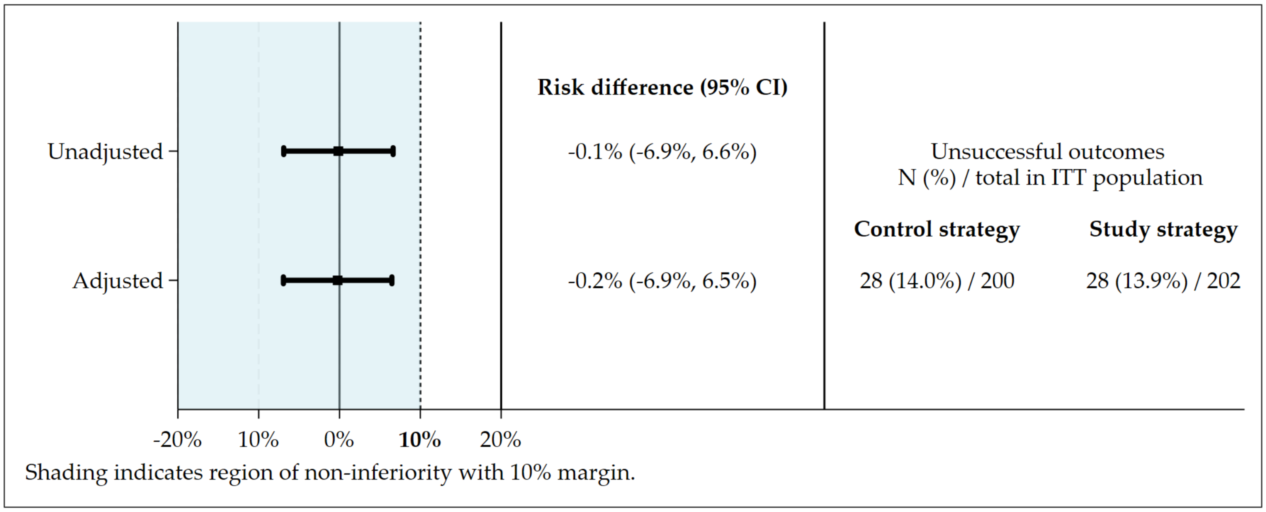

Table S3.22: Time of last negative culture from randomization.

| Parameter | Control Strategy | Study Strategy | Total |
|---|---|---|---|
| **Cured at end of treatment, culture negative when last seen** | | | |
| Median (IQR) | 71.5 (60.1, 72.0) | 66.4 (59.4, 72.6) | 71.5 (59.8, 72.1) |
| Min, Max | 24.0, 72.1 | 36.1, 75.4 | 24.0, 75.4 |
| Number | 10 | 14 | 24 |

#### 3.11 Change in resistance profile during and after treatment

There were a total of 14 recurrences and 17 treatment failures. Among these 31, listings below show baseline drug susceptibility and any changes post-randomization.

BDQ = Bedaquiline, CLZ = Clofazimine, Ethion = Ethionamide, FQ = Any fluoroquinolone, INH = Isoniazid, Inj = Any injectable aminoglycoside, LZD = Linezolid, PAS = Para-aminosalicylic acid.

Table S3.23: Changes in drug resistance: Participants with FQ-sensitive disease at baseline.

| | Strategy | Baseline resistance | Baseline sensitivity | New resistance post-randomization | New sensitivity post-randomization |
|---|---|---|---|---|---|
| **Recurrence after cure at end of treatment** | | | | | |
| 1 | Control | | FQ | | LZD, PAS, CLZ, Ethion, BDQ |
| 2 | Control | | INH, FQ, Inj | | Ethion |
| 3 | Control | | FQ | No post-baseline data | No post-baseline data |
| 4 | Study | | INH, FQ | | Inj |
| 5 | Study | INH | FQ, Inj | | BDQ, CLZ, Ethion, LZD, PAS |
| 6 | Study | INH | FQ | Ethion | BDQ, CLZ, PAS, LZD |
| 7 | Study | | INH, FQ, Inj | | BDQ, LZD |
| **Treatment failed** | | | | | |
| 8 | Control | | INH, FQ | | Inj |
| 9 | Control | INH | FQ, Inj | FQ, BDQ, Inj | CLZ, LZD, PAS |
| 10 | Control | INH | FQ, Inj | Ethion | CLZ, PAS, LZD, BDQ |
| 11 | Control | INH | FQ, Inj | FQ | BDQ, CLZ, LZD |
| 12 | Control | INH | FQ, Inj | | Ethion, PAS, INH, LZD, CLZ, BDQ |
| 13 | Study | INH | FQ, Inj | Ethion | BDQ, CLZ, LZD |
| 14 | Study | INH | FQ, Inj | | PAS, BDQ, Ethion, CLZ, LZD |
| 15 | Study | | INH, FQ, Inj | INH | BDQ, LZD |

Table S3.24: Changes in drug resistance: Participants with FQ-resistant disease at baseline.

| | Strategy | Baseline resistance | Baseline sensitivity | New resistance post-randomization | New sensitivity post-randomization |
|---|---|---|---|---|---|
| **Treatment failed** | | | | | |
| 16 | Control | INH, FQ, Inj | None | BDQ, Ethion | LZD, FQ, PAS |
| 17 | Control | INH, FQ, Inj | LZD, BDQ, CLZ | No post-baseline data | No post-baseline data |
| 18 | Study | INH, FQ, Inj | LZD, BDQ, CLZ | No post-baseline data | No post-baseline data |
| 19 | Study | INH, FQ, Inj | LZD, BDQ | No post-baseline data | No post-baseline data |
| 20 | Study | INH, FQ, Inj | LZD, BDQ, CLZ | Ethion | |
| 21 | Study | INH, FQ, Inj | LZD, BDQ, CLZ | No post-baseline data | No post-baseline data |
| 22 | Study | INH, FQ, Inj | LZD, BDQ, CLZ | Ethion, LZD, BDQ, CLZ | PAS |
| 23 | Study | INH, FQ, Inj | LZD, BDQ | BDQ, Ethion, CLZ | PAS |
| **Recurrence after cure at end of treatment** | | | | | |
| 24 | Control | INH, FQ, Inj | LZD, BDQ, CLZ | CLZ, Ethion, BDQ | PAS |
| 25 | Control | INH, FQ, Inj | LZD, BDQ, CLZ | Ethion, BDQ, CLZ | FQ, PAS |
| 26 | Control | INH, FQ | LZD, BDQ, CLZ, Inj | Ethion, CLZ, BDQ | PAS, Ethion |
| 27 | Study | INH, FQ | LZD, BDQ, CLZ, Inj | BDQ, Ethion, CLZ | PAS, FQ |
| 28 | Study | INH, FQ, Inj | LZD, BDQ, CLZ | CLZ, BDQ | |
| 29 | Study | INH, FQ, Inj | LZD, BDQ, CLZ | BDQ, Ethion, CLZ | PAS |
| 30 | Control | INH, FQ, BDQ, Inj | LZD | Ethion | FQ, PAS |

Table S3.25: Changes in drug resistance: Participants with missing FQ Resistance DST at baseline.

| | Strategy | Baseline resistance | Baseline sensitivity | New resistance post-randomization | New sensitivity post-randomization |
|---|---|---|---|---|---|
| **Treatment failed** | | | | | |
| 31 | Study | | None | INH | FQ, BDQ, LZD, Inj |

##### 3.12 Subgroup Analyses of the Primary Efficacy and Safety Outcomes

Extensive subgroup analyses are provided for information only; they are not intended to be used for interference and should be interpreted with considerable caution.

A p-value for an interaction test is presented to show where there might be heterogeneity of treatment effect. Please note that none of these p-values have been adjusted for multiplicity. Where there are no unsuccessful outcomes in a treatment arm within a subgroup, an interaction test is not possible and the p-value is given as NA.

Many of these analyses were not pre-specified and the number of subgroup analyses presented means that a low interaction p-value may be a chance finding and not indicative of a true underlying interaction.

For more information on how to interpret sub-group analyses with appropriate caution, see:

- Wang R, Lagakos SW, Ware JH, Hunter DJ, Drazen JM. Statistics in medicine–reporting of subgroup analyses in clinical trials. N Engl J Med. 2007;357(21):2189-94.
- https://www.nejm.org/doi/full/10.1056/NEJMsr077003

##### 3.12.1 Study Site

Figure S3.27: Subgroup by study site: Non-inferiority plot of the primary efficacy outcome.

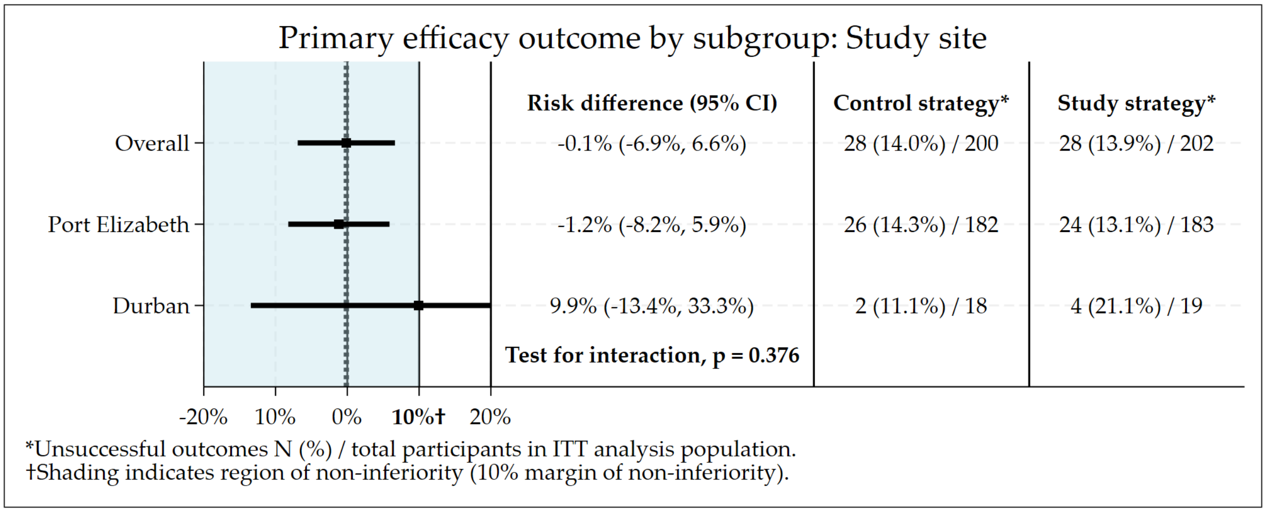

Figure S3.28: Subgroup by study site: Risk difference plot of the primary safety outcome.

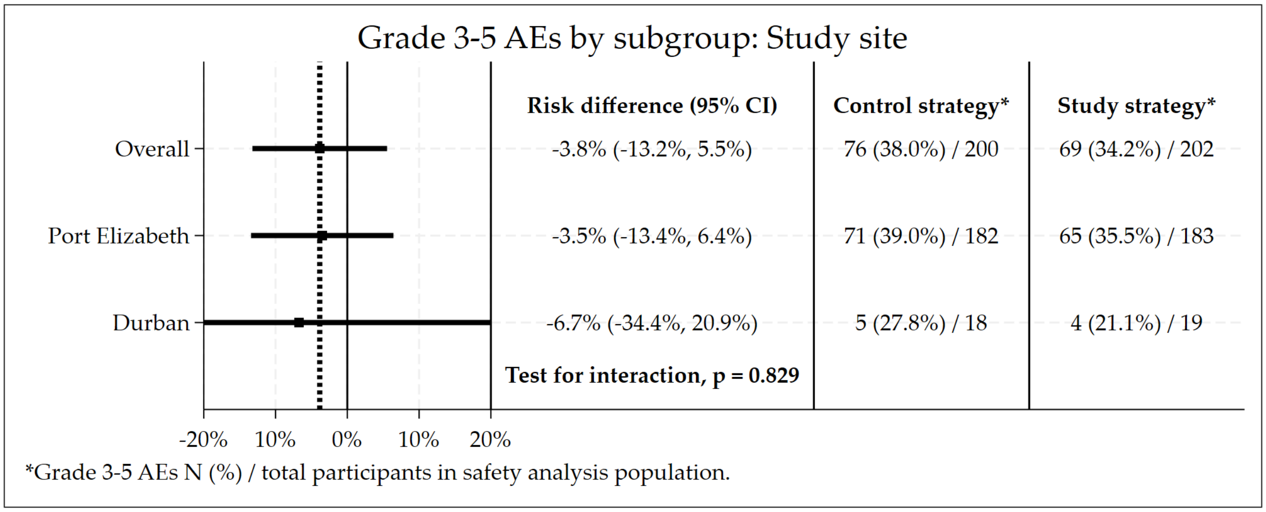

##### 3.12.2 Gender

Figure S3.29: Subgroup by gender: Non-inferiority plot of the primary efficacy outcome.

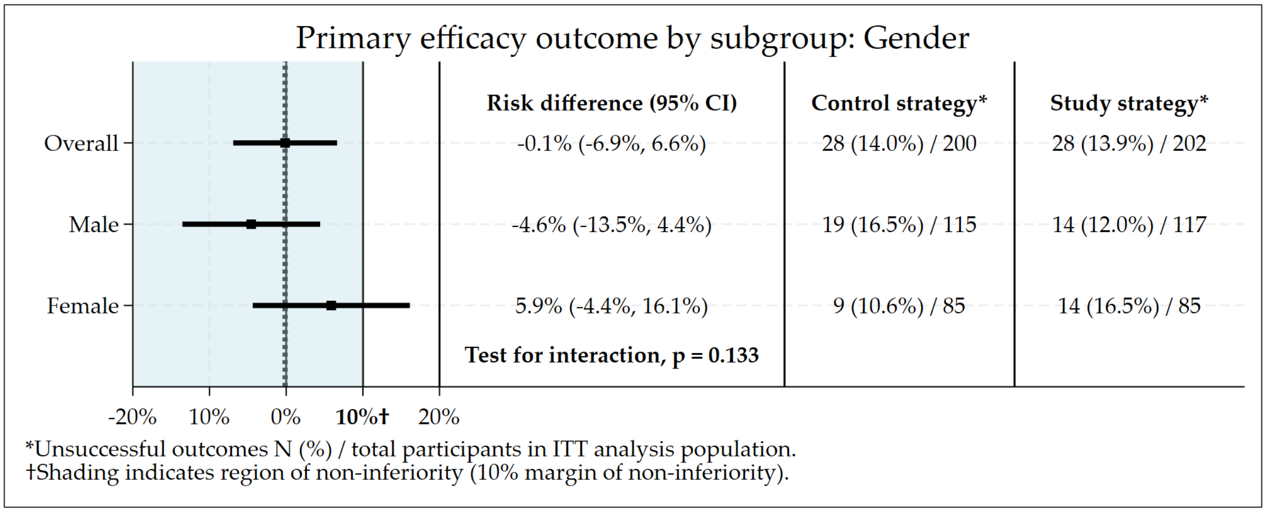

Figure S3.30: Subgroup by gender: Risk difference plot of the primary safety outcome.

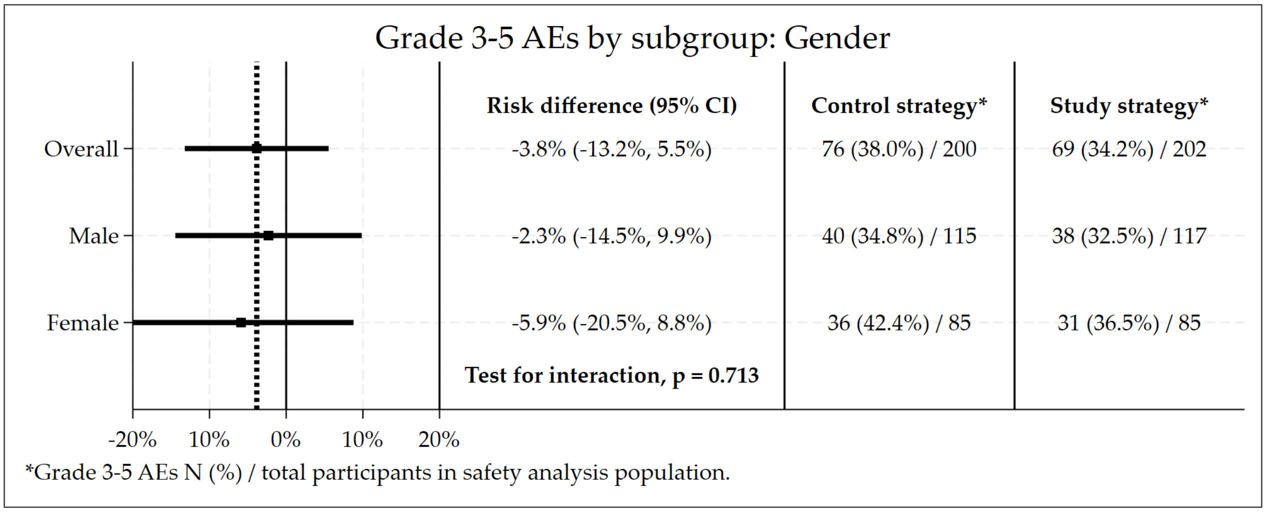

##### 3.12.3 Pregnancy

Figure S3.31: Subgroup by pregnancy status: Non-inferiority plot of the primary efficacy outcome.

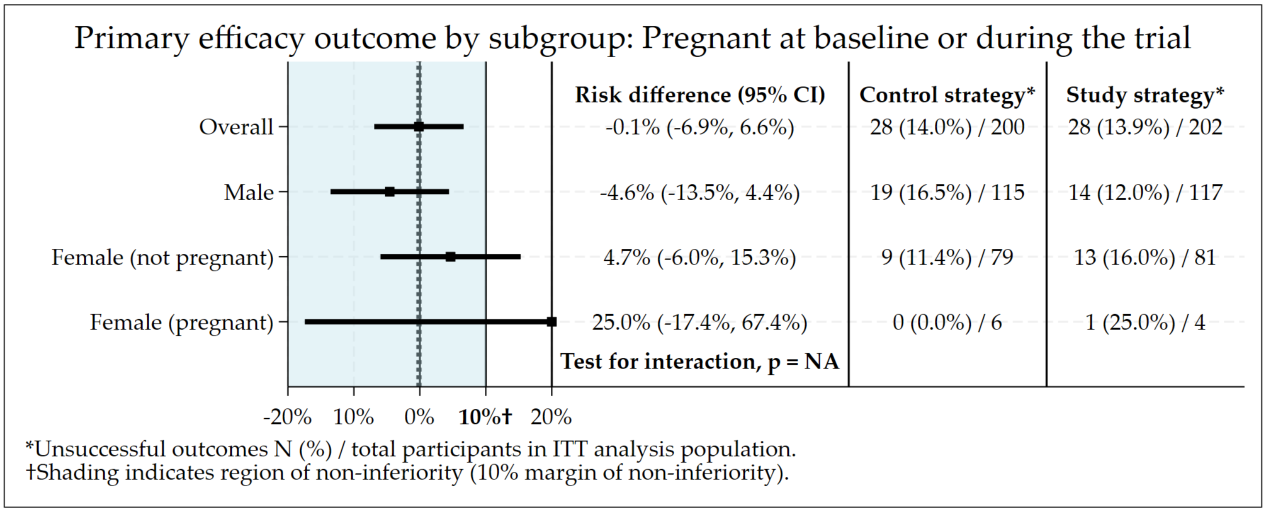

Figure S3.32: Subgroup by pregnancy status: Risk difference plot of the primary safety outcome.

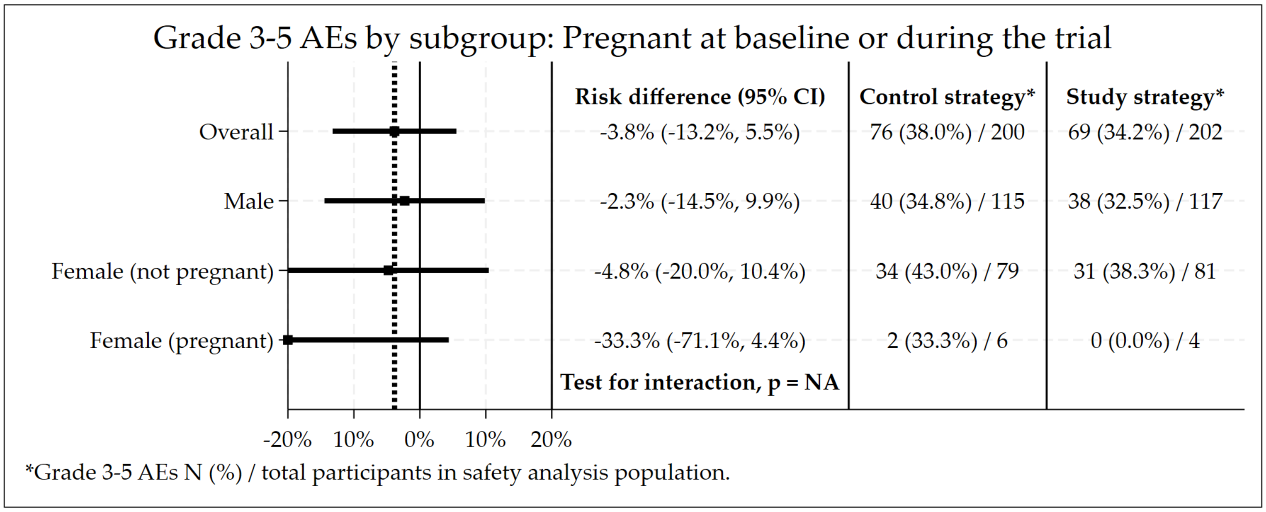

##### 3.12.4 Age at Baseline

Figure S3.33: Subgroup by age (years): Non-inferiority plot of the primary efficacy outcome.

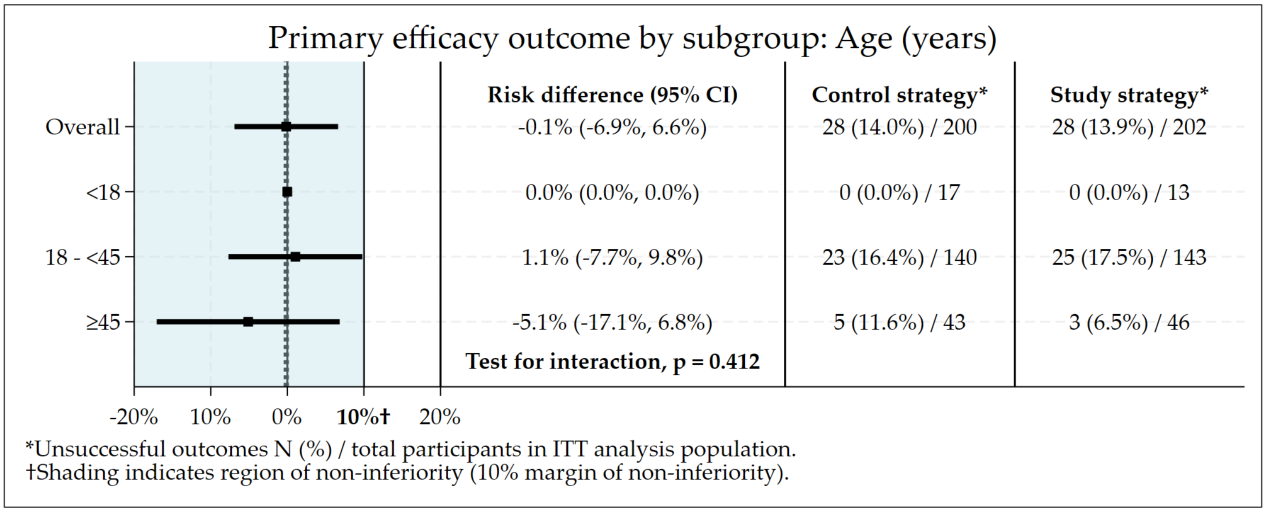

Figure S3.34: Subgroup by age (years): Risk difference plot of the primary safety outcome.

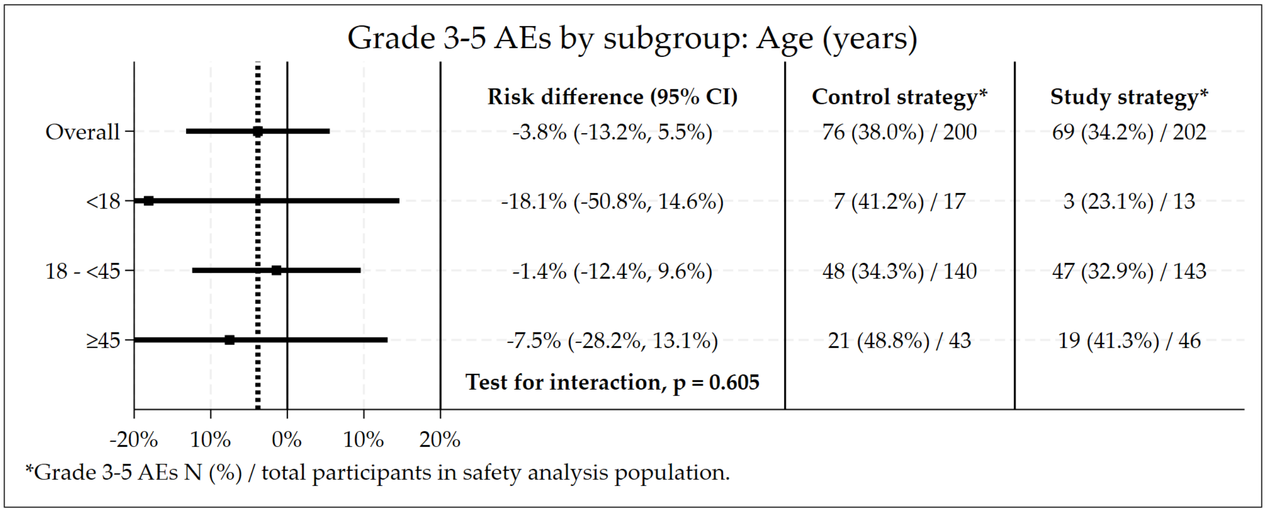

##### 3.12.5 BMI

Figure S3.35: Subgroup by BMI (kg/$m^2$): Non-inferiority plot of the primary efficacy outcome.

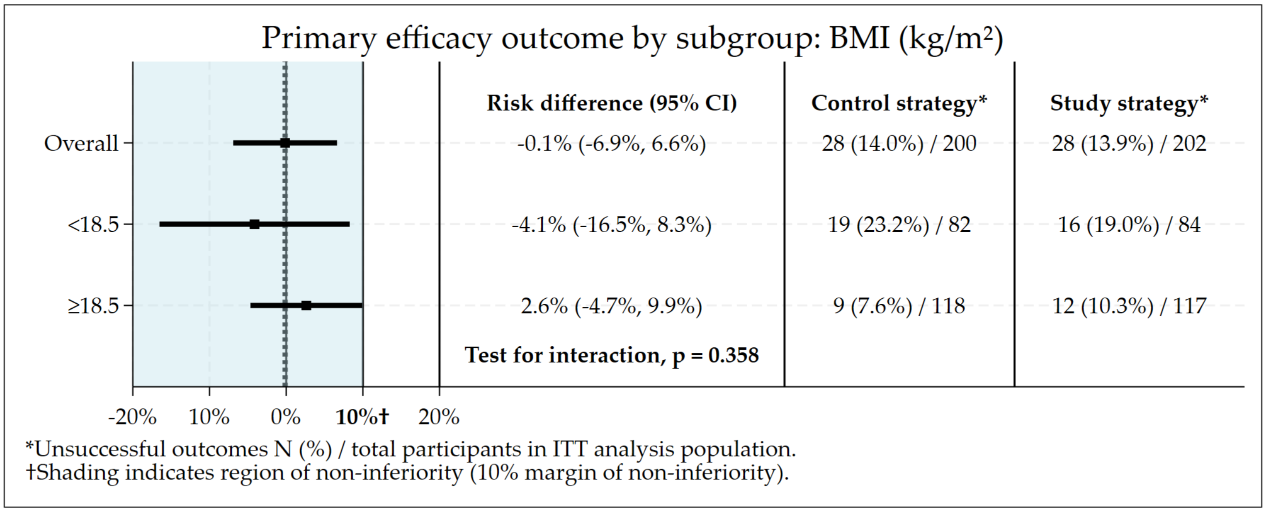

Figure S3.36: Subgroup by BMI (kg/$m^2$): Risk difference plot of the primary safety outcome.

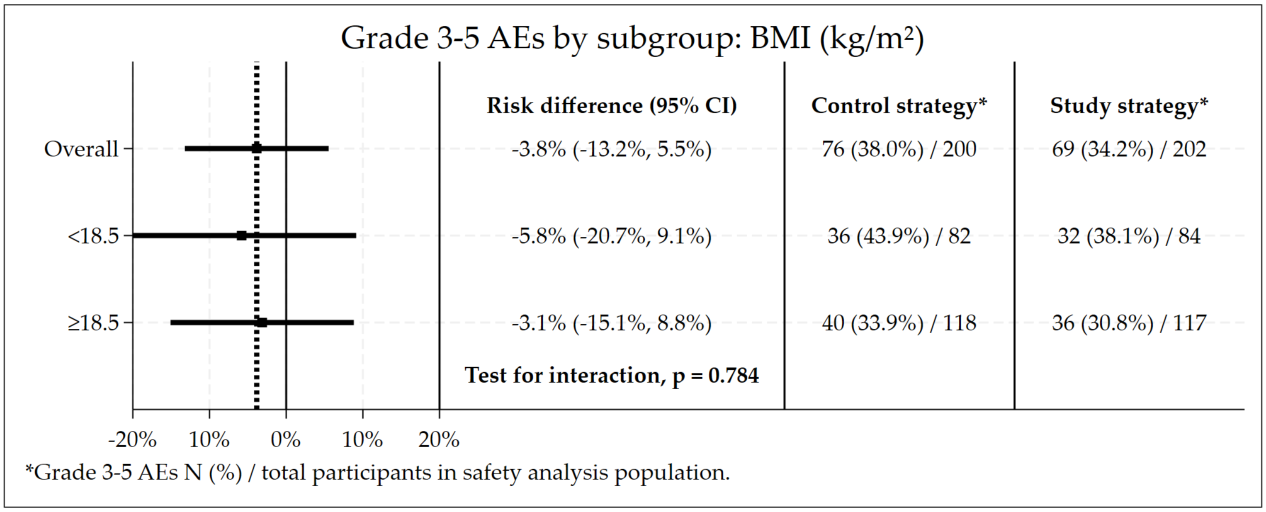

##### 3.12.6 Weight

Figure S3.37: Subgroup by Weight (kg): Non-inferiority plot of the primary efficacy outcome.

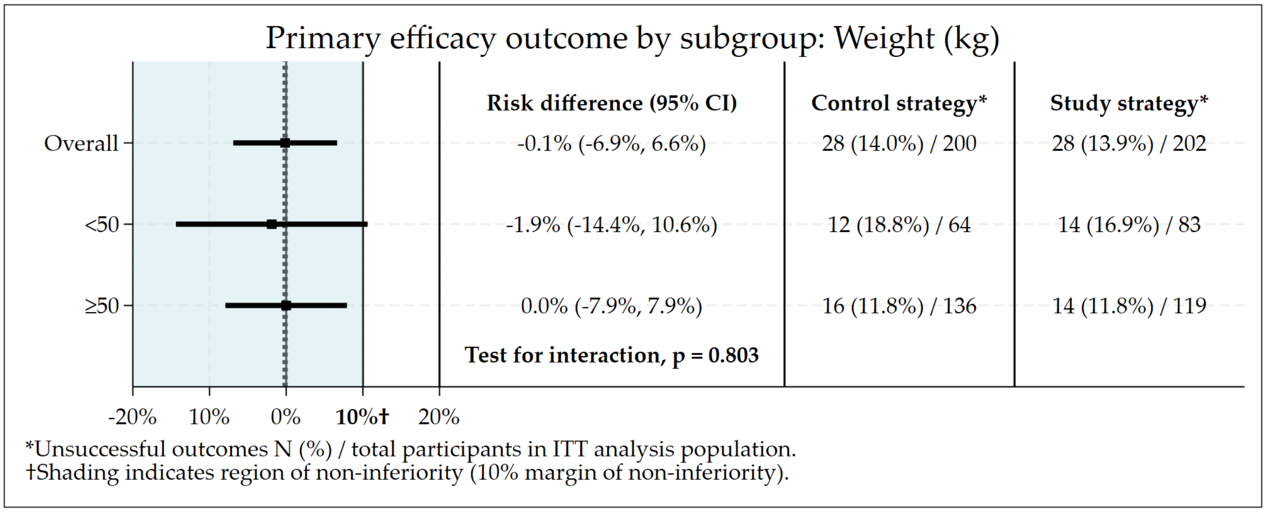

Figure S3.38: Subgroup by Weight (kg): Risk difference plot of the primary safety outcome.

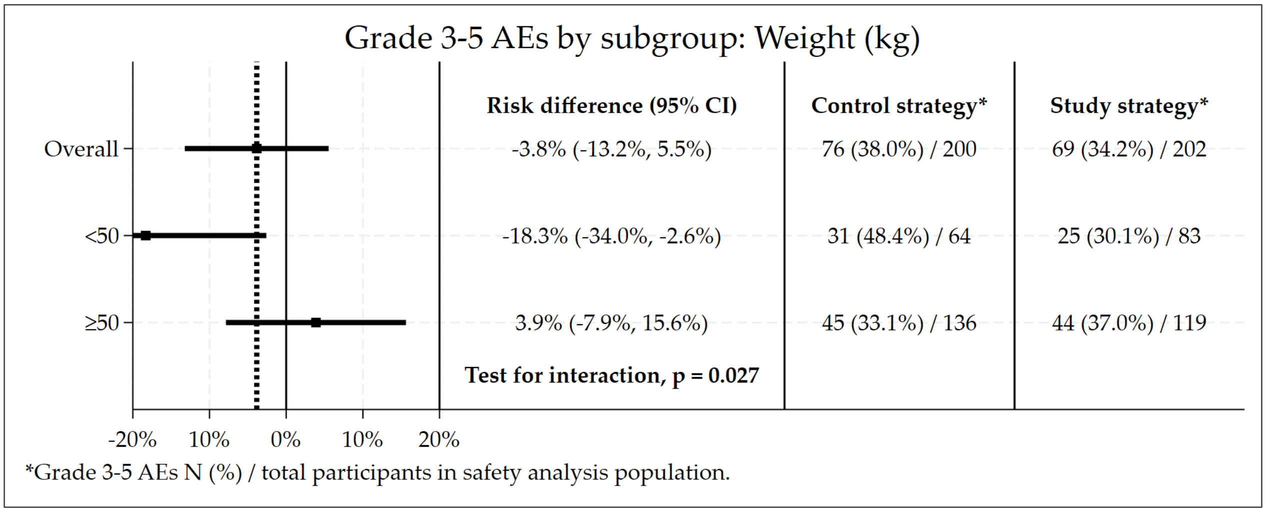

##### 3.12.7 Highest Level of Education

Figure S3.39: Subgroup by education: Non-inferiority plot of the primary efficacy outcome.

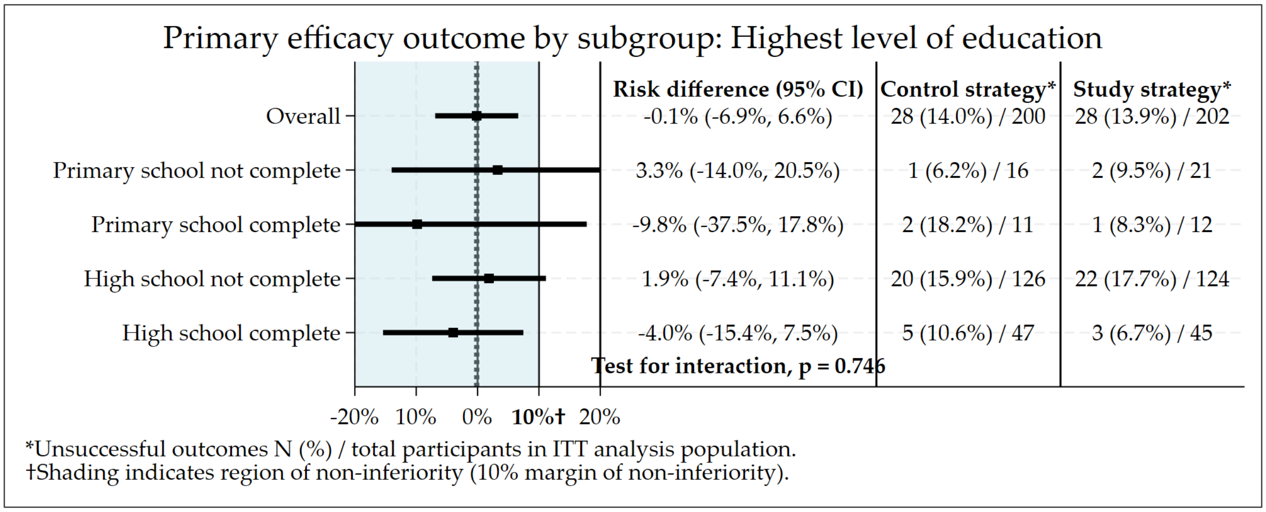

Figure S3.40: Subgroup by education: Risk difference plot of the primary safety outcome.

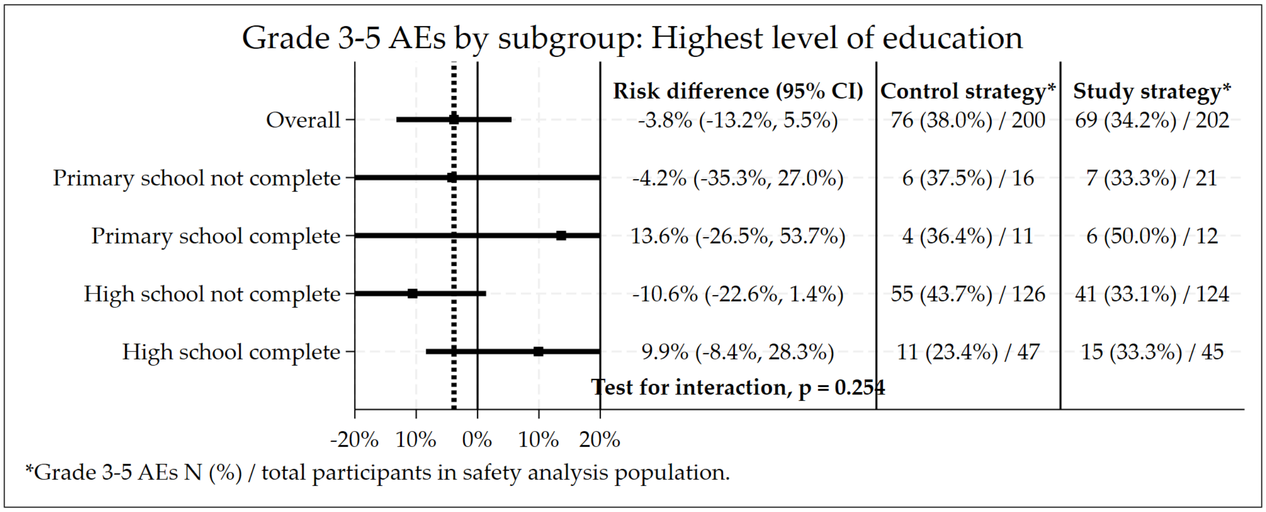

##### 3.12.8 Race

Figure S3.41: Subgroup by race: Non-inferiority plot of the primary efficacy outcome.

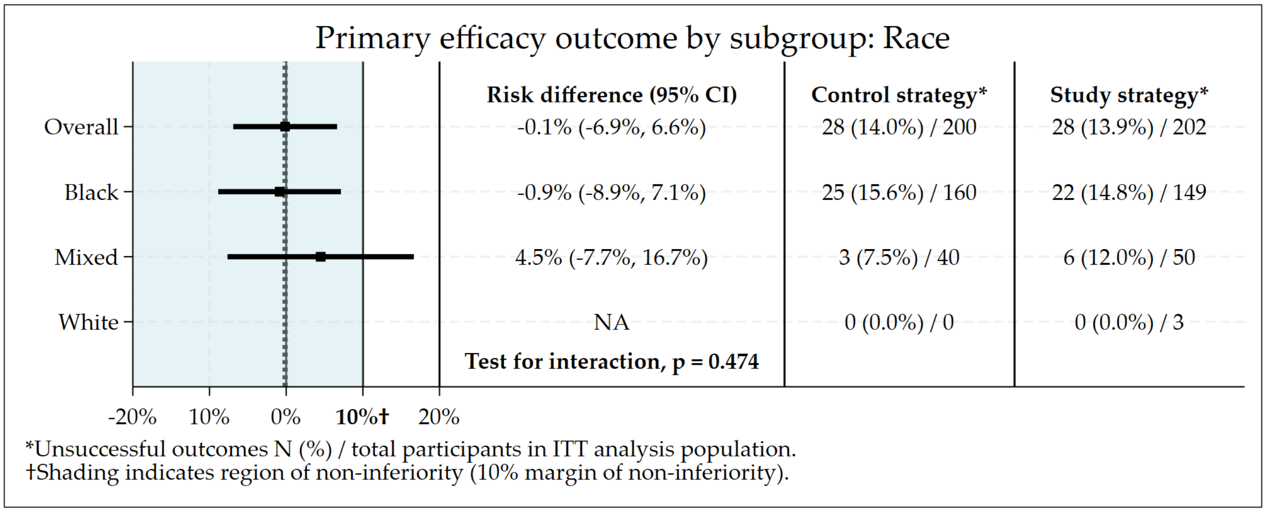

Figure S3.42: Subgroup by race: Risk difference plot of the primary safety outcome.

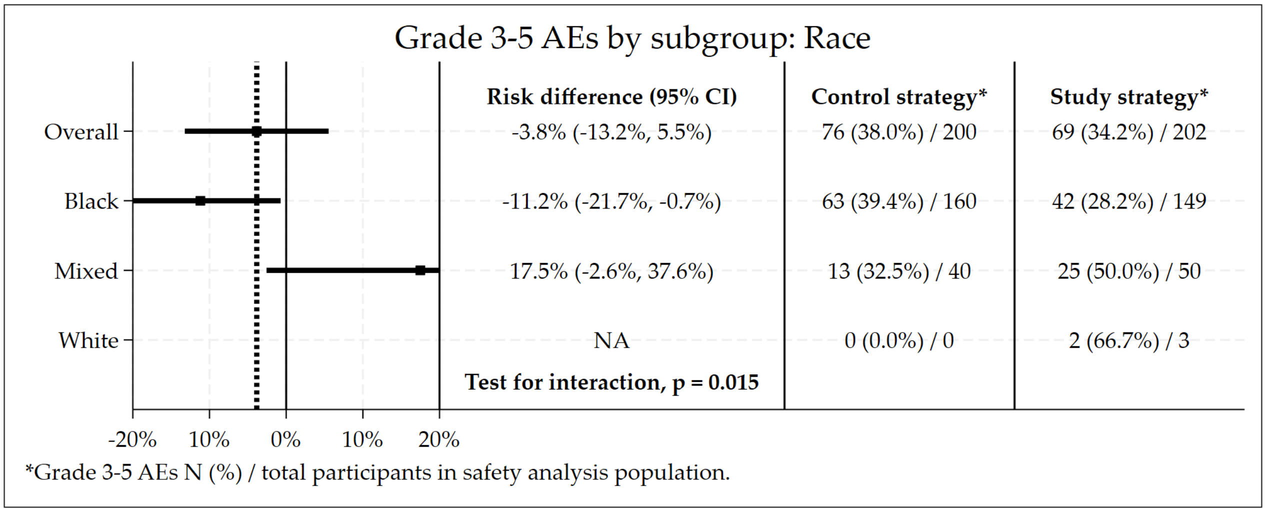

##### 3.12.9 HIV Status

Figure S3.43: Subgroup by HIV status: Non-inferiority plot of the primary efficacy outcome.

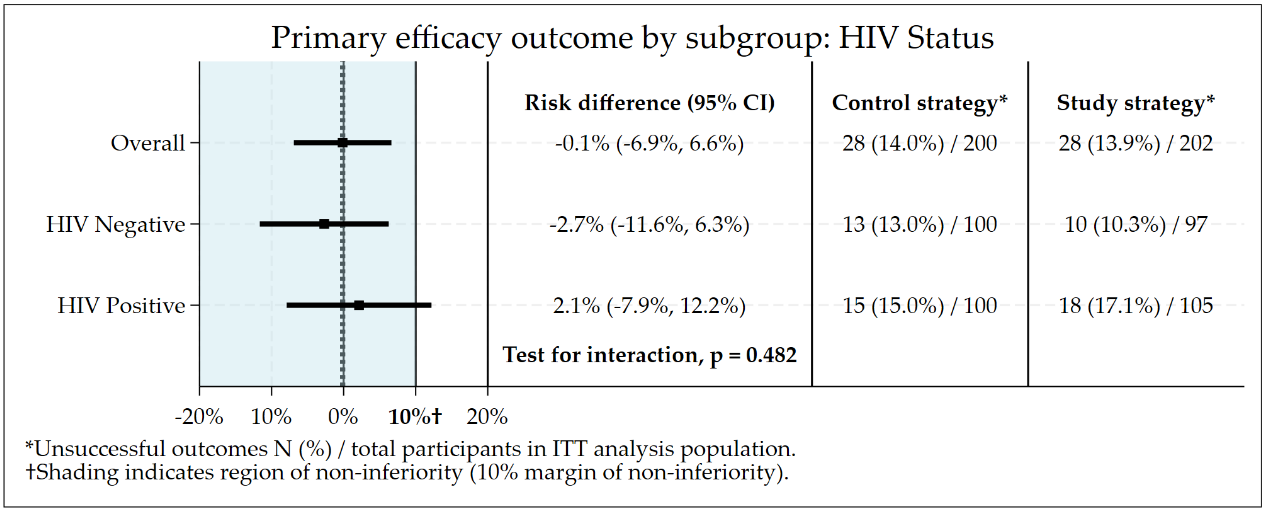

Figure S3.44: Subgroup by HIV status: Risk difference plot of the primary safety outcome.

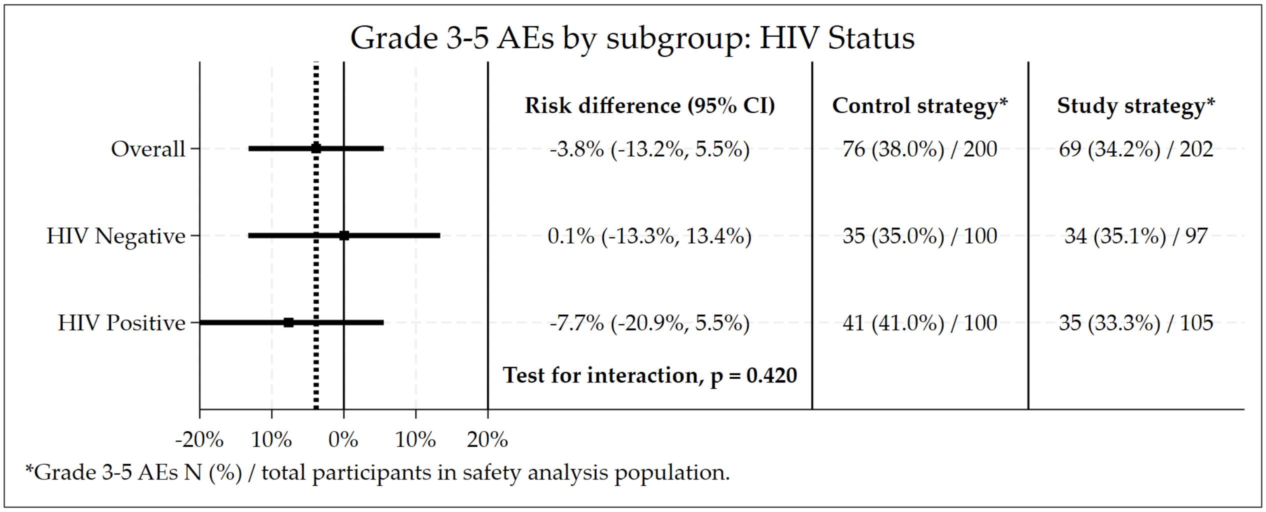

##### 3.12.10 CD4 Count

Figure S3.45: Subgroup by CD4 count (cells/mm$^3$): Non-inferiority plot of the primary efficacy outcome.

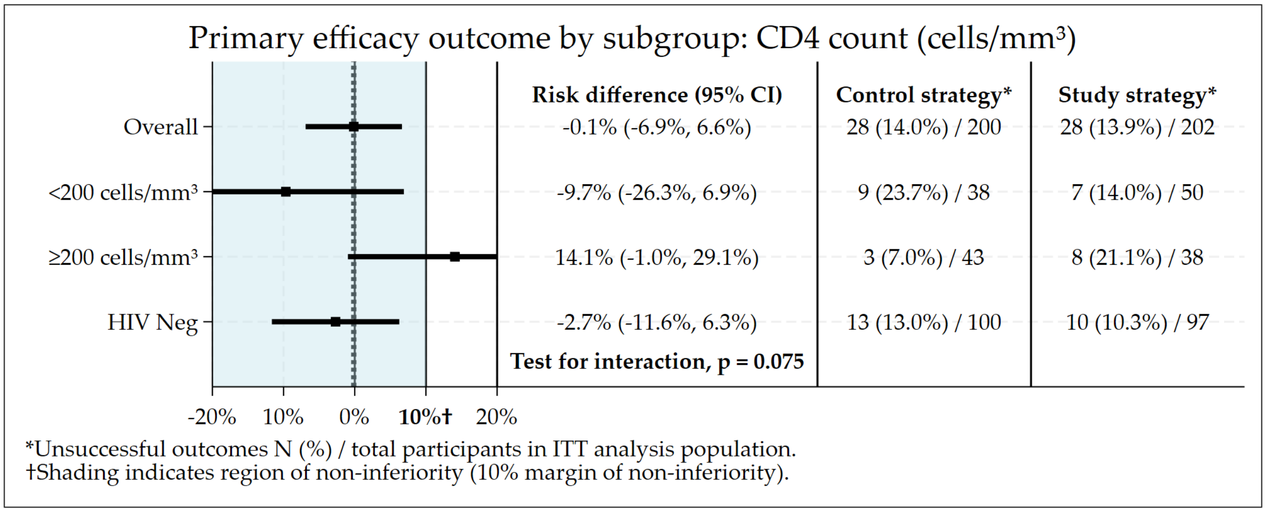

Figure S3.46: Subgroup by CD4 count (cells/mm$^3$): Risk difference plot of the primary safety outcome.

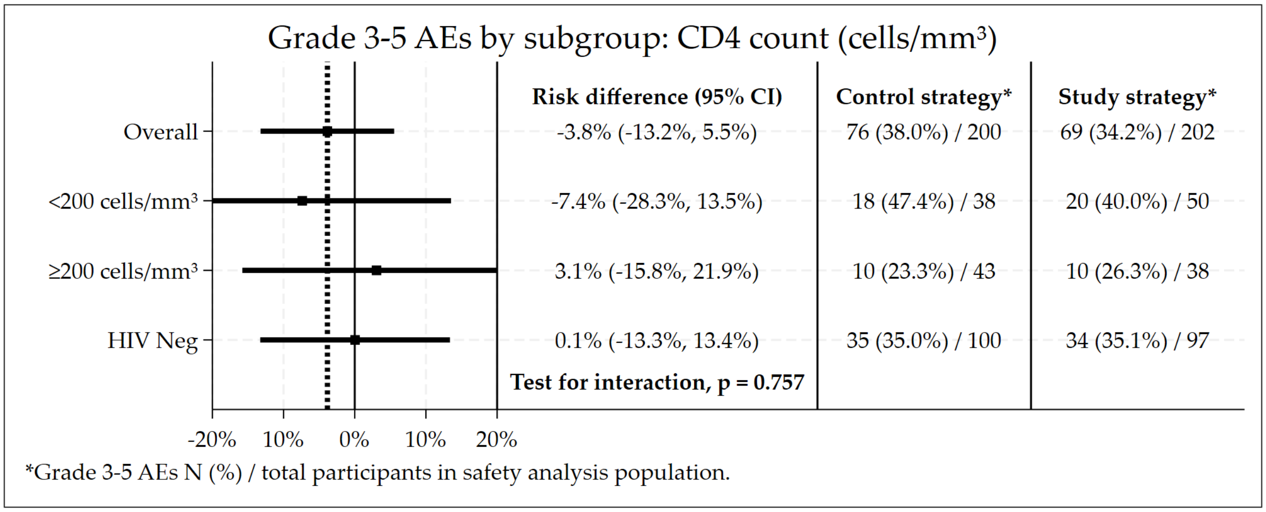

##### 3.12.11 HIV Viral Load

Figure S3.47: Subgroup by HIV Viral Load (copies/mm$^3$): Non-inferiority plot of the primary efficacy outcome.

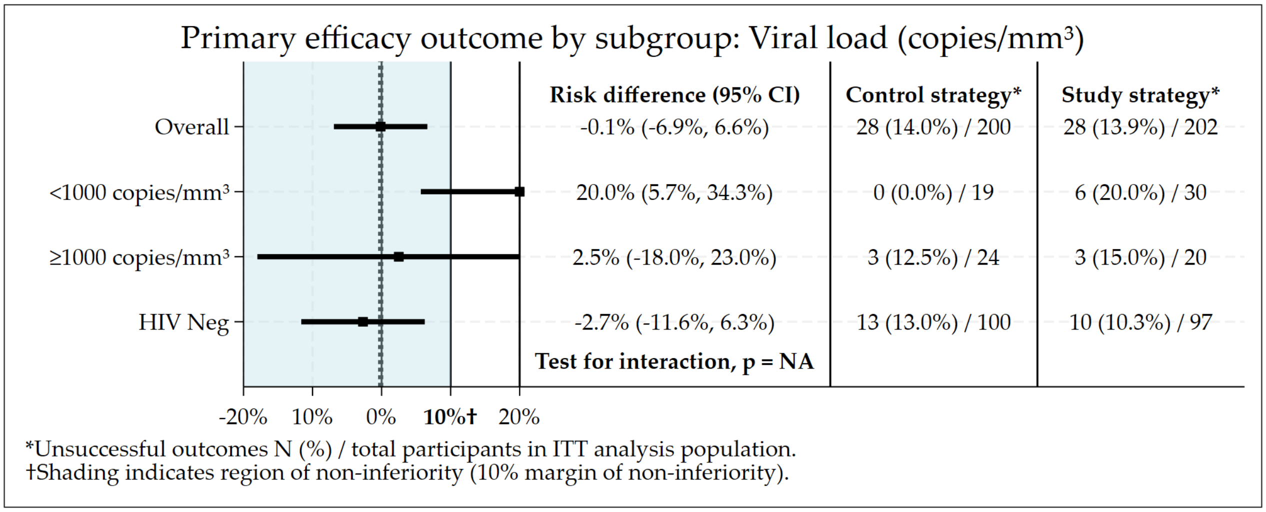

Figure S3.48: Subgroup by HIV Viral Load (copies/mm$^3$): Risk difference plot of the primary safety outcome.

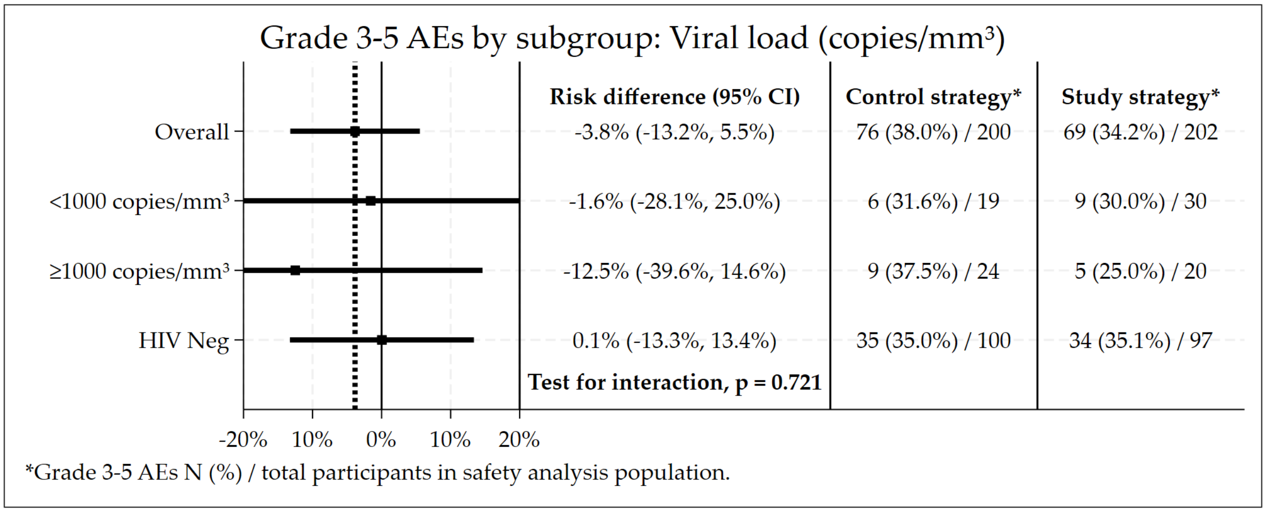

##### 3.12.12 Previous Diagnosis of TB

Figure S3.49: Subgroup by previous diagnosis of TB: Non-inferiority plot of the primary efficacy outcome.

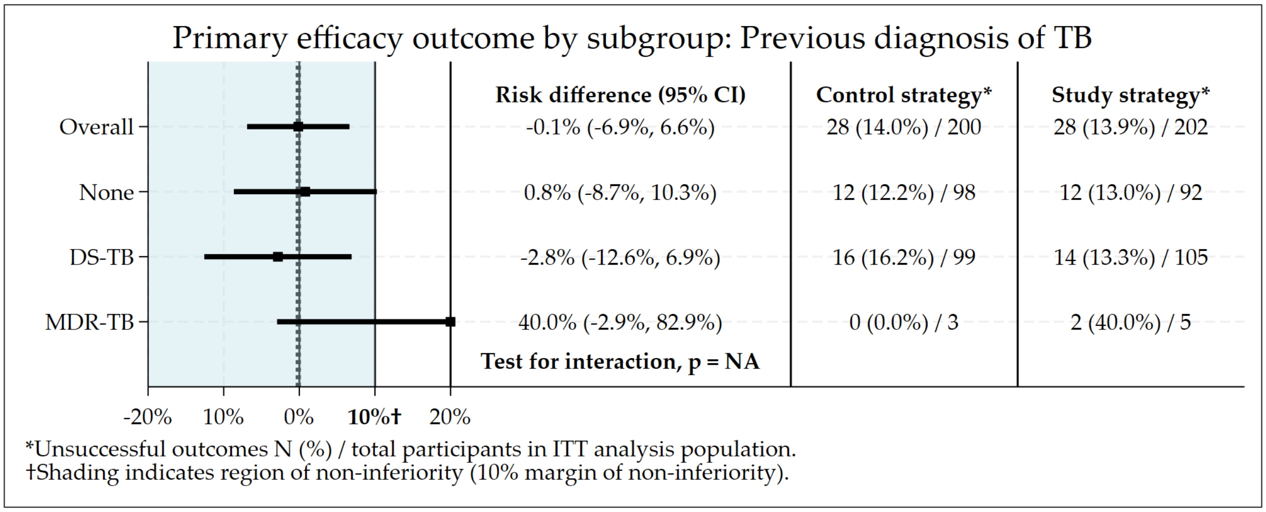

Figure S3.50: Subgroup by previous diagnosis of TB: Risk difference plot of the primary safety outcome.

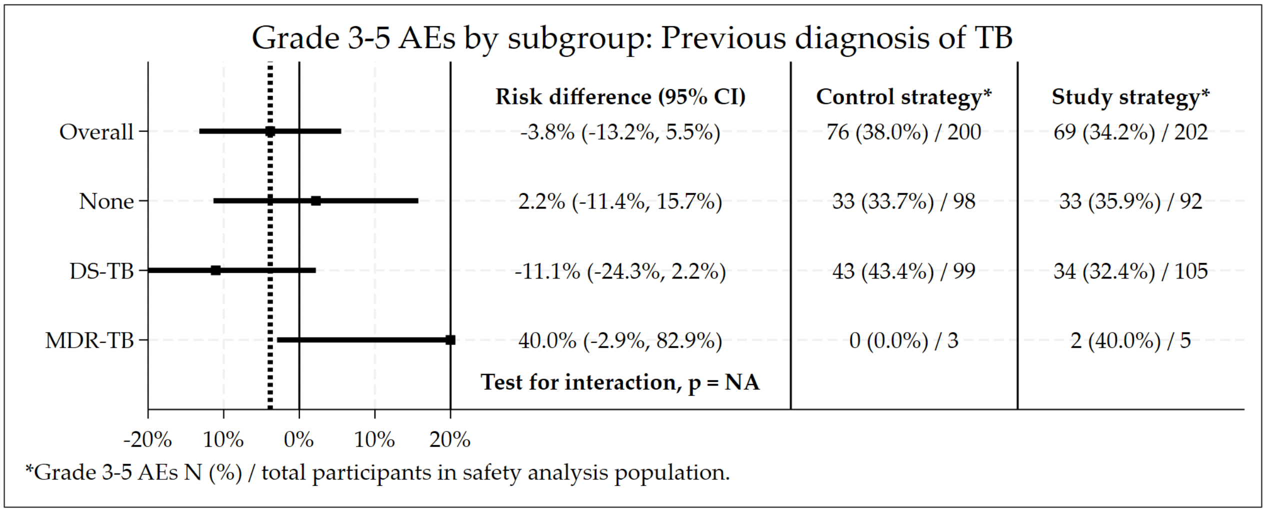

##### 3.12.13 MGIT Culture Result at Baseline

Figure S3.51: Subgroup by MGIT culture at baseline: Non-inferiority plot of the primary efficacy outcome.

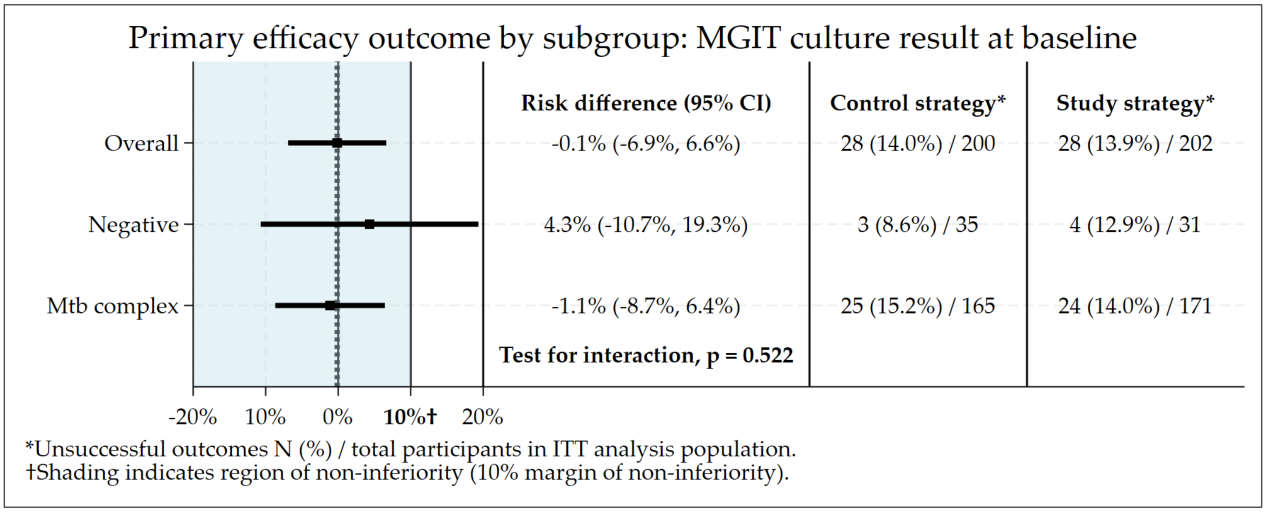

Figure S3.52: Subgroup by MGIT culture at baseline: Risk difference plot of the primary safety outcome.

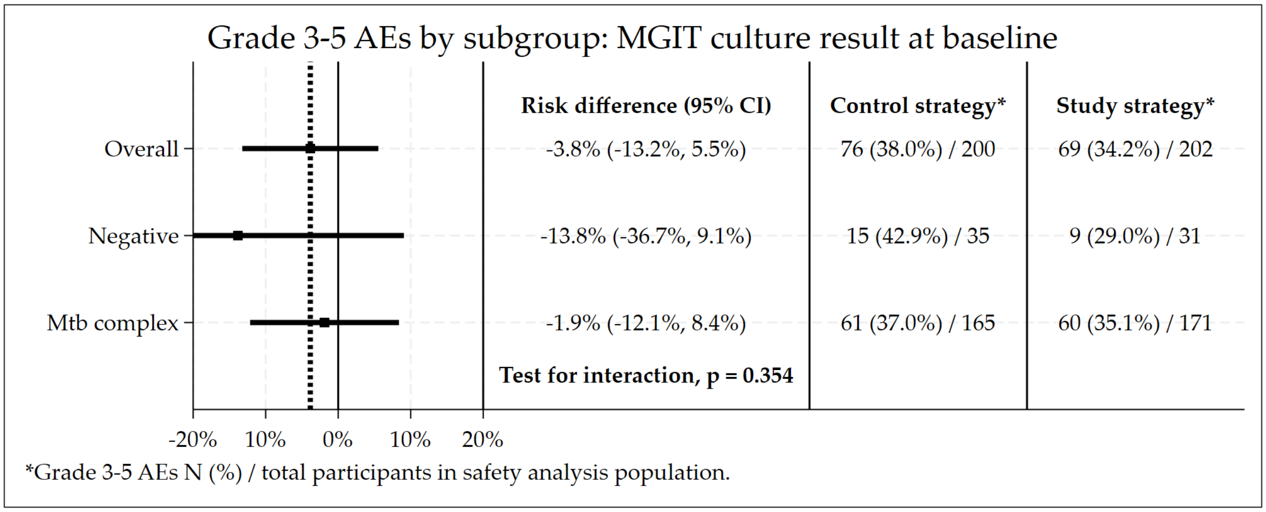

##### 3.12.14 Smear Grading at Baseline

Figure S3.53: Subgroup by smear grading at baseline: Non-inferiority plot of the primary efficacy outcome.

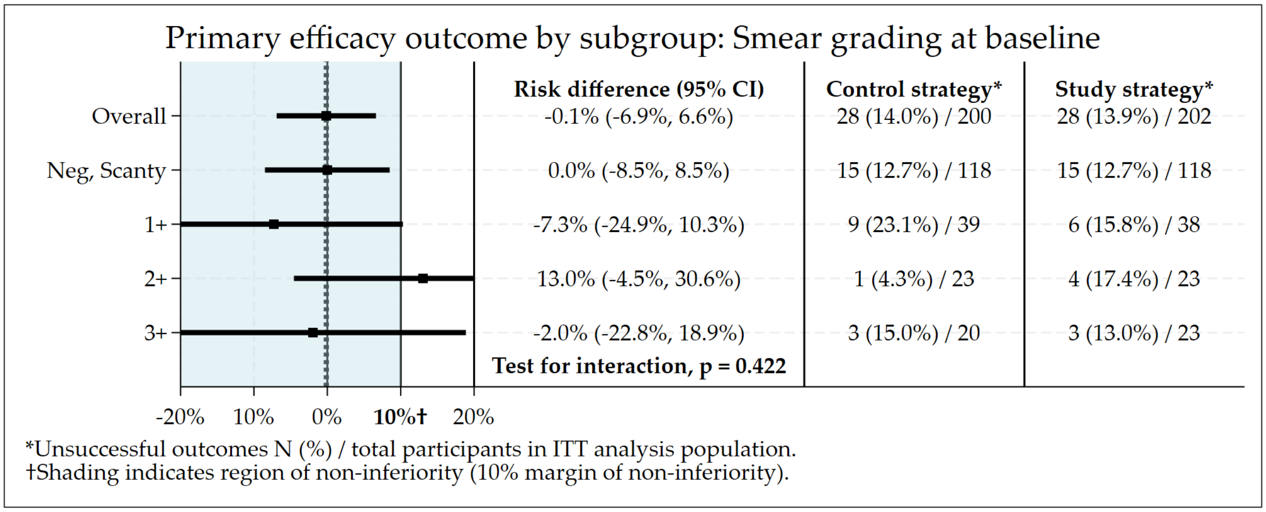

Figure S3.54: Subgroup by smear grading at baseline: Risk difference plot of the primary safety outcome.

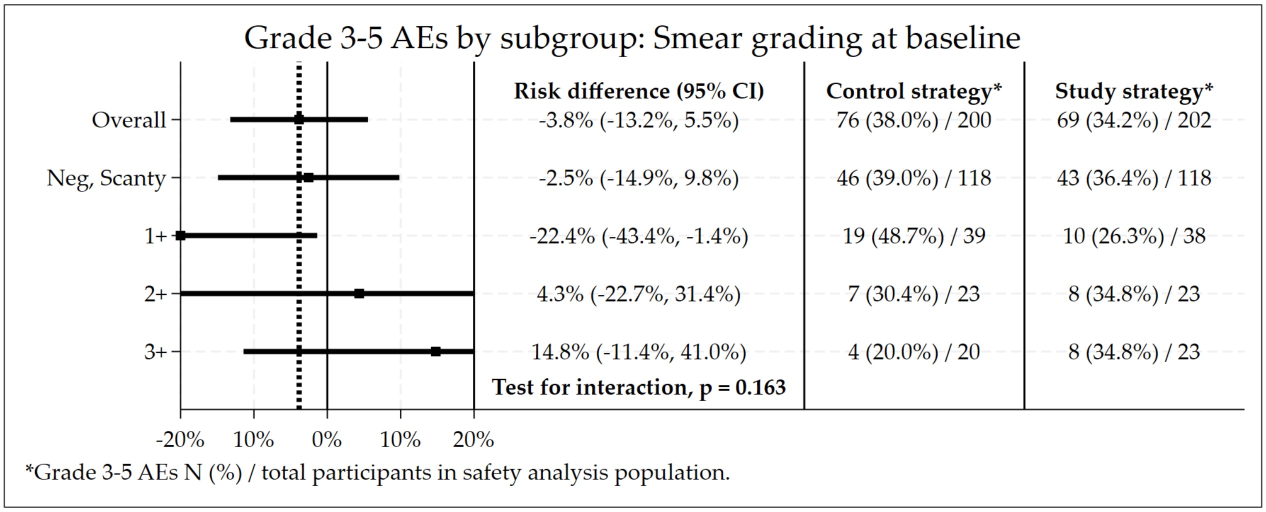

##### 3.12.15 Injectable Resistance at Baseline

Figure S3.55: Subgroup by injectable resistance: Non-inferiority plot of the primary efficacy outcome.

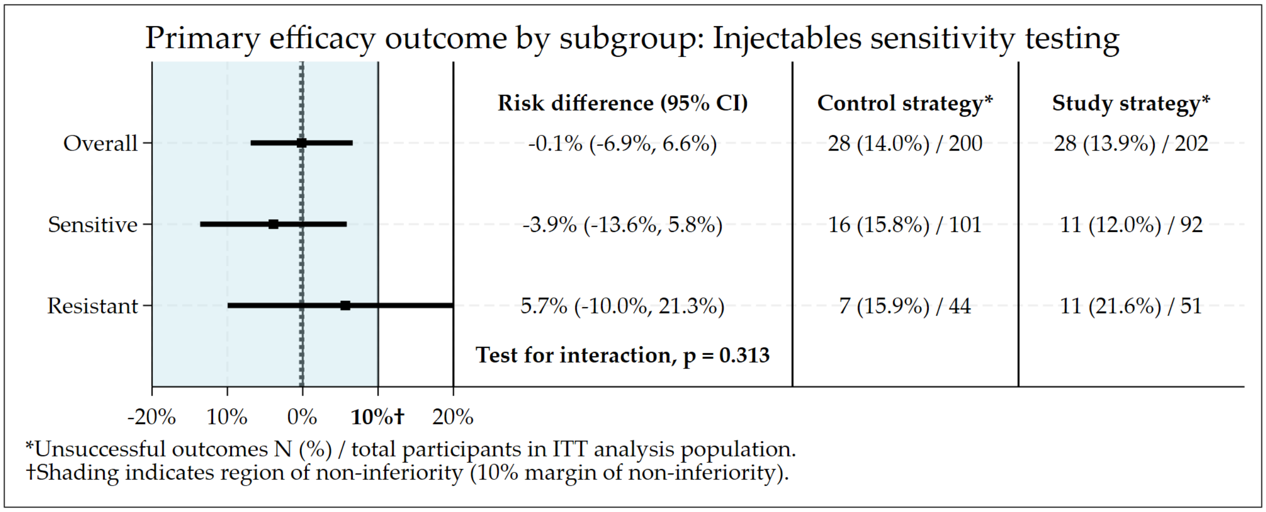

Figure S3.56: Subgroup by injectable resistance: Risk difference plot of the primary safety outcome.

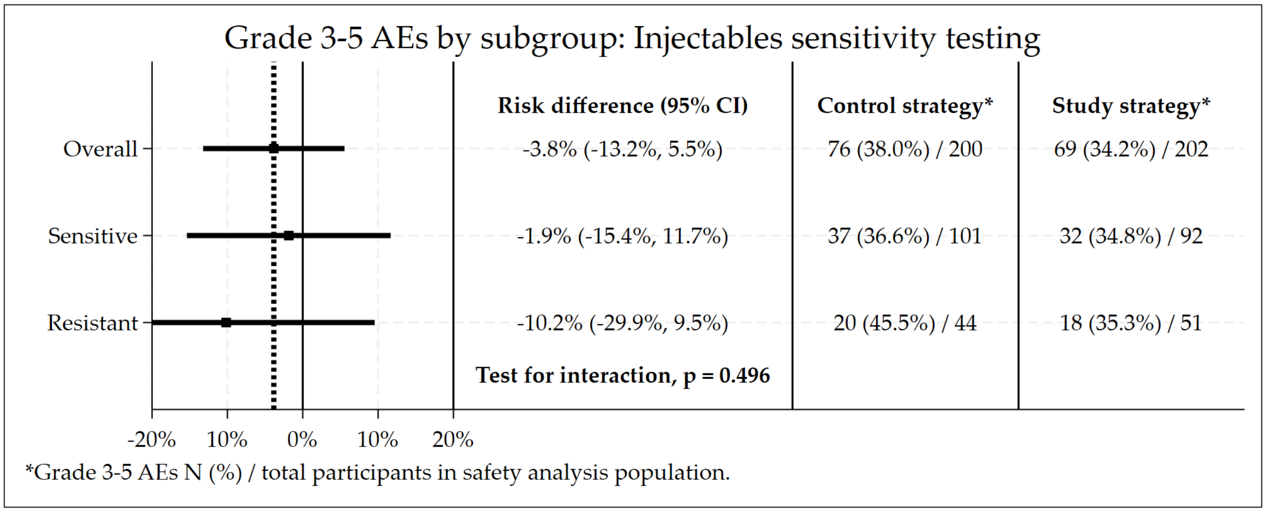

##### 3.12.16 Fluoroquinolone Resistance at Baseline

Figure S3.57: Subgroup by fluoroquinolone resistance: Non-inferiority plot of the primary efficacy outcome.

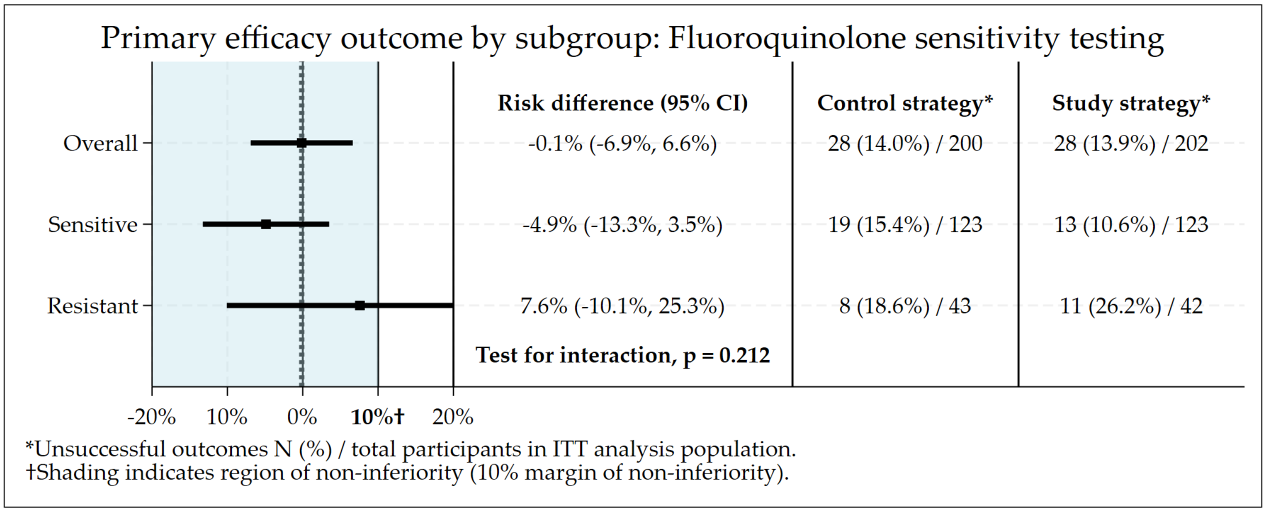

Figure S3.58: Subgroup by fluoroquinolone resistance: Risk difference plot of the primary safety outcome.

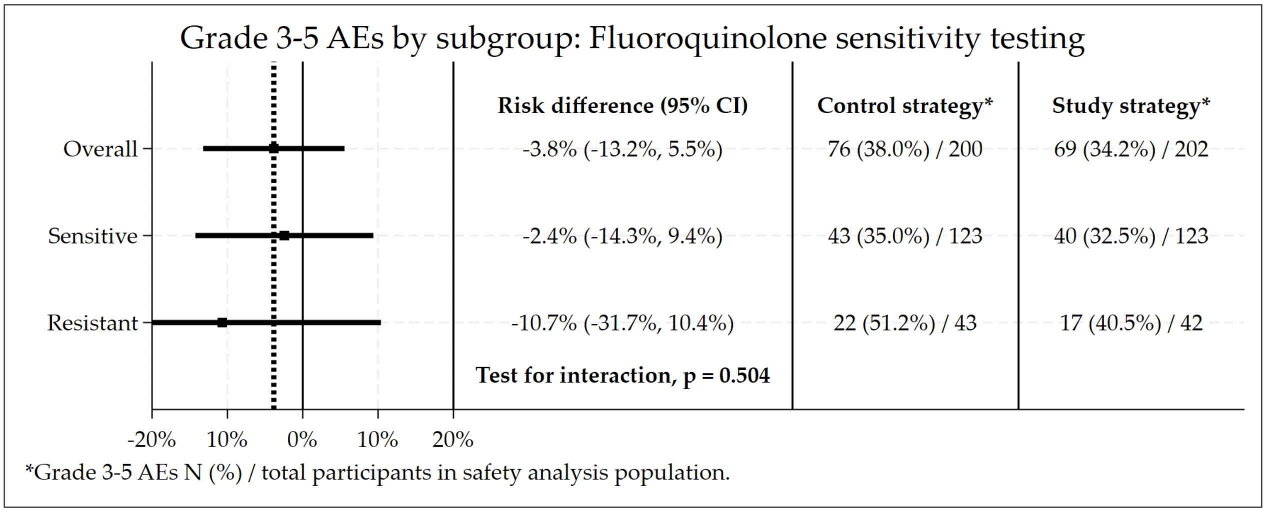

##### 3.12.17 Bedaquiline Resistance at Baseline

Figure S3.59: Subgroup by bedaquiline resistance: Non-inferiority plot of the primary efficacy outcome.

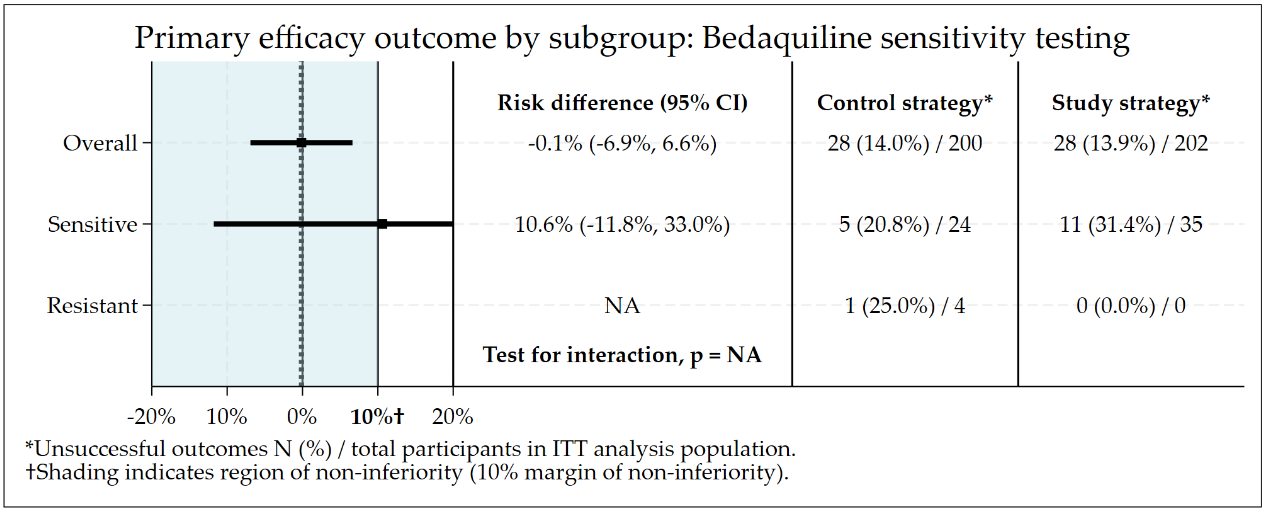

Figure S3.60: Subgroup by bedaquiline resistance: Risk difference plot of the primary safety outcome.

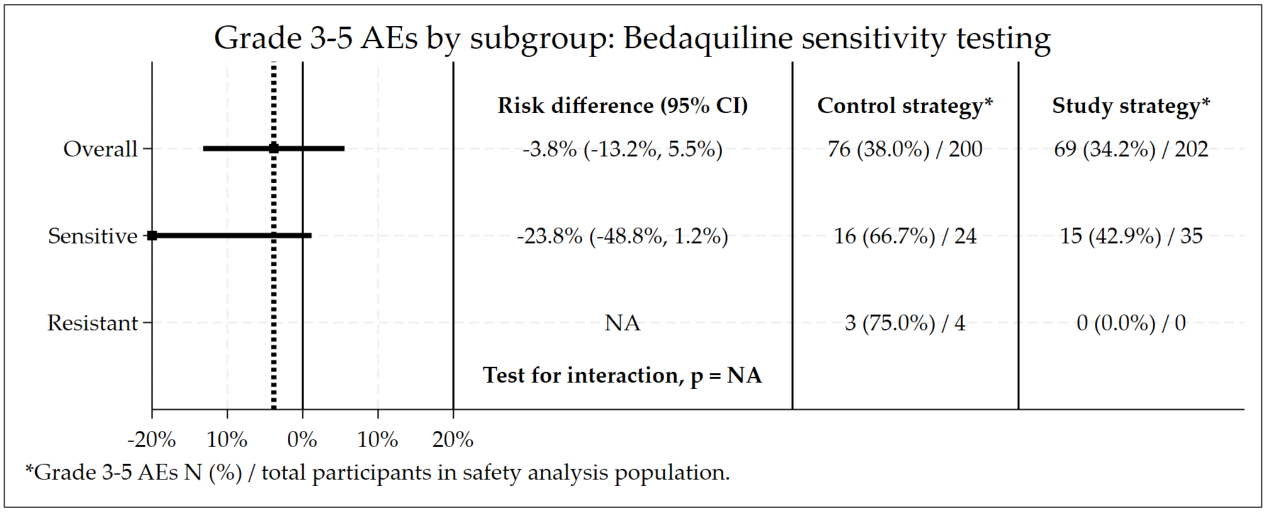

##### 3.12.18 Clofazimine Resistance at Baseline

Figure S3.61: Subgroup by clofazimine resistance: Non-inferiority plot of the primary efficacy outcome.

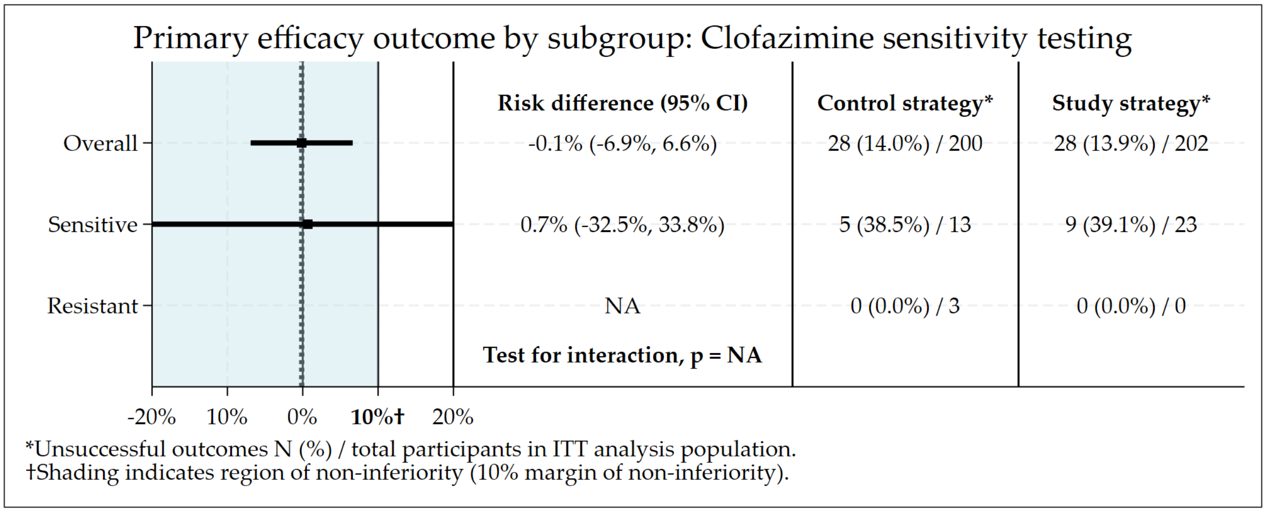

Figure S3.62: Subgroup by clofazimine resistance: Risk difference plot of the primary safety outcome.

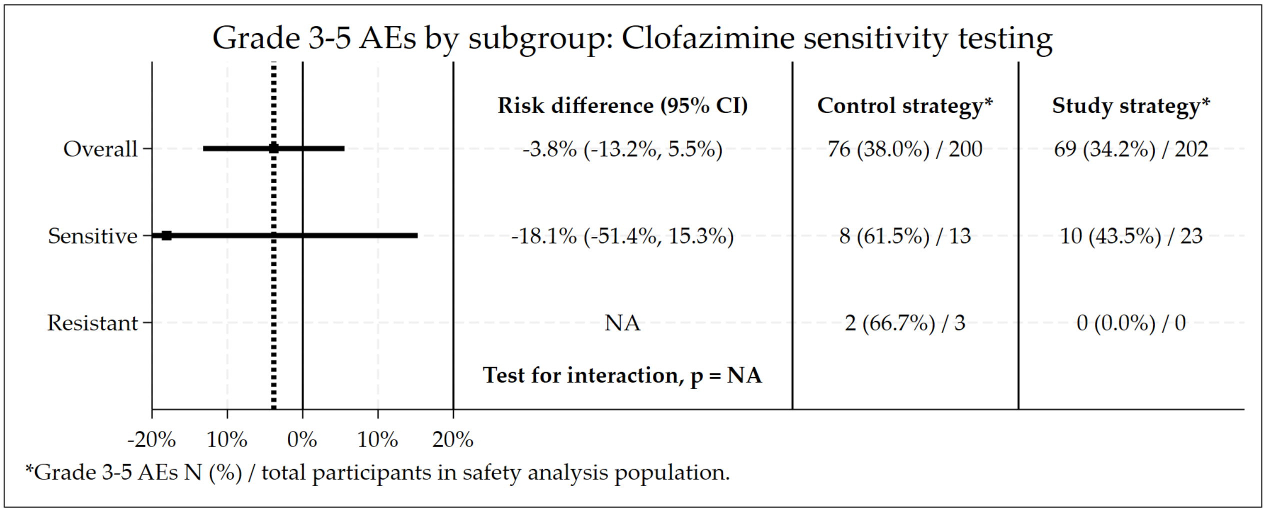

##### 3.12.19 Linezolid Resistance at Baseline

Figure S3.63: Subgroup by linezolid resistance: Non-inferiority plot of the primary efficacy outcome.

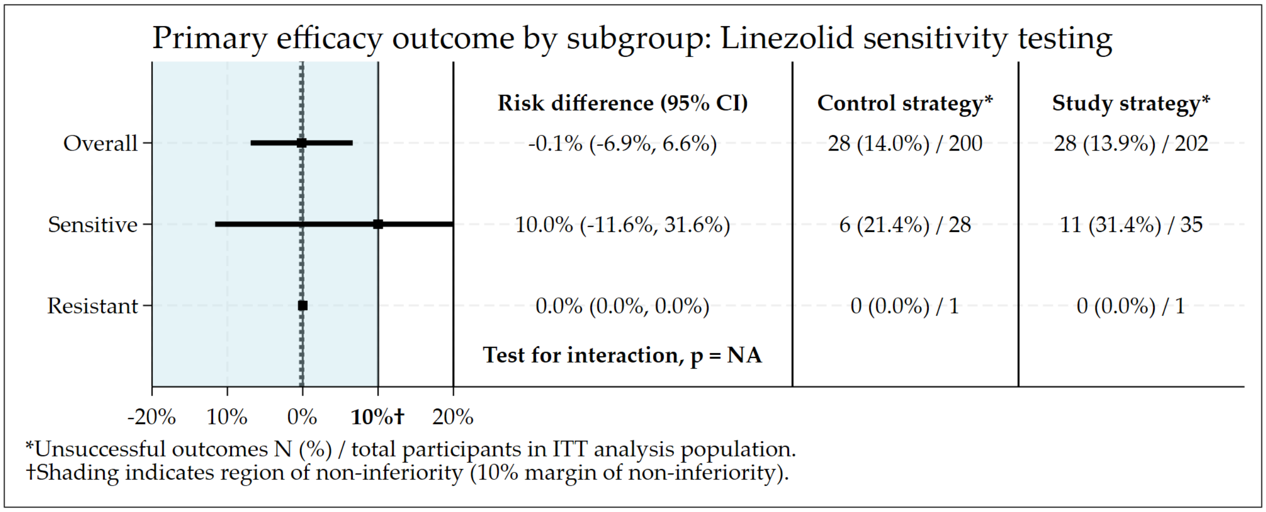

Figure S3.64: Subgroup by linezolid resistance: Risk difference plot of the primary safety outcome.

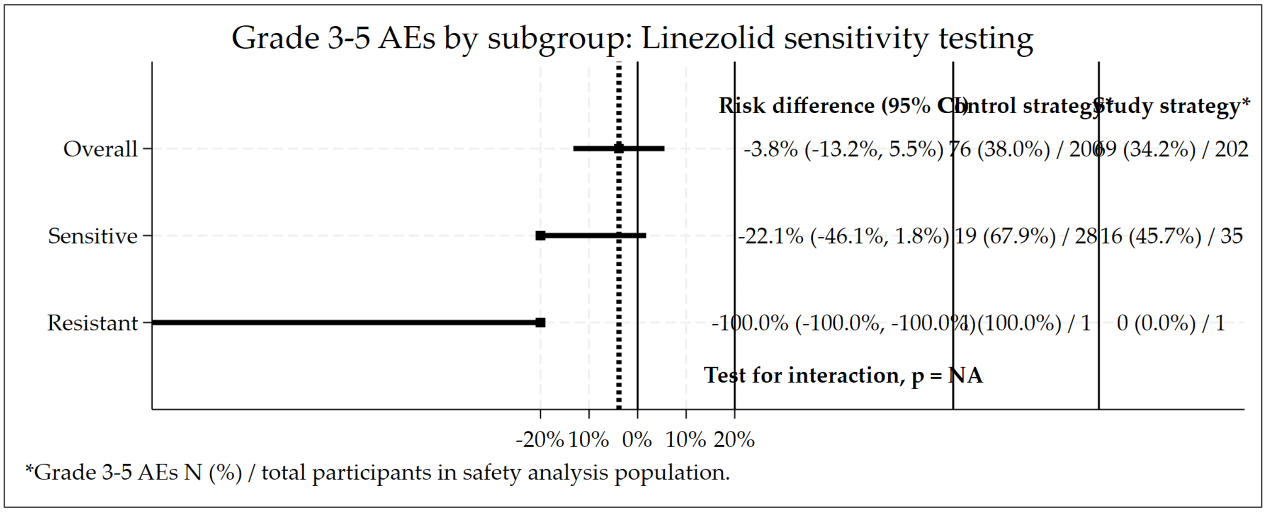

##### 3.12.20 Isoniazid Resistance at Baseline

Figure S3.65: Subgroup by isoniazid resistance: Non-inferiority plot of the primary efficacy outcome.

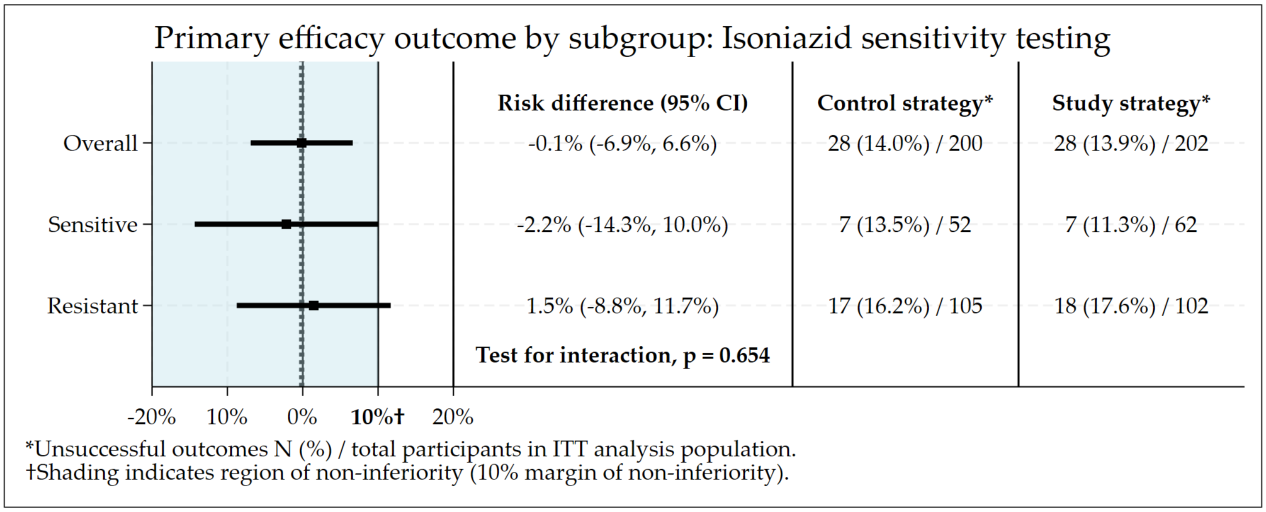

Figure S3.66: Subgroup by isoniazid resistance: Risk difference plot of the primary safety outcome.

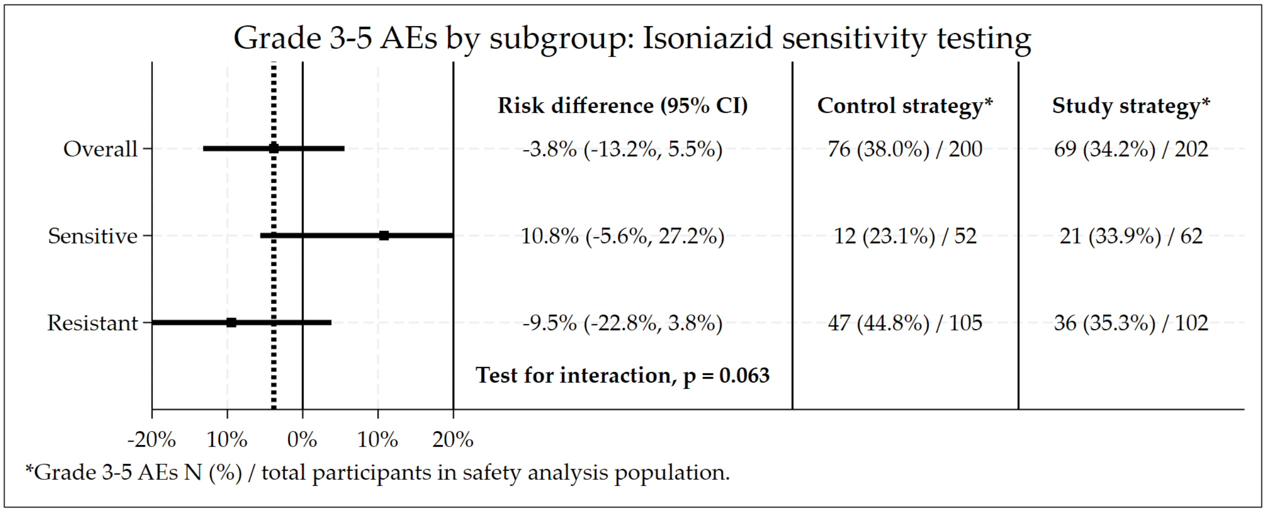

##### 3.12.21 Fluoroquinolone and Bedaquiline Resistance at Baseline

Figure S3.67: Subgroup by baseline resistance pattern: Non-inferiority plot of the primary efficacy outcome.

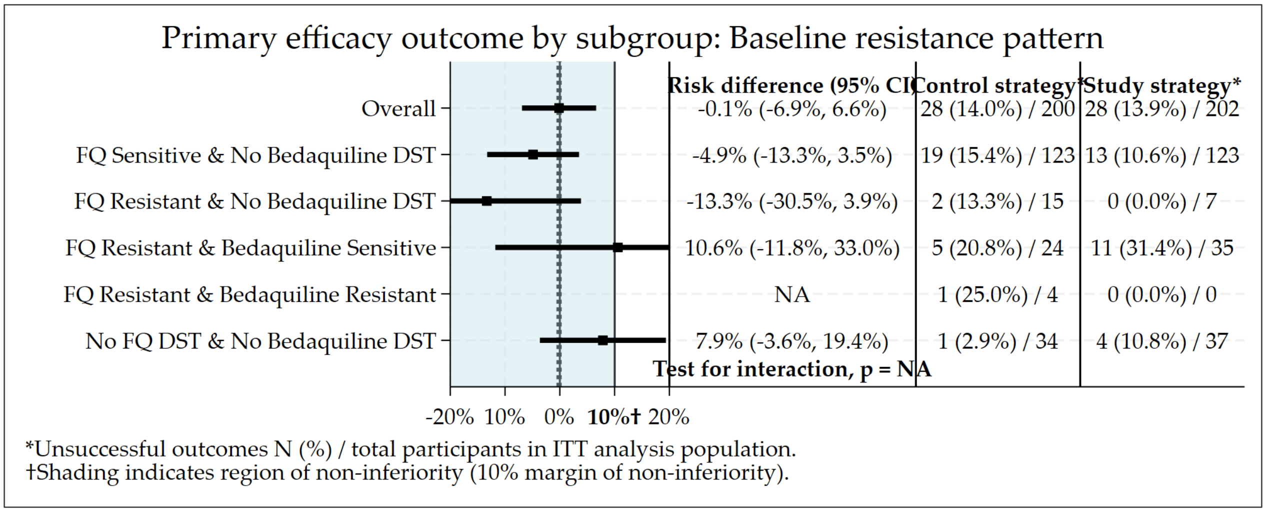

Figure S3.68: Subgroup by baseline resistance pattern: Risk difference plot of the primary safety outcome.

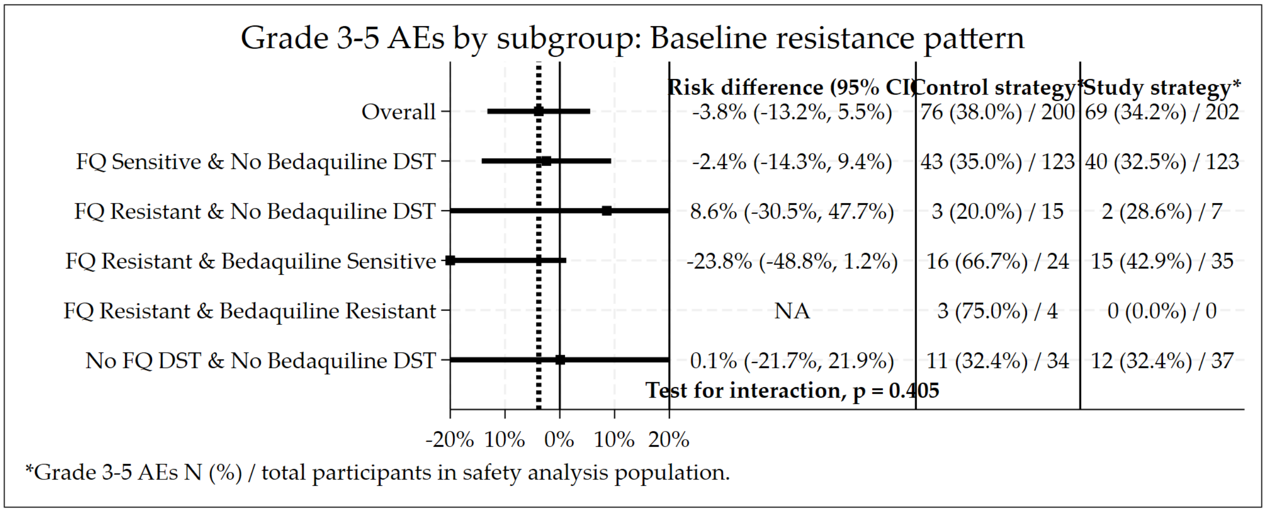

##### 3.12.22 ALT at Baseline

Figure S3.69: Subgroup by ALT (U/L): Non-inferiority plot of the primary efficacy outcome.

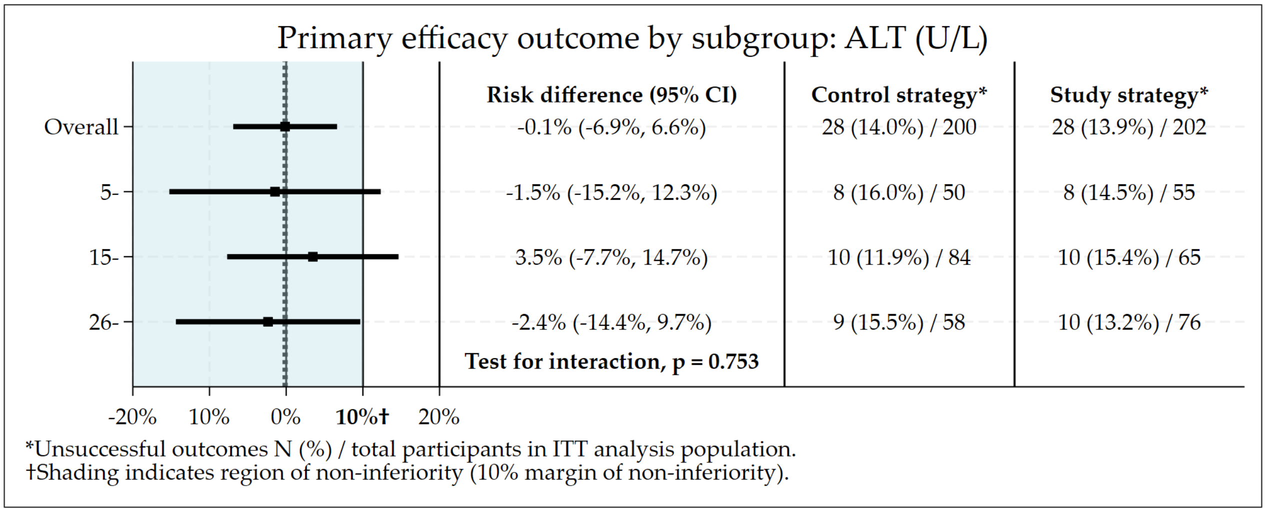

Figure S3.70: Subgroup by ALT (U/L): Risk difference plot of the primary safety outcome.

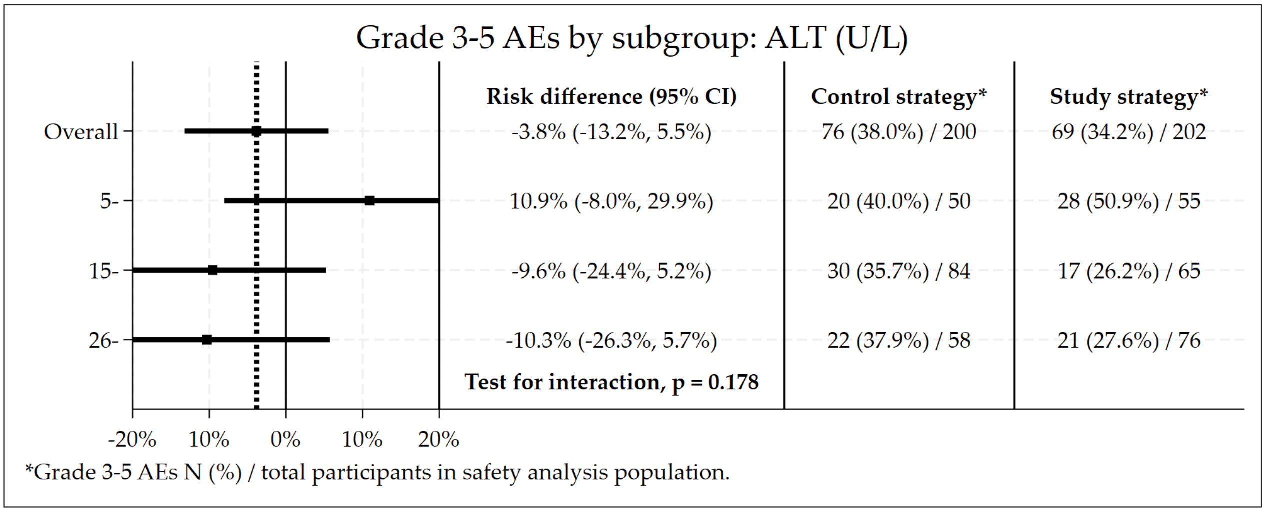

##### 3.12.23 Haemoglobin at Baseline

Figure S3.71: Subgroup by haemoglobin (g/dL): Non-inferiority plot of the primary efficacy outcome.

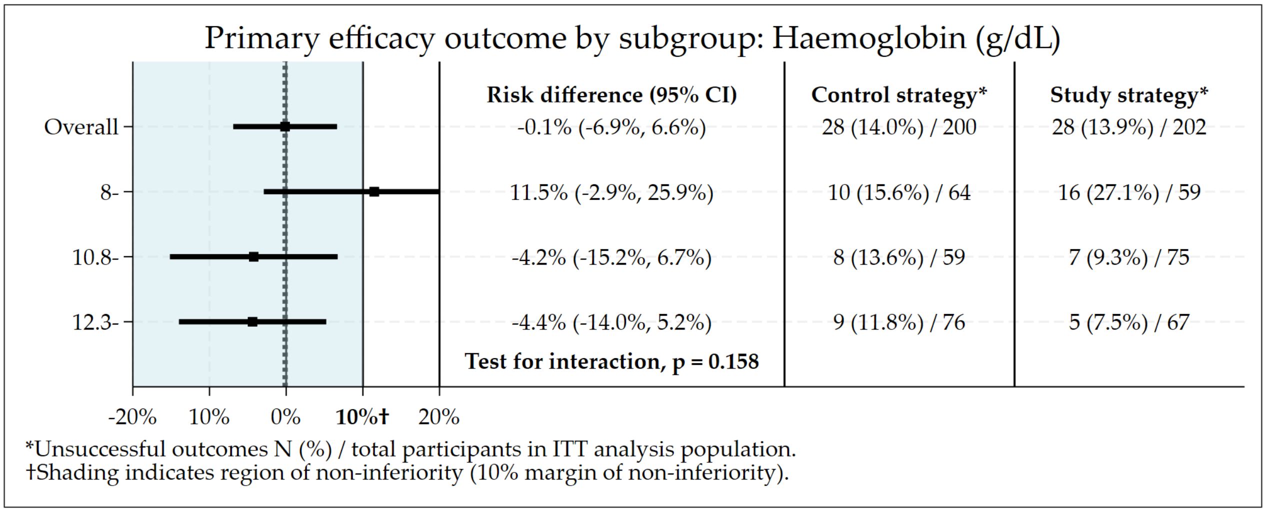

Figure S3.72: Subgroup by haemoglobin (g/dL): Risk difference plot of the primary safety outcome.

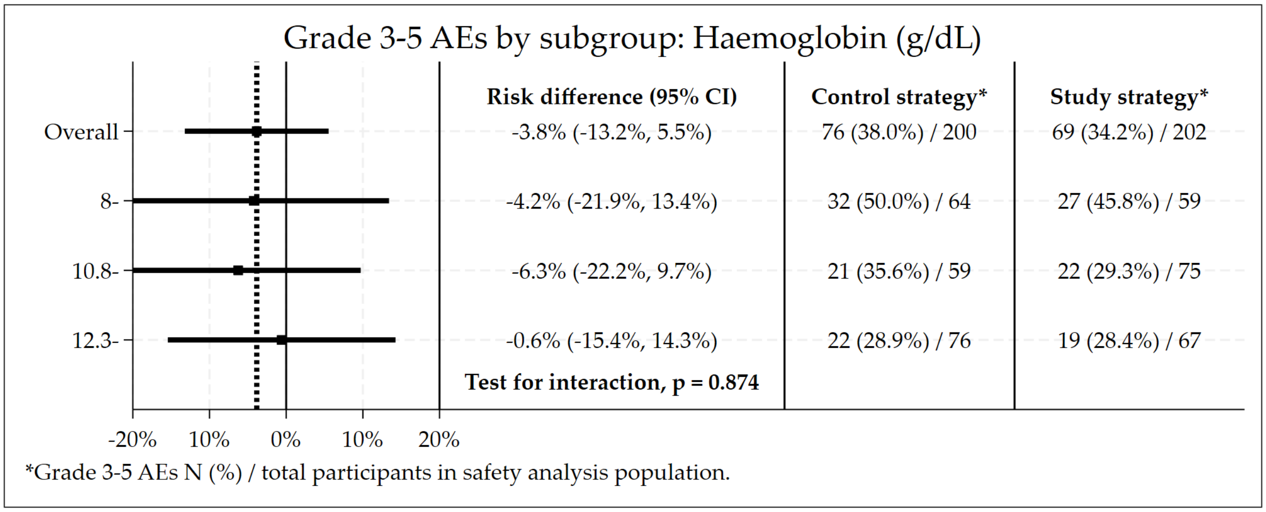

##### 3.12.24 Platelets at Baseline

Figure S3.73: Subgroup by platelets (x$10^9$/L): Non-inferiority plot of the primary efficacy outcome.

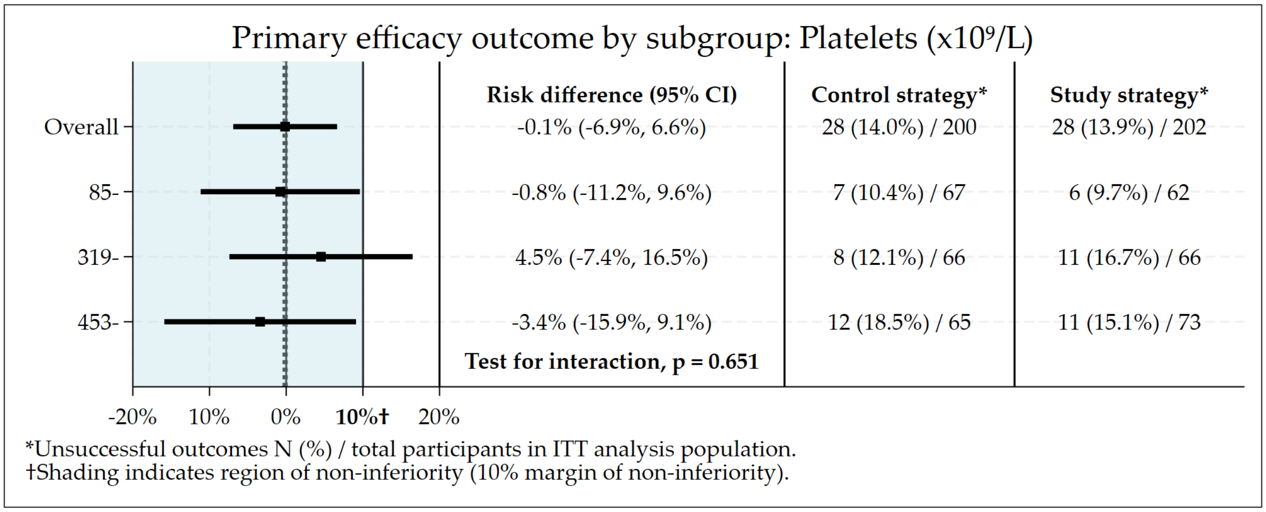

Figure S3.74: Subgroup by platelets (x$10^9$/L): Risk difference plot of the primary safety outcome.

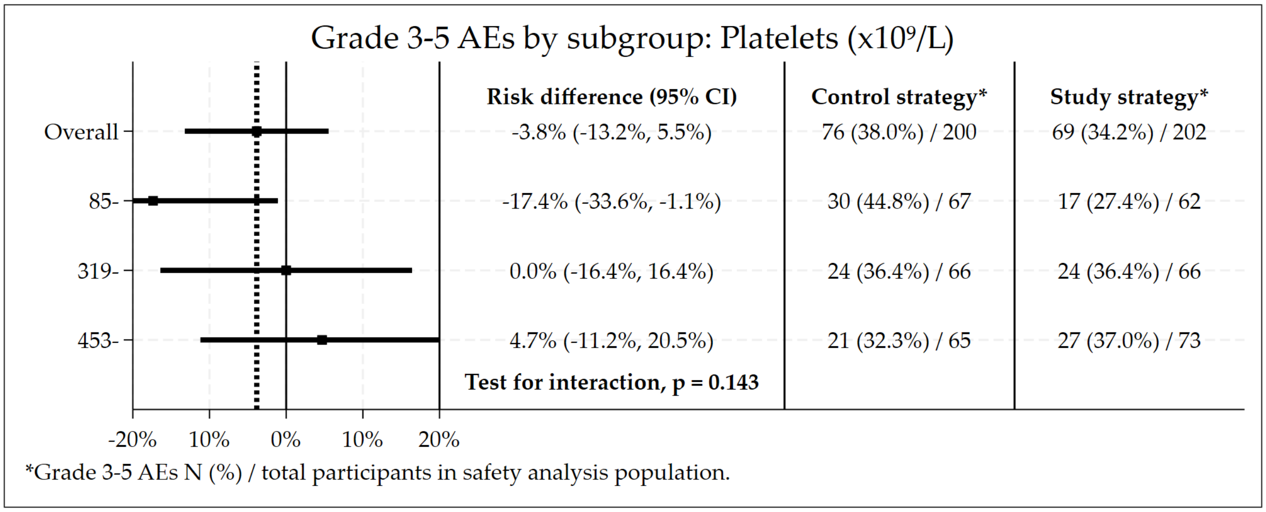

##### 3.12.25 Neutrophils at Baseline

Figure S3.75: Subgroup by neutrophils (x$10^9$/L): Non-inferiority plot of the primary efficacy outcome.

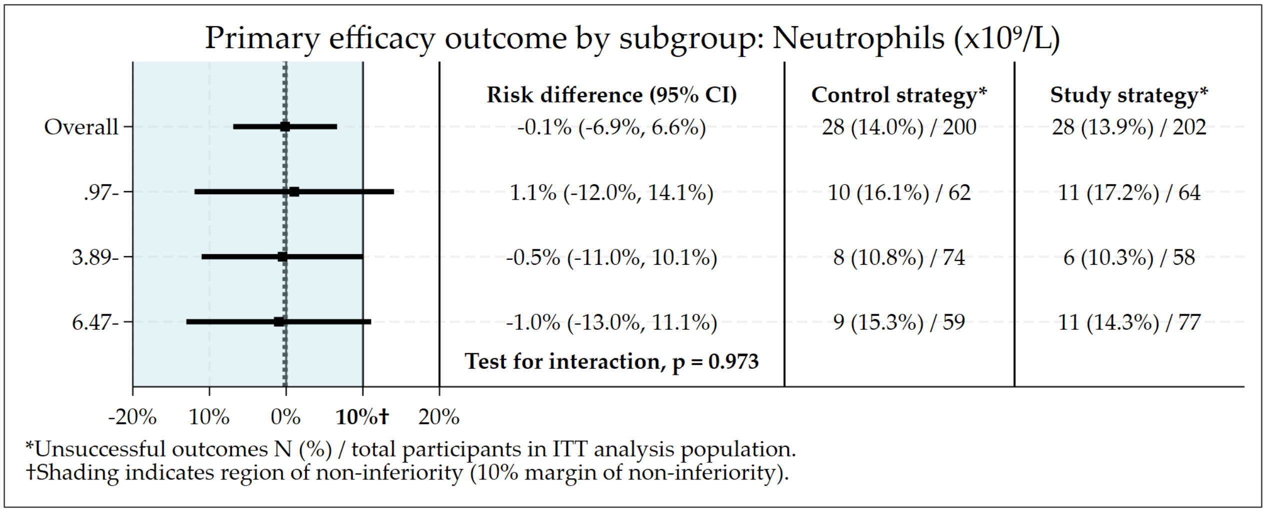

Figure S3.76: Subgroup by neutrophils (x$10^9$/L): Risk difference plot of the primary safety outcome.

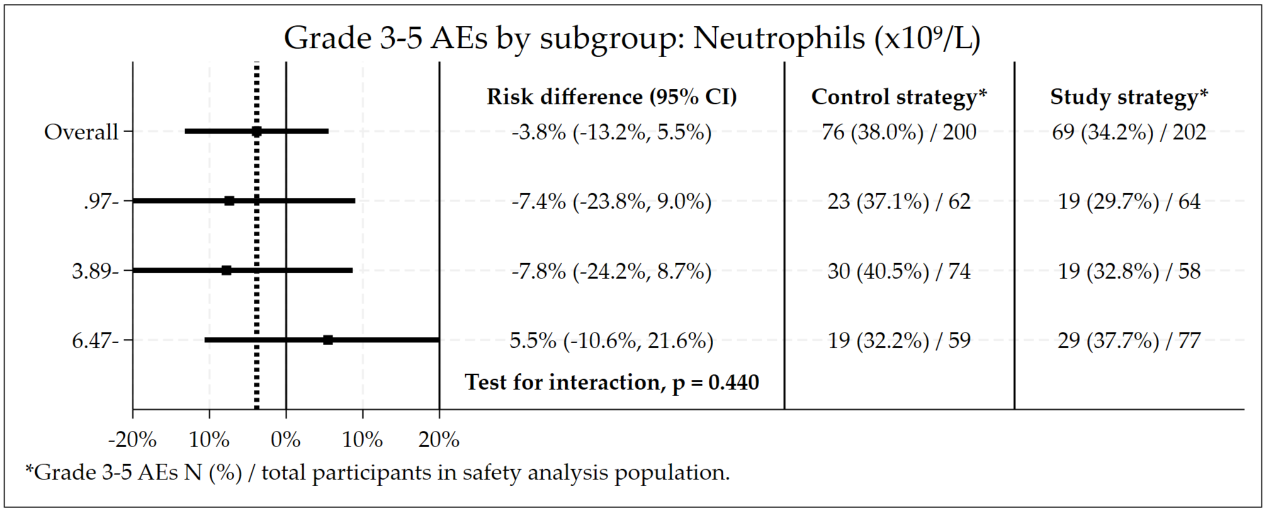

##### 3.12.26 Creatinine at Baseline

Figure S3.77: Subgroup by creatinine (mmol/L): Non-inferiority plot of the primary efficacy outcome.

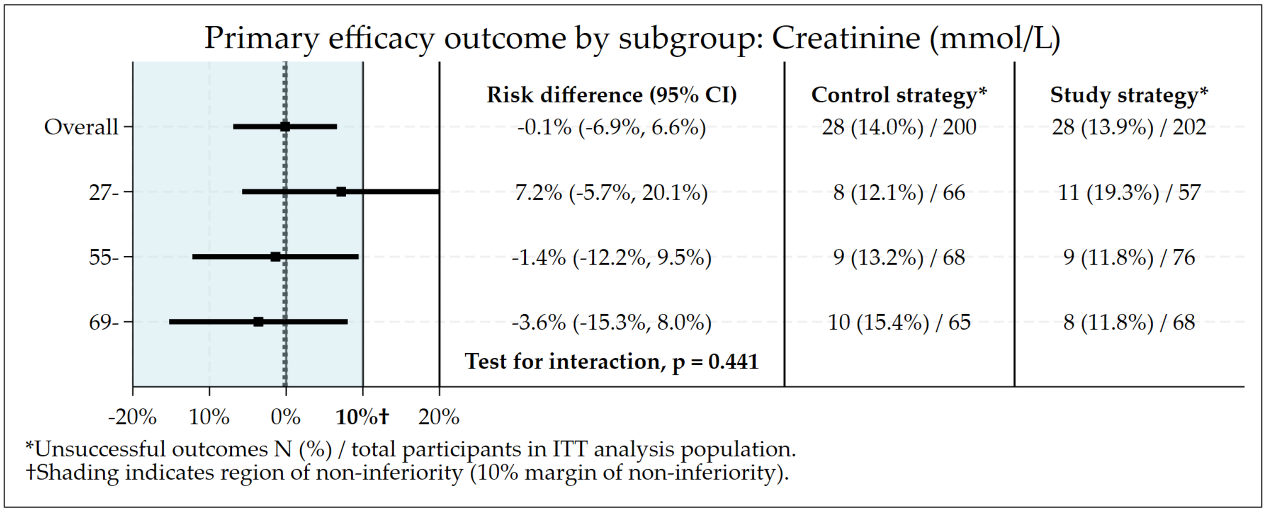

Figure S3.78: Subgroup by creatinine (mmol/L): Risk difference plot of the primary safety outcome.

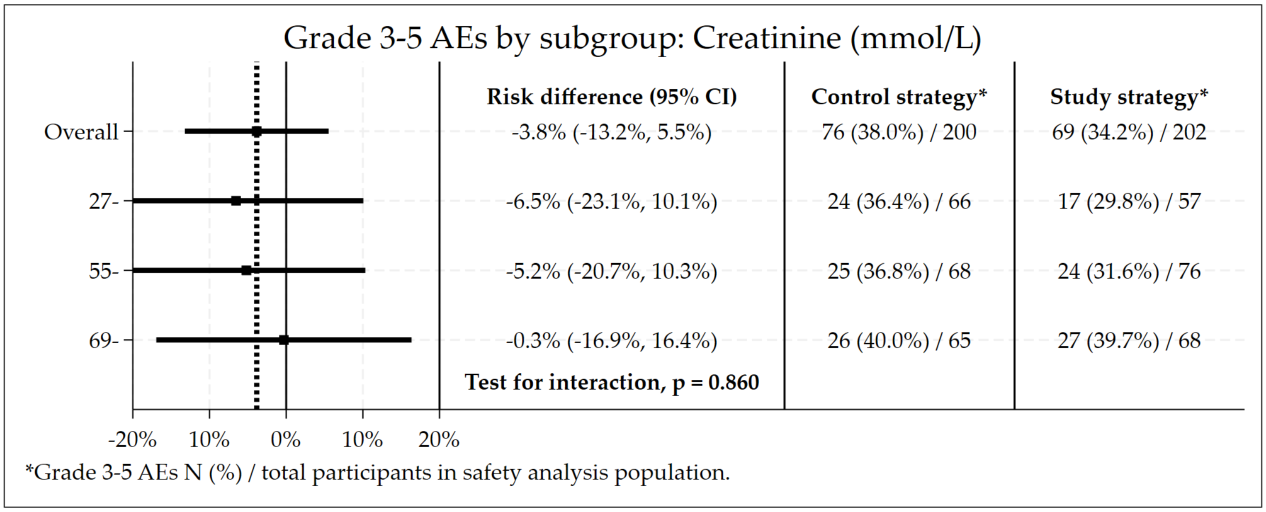

##### 3.12.27 Albumin at Baseline

Figure S3.79: Subgroup by albumin (g/L): Non-inferiority plot of the primary efficacy outcome.

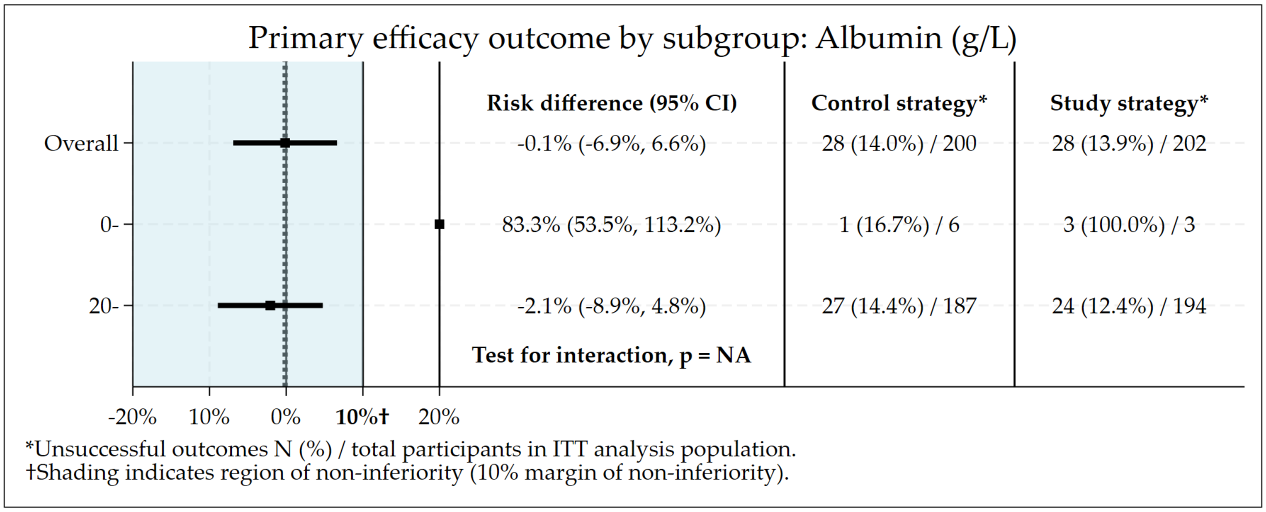

Figure S3.80: Subgroup by albumin (g/L): Risk difference plot of the primary safety outcome.

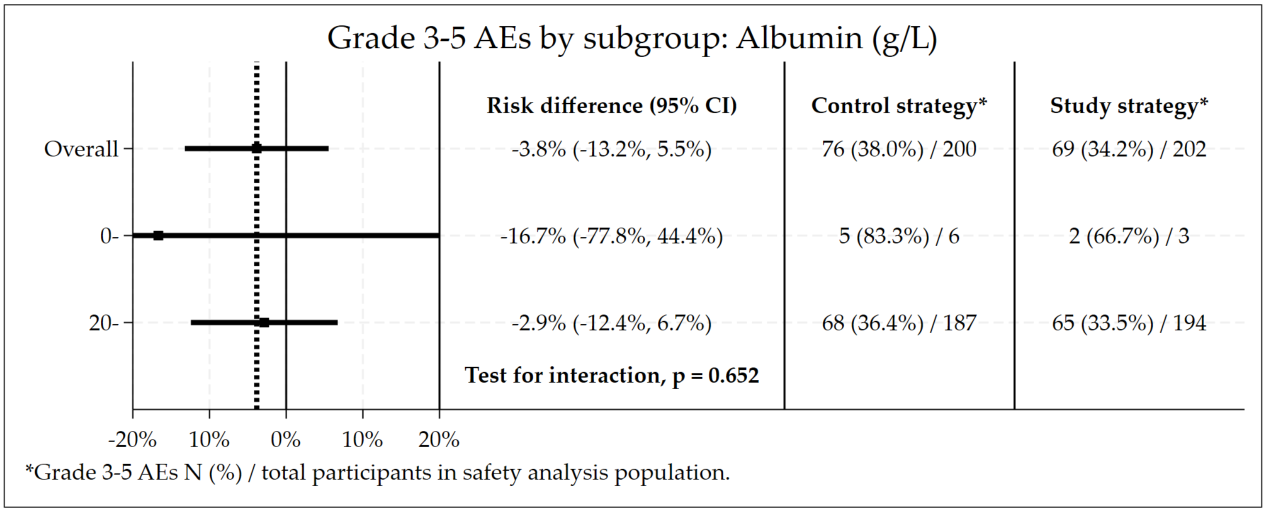

##### 3.12.28 Cavities on Chest X-ray at Baseline

Figure S3.81: Subgroup by cavities on chest x-ray: Non-inferiority plot of the primary efficacy outcome.

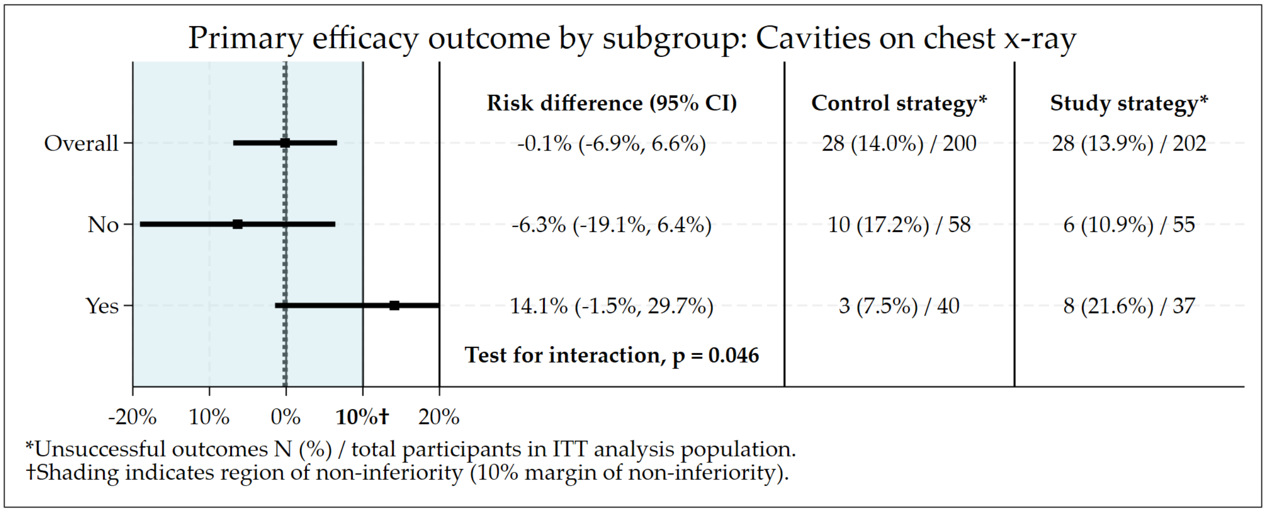

Figure S3.82: Subgroup by cavities on chest x-ray: Risk difference plot of the primary safety outcome.

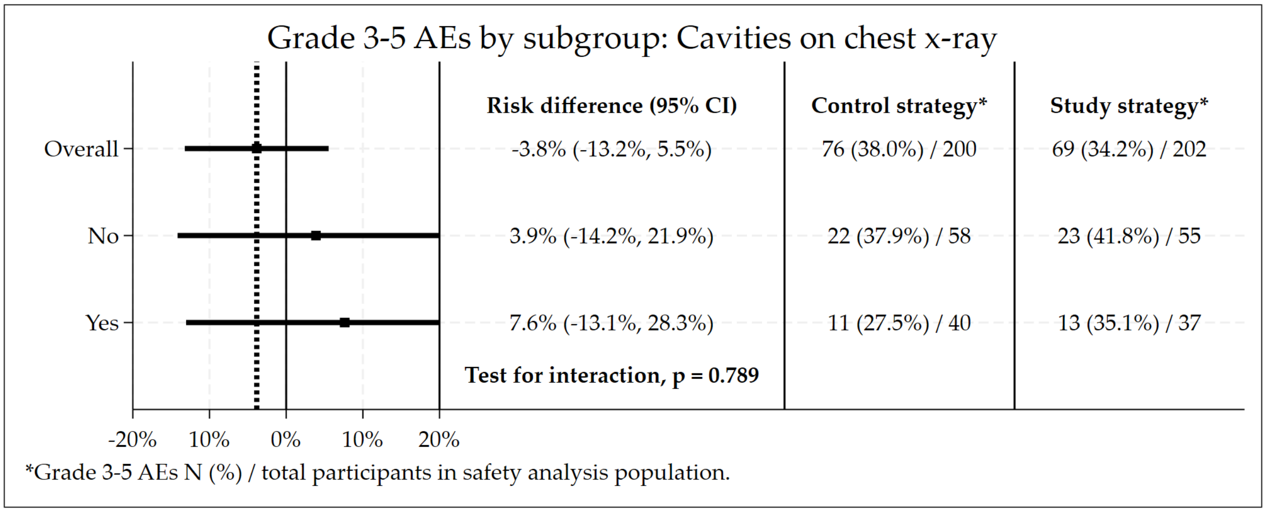

##### 3.12.29 Chest X-ray Results at Baseline

Figure S3.83: Subgroup by chest x-ray result: Non-inferiority plot of the primary efficacy outcome.

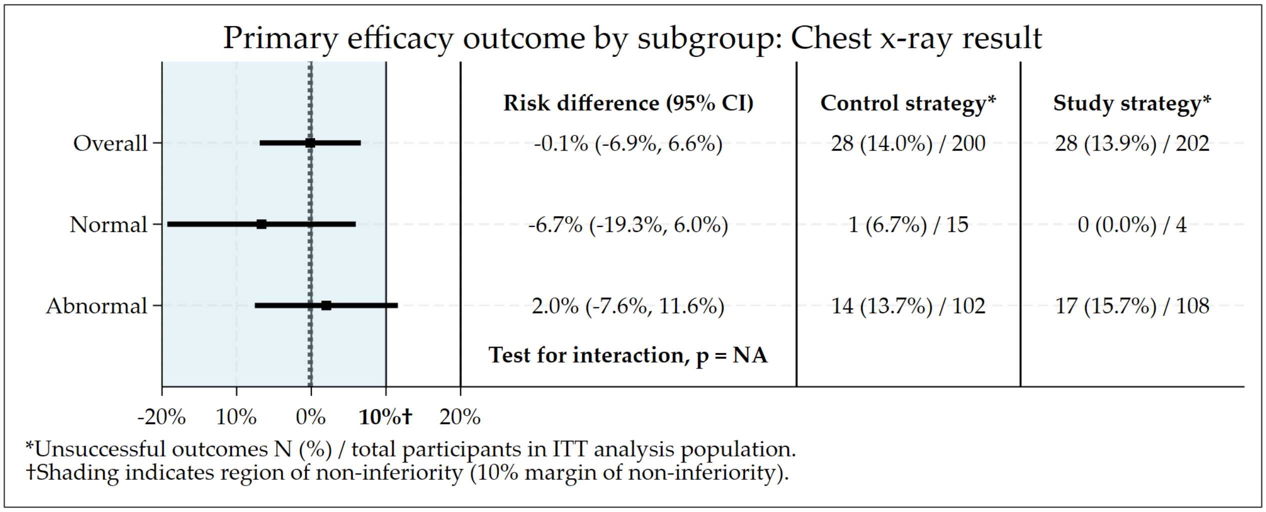

Figure S3.84: Subgroup by chest x-ray result: Risk difference plot of the primary safety outcome.

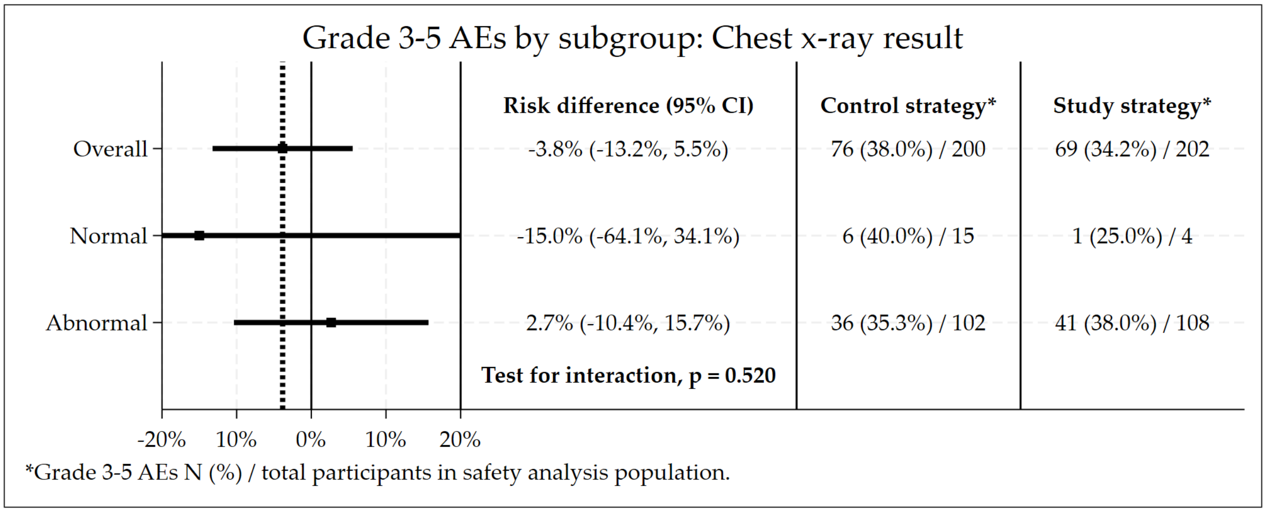

##### 3.12.30 Baseline Chest X-ray Consistent with TB

Figure S3.85: Subgroup by chest x-ray consistent with TB: Non-inferiority plot of the primary efficacy outcome.

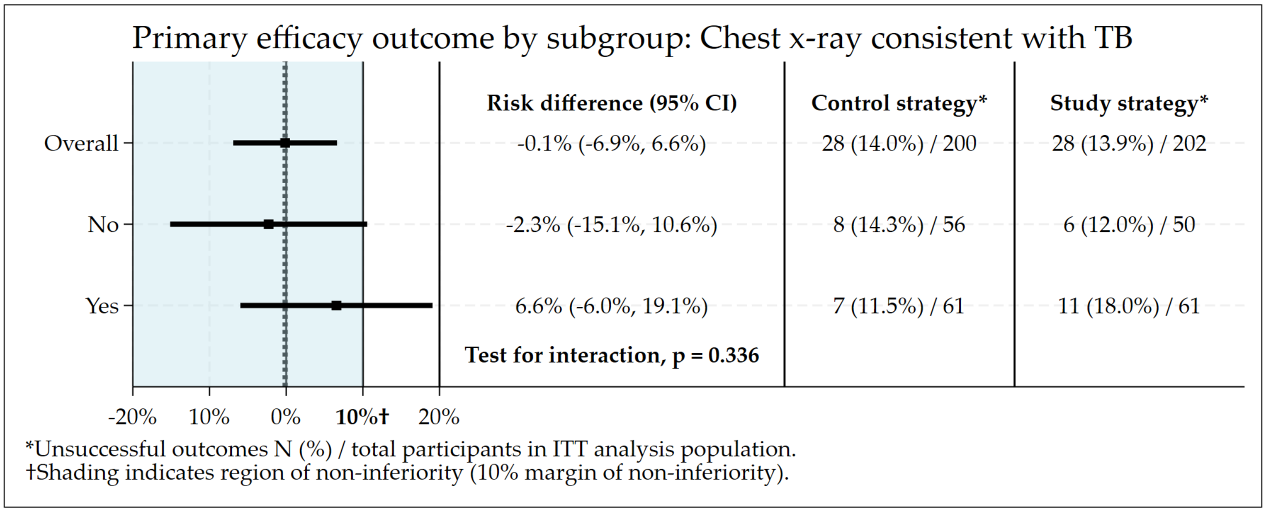

Figure S3.86: Subgroup by chest x-ray consistent with TB: Risk difference plot of the primary safety outcome.

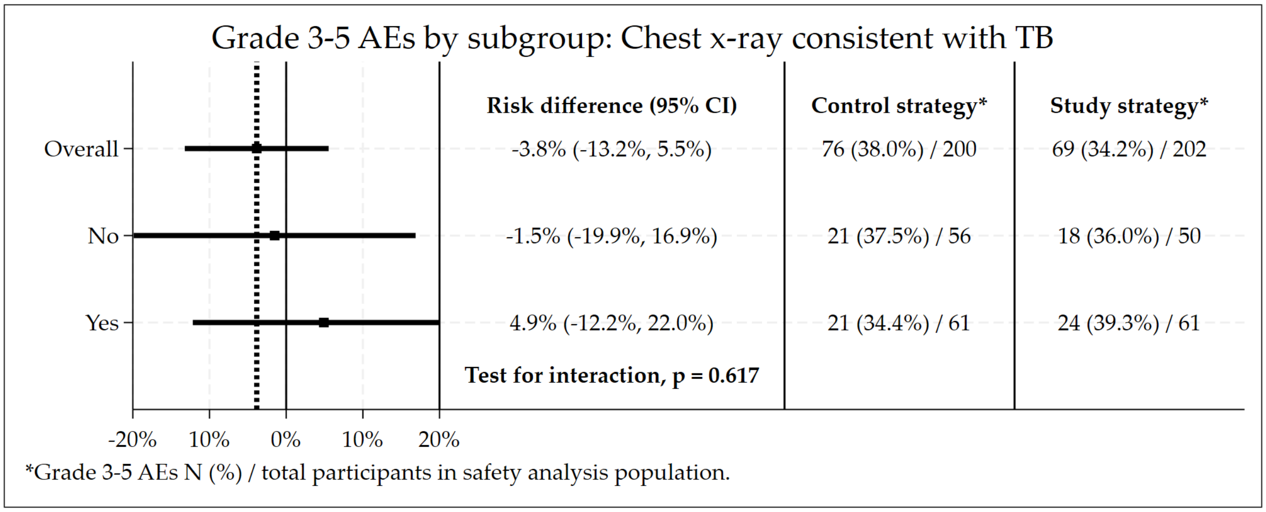

#### 3.13 Secondary Outcome: Composite efficacy and safety outcome

- Protocol-specified secondary composite efficacy and safety outcome.
- Restricted to participants that have been in the study for at least 76 weeks.

Table S3.26: Composite efficacy and safety outcome (only considering grade 3-4 AEs on treatment).

| Primary Outcome | Control Strategy | Study Strategy | Total |
|---|---|---|---|
| **Total randomized (ITT population)** | **200** | **202** | **402** |
| **No event** | **117 (58.5%)** | **127 (62.9%)** | **244 (60.7%)** |
| **Total Events** | **83 (41.5%)** | **75 (37.1%)** | **158 (39.3%)** |
| Grade 3-4 AE only | 55 (27.5%) | 47 (23.3%) | 102 (25.4%) |
| Both grade 3-4 AE and unsuccessful outcome | 19 (9.5%) | 16 (7.9%) | 35 (8.7%) |
| Unsuccessful outcome only | 9 (4.5%) | 12 (5.9%) | 21 (5.2%) |
| **Unadjusted difference** | | | **-4.4% (-13.9%, 5.2%)** |
| **Adjusted difference (HIV and site)** | | | **-4.4% (-13.8%, 5.1%)** |

Table S3.27: Composite efficacy and safety outcome (considering grade 3-4 AEs ocurring at any time during the study).

| Primary Outcome | Control Strategy | Study Strategy | Total |
|---|---|---|---|
| **Total randomized (ITT population)** | **200** | **202** | **402** |
| **No event** | **115 (57.5%)** | **123 (60.9%)** | **238 (59.2%)** |
| **Total Events** | **85 (42.5%)** | **79 (39.1%)** | **164 (40.8%)** |
| Grade 3-4 AE only | 57 (28.5%) | 51 (25.2%) | 108 (26.9%) |
| Both grade 3-4 AE and unsuccessful outcome | 19 (9.5%) | 18 (8.9%) | 37 (9.2%) |
| Unsuccessful outcome only | 9 (4.5%) | 10 (5.0%) | 19 (4.7%) |
| **Unadjusted difference** | | | **-3.4% (-13.0%, 6.2%)** |
| **Adjusted difference (HIV and site)** | | | **-3.4% (-12.9%, 6.1%)** |

#### 3.14 Detailed efficay and safety outcomes among children

##### 3.14.1 Baseline Characteristics

Table S3.28: Summary of baseline characteristics by strategy.

| | Control Strategy | Study Strategy | Total |
|---|---|---|---|
| **Total randomised** | | | |
| | 17 | 13 | 30 |
| **Age (years)** | | | |
| Median (IQR) | 16.0 (13.0, 16.0) | 16.0 (15.0, 17.0) | 16.0 (14.0, 17.0) |
| Min, Max | 8.0, 17.0 | 10.0, 17.0 | 8.0, 17.0 |
| **Weight (kg)** | | | |
| Median (IQR) | 46.7 (37.1, 50.2) | 42.8 (39.7, 47.2) | 45.4 (37.1, 50.2) |
| Min, Max | 25.1, 58.0 | 28.0, 70.4 | 25.1, 70.4 |
| **Gender** | | | |
| Male | 7 (41%) | 5 (38%) | 12 (40%) |
| Female | 10 (59%) | 8 (62%) | 18 (60%) |

Table S3.28: Summary of baseline characteristics by strategy. *(continued)*

| | Control Strategy | Study Strategy | Total |
|---|---|---|---|
| **HIV Status** | | | |
| HIV Negative | 13 (76%) | 12 (92%) | 25 (83%) |
| HIV Positive | 4 (24%) | 1 (8%) | 5 (17%) |
| **CD4 Count, categorical** | | | |
| <200 | 2 (12%) | 0 | 2 (7%) |
| ≥200 | 2 (12%) | 0 | 2 (7%) |
| HIV Neg | 13 (76%) | 12 (92%) | 25 (83%) |
| Missing | 0 | 1 (8%) | 1 (3%) |
| **Previous diagnosis of TB** | | | |
| None | 12 (71%) | 10 (77%) | 22 (73%) |
| DS-TB | 5 (29%) | 3 (23%) | 8 (27%) |
| **Duration of treatment at previous TB episode** | | | |
| <6 months | 2 (12%) | 2 (15%) | 4 (13%) |
| 6-12 months | 3 (18%) | 1 (8%) | 4 (13%) |
| No previous diagnosis | 12 (71%) | 10 (77%) | 22 (73%) |
| **Outcome of previous TB episode** | | | |
| Treatment completed | 1 (6%) | 2 (15%) | 3 (10%) |
| Cured | 4 (24%) | 0 | 4 (13%) |
| Treatment failed | 0 | 1 (8%) | 1 (3%) |
| No previous diagnosis | 12 (71%) | 10 (77%) | 22 (73%) |
| **CD4 Count** | | | |
| Median (IQR) | 242.0 (59.0, 397.0) | . (., .) | 242.0 (59.0, 397.0) |
| Min, Max | 12.0, 416.0 | ., . | 12.0, 416.0 |
| Number missing | 13 | 13 | 26 |

Table S3.29: Summary of baseline bacteriology by strategy.

| | Control Strategy | Study Strategy | Total |
|---|---|---|---|
| **Total randomised** | | | |
| | 17 | 13 | 30 |
| **Smear grading** | | | |
| Neg | 12 (71%) | 7 (54%) | 19 (63%) |
| 1+ | 2 (12%) | 5 (38%) | 7 (23%) |
| 2+ | 2 (12%) | 0 | 2 (7%) |
| 3+ | 1 (6%) | 1 (8%) | 2 (7%) |
| **MGIT culture result** | | | |
| Negative | 4 (24%) | 2 (15%) | 6 (20%) |
| Mtb complex | 13 (76%) | 11 (85%) | 24 (80%) |
| **Rifampicin Resistance** | | | |
| Resistant | 17 (100%) | 13 (100%) | 30 (100%) |
| **Isoniazid Resistance** | | | |
| Sensitive | 5 (29%) | 7 (54%) | 12 (40%) |
| Resistant | 8 (47%) | 5 (38%) | 13 (43%) |
| Missing | 4 (24%) | 1 (8%) | 5 (17%) |
| **Fluoroquinolone Resistance** | | | |
| Sensitive | 8 (47%) | 10 (77%) | 18 (60%) |
| Resistant | 5 (29%) | 2 (15%) | 7 (23%) |
| Indeterminate | 1 (6%) | 0 | 1 (3%) |
| Missing | 3 (18%) | 1 (8%) | 4 (13%) |
| **Linezolid Resistance** | | | |

| | | | |
|---|---|---|---|
| Sensitive | 4 (24%) | 2 (15%) | 6 (20%) |
| Missing | 13 (76%) | 11 (85%) | 24 (80%) |
| **Bedaquiline Resistance** | | | |
| Sensitive | 3 (18%) | 2 (15%) | 5 (17%) |
| Resistant | 1 (6%) | 0 | 1 (3%) |
| Missing | 13 (76%) | 11 (85%) | 24 (80%) |
| **Clofazimine Resistance** | | | |
| Resistant | 1 (6%) | 0 | 1 (3%) |
| Missing | 16 (94%) | 13 (100%) | 29 (97%) |
| **Injectables Resistance** | | | |
| Sensitive | 9 (53%) | 8 (62%) | 17 (57%) |
| Resistant | 3 (18%) | 2 (15%) | 5 (17%) |
| Indeterminate | 1 (6%) | 2 (15%) | 3 (10%) |
| Missing | 4 (24%) | 1 (8%) | 5 (17%) |

##### 3.14.2 Safety: Adverse Events

Table S3.30: Overall safety summary by strategy.

| | Control Strategy | Study Strategy | Total | RD (95% CI) |
|---|---|---|---|---|
| **Randomised and starting treatment** | **17** | **13** | **30** | |
| Grade 3-5 AEs | 7 (41.2%) | 3 (23.1%) | 10 (33.3%) | 18.1% (-14.6%, 50.8%) |
| Grade 3-5 AEs during treatment | 6 (35.3%) | 3 (23.1%) | 9 (30.0%) | 12.2% (-20.0%, 44.5%) |
| Grade 3-5 AEs during treatment, at least possibly related | 5 (29.4%) | 3 (23.1%) | 8 (26.7%) | 6.3% (-25.2%, 37.9%) |
| SAEs | 3 (17.6%) | 1 (7.7%) | 4 (13.3%) | 10.0% (-13.2%, 33.2%) |
| SAEs during treatment | 3 (17.6%) | 1 (7.7%) | 4 (13.3%) | 10.0% (-13.2%, 33.2%) |
| Notable Events (NEs) | 2 (11.8%) | 3 (23.1%) | 5 (16.7%) | -11.3% (-38.9%, 16.2%) |
| Death at any time | 0 (0.0%) | 0 (0.0%) | 0 (0.0%) | 0.0% (0.0%, 0.0%) |
| Death during treatment | 0 (0.0%) | 0 (0.0%) | 0 (0.0%) | 0.0% (0.0%, 0.0%) |
| Death after treatment | 0 (0.0%) | 0 (0.0%) | 0 (0.0%) | 0.0% (0.0%, 0.0%) |
| Anaemia leading to treatment discontinuation | 0 (0.0%) | 1 (7.7%) | 1 (3.3%) | -7.7% (-22.2%, 6.8%) |
| Anaemia leading to blood transfusion | 1 (5.9%) | 1 (7.7%) | 2 (6.7%) | -1.8% (-20.1%, 16.5%) |
| Grade 3-5 liver abnormality | 0 (0.0%) | 0 (0.0%) | 0 (0.0%) | 0.0% (0.0%, 0.0%) |
| Perihperal neuropathy leading to treatment discontinuation | 2 (11.8%) | 1 (7.7%) | 3 (10.0%) | 4.1% (-17.0%, 25.2%) |
| Optic neuropathy leading to treatment discontinuation | 1 (5.9%) | 1 (7.7%) | 2 (6.7%) | -1.8% (-20.1%, 16.5%) |
| QTcF ≥ 480ms | 1 (5.9%) | 0 (0.0%) | 1 (3.3%) | 5.9% (-5.3%, 17.1%) |
| QTcF ≥ 500ms | 0 (0.0%) | 0 (0.0%) | 0 (0.0%) | 0.0% (0.0%, 0.0%) |

- A **Notable event (NE)** is any one of the following:
  - Grade 3 and 4 cardiac adverse event
  - Grade 3 or 4 liver toxicity
  - One or more drugs may need to be suspended permanently due to severe toxicity
  - Pregnancy occurs in female participants during the trial (both on or off treatment)

Table S3.31: Summary of grade 3-5 AEs by MedDRA coding and strategy sorted by decreasing frequency.

| | Control Strategy | Study Strategy | Total |
|---|---|---|---|
| **Total randomized** | **17** | **13** | **30** |
| **No Grade 3-5 AE** | **10 (58.8%)** | **10 (76.9%)** | **20 (66.7%)** |
| **Any Grade 3-5 AE** | **7 (41.2%)** | **3 (23.1%)** | **10 (33.3%)** |
| Neuropathy peripheral | 3 (17.6%) | 1 (7.7%) | 4 (13.3%) |
| Anaemia | 2 (11.8%) | 1 (7.7%) | 3 (10.0%) |
| Optic neuritis | 1 (5.9%) | 1 (7.7%) | 2 (6.7%) |

Table S3.31: Summary of grade 3-5 AEs by MedDRA coding and strategy sorted by decreasing frequency. *(continued)*

| | **Control Strategy** | **Study Strategy** | **Total** |
|---|---|---|---|
| Neutropenia | 1 (5.9%) | 0 | 1 (3.3%) |
| Treatment failure | 1 (5.9%) | 0 | 1 (3.3%) |
| Weight decreased | 1 (5.9%) | 0 | 1 (3.3%) |

##### 3.14.3 Primary efficacy outcome

Table S3.32: Summary of week 76 primary efficacy outcome.

| **Primary Outcome** | **Control Strategy** | **Study Strategy** | **Total** |
|---|---|---|---|
| **Total randomized (ITT population)** | **17** | **13** | **30** |
| **Successful outcome at end of treatment and follow-up** | | | |
| **Total** | **17 (100.0%)** | **13 (100.0%)** | **30 (100.0%)** |
| Cured at end of treatment, and end of follow-up | 17 (100.0%) | 13 (100.0%) | 30 (100.0%) |
